# Bezisterim-associated anti-inflammatory epigenetic modulation of age acceleration and Alzheimer’s disease genes

**DOI:** 10.1101/2025.10.20.25338385

**Authors:** Christopher Reading, Jiayan Yan, Clarence Ahlem, Penelope Markham, Stephen O’Quinn, Joseph M. Palumbo, Varun B. Dwaraka

**Affiliations:** BioVie Inc., Carson City, NV, USA; Perissos, Inc., Wake Forest, NC, USA; TruDiagnostic, Inc., Lexington, KY, USA

**Keywords:** Aging, Alzheimer’s disease, bezisterim, biological clocks, DNA methylation, epigenetics, neuroinflammatory diseases

## Abstract

Treatments with the ability to slow or reduce biological age have therapeutic potential in diseases of aging, including late-onset Alzheimer’s disease (AD). We previously reported that bezisterim, a novel anti-inflammatory insulin sensitizer, modulated epigenetic age acceleration (EAA) in a randomized, placebo-controlled, 30-week AD trial. Here, we expand on those findings through integrative mechanistic analyses linking bezisterim-induced EAA changes with clinical outcomes. Thirty weeks of bezisterim treatment in patients with mild-to-moderate AD showed favorable trends for reduced EAA across 13 independent biological clocks versus placebo. The reduced EAA was predominantly associated with inflammation, cognition, and transcription factor genes that orchestrate broader gene networks. Genome-wide methylation profiling revealed 2581 genes with significant differential promoter methylation (DPM) between the bezisterim and placebo groups. We identified 447 of these as having potentially beneficial DPM based on expected expression related to published aging and AD activities; 179 were AD hub genes.

In addition, more than 1000 bezisterim treatment–related, potentially beneficial differential promoter methylation (PBDPM) genes associated with microglial neuroinflammation, pro-inflammatory kinase activity, cognitive decline, lipid metabolism, and transcriptional regulation were correlated with directional improvement in individual neurologic and metabolic clinical measures. The observed changes in PBDPM genes might contribute to the previously reported clinical effects of bezisterim in AD.

Bezisterim appears to exert pleiotropic effects through coordinated modulation of aging-related epigenetic programs, potentially counteracting epigenetic-driven neurodegenerative processes at the intersection of inflammation, metabolism, and transcriptional control.

## 1. Introduction

The pathogenesis and progression of Alzheimer’s disease (AD) have historically been attributed to the accumulation of amyloid β (Aβ) plaques and neurofibrillary tangles, which are believed to trigger innate immune responses to damage-associated molecular patterns (DAMPs), accelerate aging, and alter transcriptional profiles (Breijyeh & Karaman, 2020; Deczkowska et al., 2018; Srinivasan et al., 2020; Wei & Li, 2022; J. Zhang et al., 2024). However, neuroinflammation has recently emerged as a third core pathology of AD whereby sustained inflammatory signaling contributes to neuronal dysfunction and atrophy in cognitive disorders (Heneka et al., 2015; Jin, Chan, Wu, & Lee, 2022; Kamila et al., 2025; Y. Ma et al., 2023).

Multiple genome-wide association studies have implicated microglia as the primary cell type through which late-onset AD genetic risk is mediated. Functional analysis of gene variants suggests alterations in microglial metabolism, inflammatory signaling, lipid handling, phagocytosis, and clearance of Aβ in disease progression. In those with sporadic late-onset AD, progression appears to be driven by several factors, including accelerated aging (Thomas, Lehn, Janssen, Hildeman, & Chougnet, 2022); proinflammatory nuclear factor kappa B (NF-κB) signaling (Pan et al., 2011); metabolic dysfunction/deregulation of oxidative phosphorylation; anaerobic glycolysis (Piers et al., 2020); and glial insulin resistance (Alassaf & Rajan, 2023). These all disrupt Aβ phagocytosis, contributing to AD disease progression. The switching of microglia from a proinflammatory M1 state to an anti-inflammatory, restorative M2 state (Laudati, Curro, Navarra, & Lisi, 2017) to induce microglial phagocytosis and clearance of Aβ (Lee, Schuchman, Jin, & Bae, 2012; W. Ma et al., 2023) is highly dependent on microglial metabolism and disrupted by the inability to switch between oxidative phosphorylation and glycolysis. It is possible that bezisterim facilitates this switch to the M2 phenotype in excessive proinflammatory environments driven by aging, metabolic dysregulation, and AD disease progression.

Inflammatory signaling pathways involved in normal aging processes become dysregulated in AD and other neurocognitive disorders. The mechanisms by which this happens remain to be elucidated, but they likely involve the persistence of immune activity via DAMPs (Ma, Jiang, & Zhou, 2024). Activated (M1) macrophages and microglia are associated with the “Warburg effect” that favors glycolysis over oxidative phosphorylation (Liberti & Locasale, 2016). The resulting metabolic inflammation impairs insulin signaling, creating glycemic and lipid dysregulation, pro-inflammatory kinase cascades, chromatin remodeling, and altered transcription factor (TF) activity. Notably, altered insulin signaling contributes to chronic inflammation in other diseases of aging, including diabetes, obesity, and cardiovascular disease. For example, peripheral impaired glucose tolerance and type 2 diabetes (T2D) can trigger insulin resistance (IR) in the brain (Chow et al., 2019), increasing Aβ, phosphorylated Tau (pTau), oxidative stress, advanced glycation end products, and apoptosis (Rorbach-Dolata & Piwowar, 2019), suggesting a shared immune-mediated mechanism of neurodegeneration across some diseases of aging.

Epigenetic modification of these inflammatory pathways has been identified as a key regulator of disease progression in AD (Gao et al., 2022; Kinney et al., 2018; Y. Ma et al., 2023). Thus, novel therapies that can target epigenetic markers and decrease age-related inflammation may alter AD progression. Bezisterim, which has been shown to have a favorable safety profile, is an oral, blood-brain barrier–permeable, anti-inflammatory, insulin-sensitizing analog of the neurosteroid androst-5-ene-3β,7β,17β-triol (beta-AET) (Ahlem et al., 2011; C. Reading et al., 2025; C. L. Reading, Flores-Riveros, Stickney, & Frincke, 2013; C. L. Reading, Stickney et al., 2013). We previously reported an exploration of potentially beneficial differences in neurologic and metabolic measures, as well as DNA methylation (DNAm) measures, between bezisterim-and placebo-treated patients with mild-to-moderate AD (C. Reading et al., 2025). Here, we present findings from an epigenetic analysis of plasma DNA samples from the aforementioned study. The findings identify significant treatment-related differential promoter methylation (DPM) of genes associated with changes in expression and reported relationships to metabolic inflammation, and epigenetic changes influencing inflammatory chromatin reorganization (i.e., histone modifications) (Lin et al., 2022) and TF alterations in gene expression. Finally, we characterize epigenetic-driven realignment of gene promoter methylation (PM) correlations with clinical measures.

## 2. Materials and Methods

### 2.1. DNAm assessment

Whole-blood specimens from the completion visit were collected in EDTA tubes, frozen and shipped to the Epigenetic Clock Development Foundation for processing on Illumina Infinium MethylationEPIC v2.0 (EPICv2) BeadChips, as was previously described (C. Reading et al., 2025). Briefly, blinded samples were randomized to chips and processed in two batches using the standard Illumina Infinium workflow; 37 samples were assayed and 33 passed Illumina quality control for downstream DNAm analyses. IDAT binary files were generated and sent to TruDiagnostic for further DNAm data processing.

### 2.2. DNAm data preprocessing (minfi-based)

R raw IDATs were read with minfi (read.metharray) and assigned the EPIC v2.0 manifest (IlluminaHumanMethylationEPICv2; 20a1.hg38) (Aryee et al., 2014). A per-run quality control report was generated. Probe-level detection *P* values were computed from methylated + unmethylated signals (detectionP, type = “m+u”). Samples with mean detection *P* > 0.05 were flagged *a priori* for exclusion; no samples were removed in this dataset, and 5’-cytosine-phosphate-guanine-3’ (CpG)-level filtering at the same threshold was evaluated but not applied. Background correction and dye-bias normalization was performed using Noob (preprocessNoob, dyeMethod = “single”), after which beta values were exported (Fortin, Triche, & Hansen, 2017). Probe intensities were collapsed to EPIC v2.0 pfx-level targets to harmonize duplicated probe families to allow for compatibility for clock quantification. For legacy models requiring HM450/EPIC v1.0 CpGs, missing sites were augmented using array- and tissue-specific medians from sesame’s imputation defaults (EPIC.imputationDefault,Blood), ensuring coverage of required loci across methods. The resulting normalized beta matrix and corresponding RGChannelSet were saved as .RDS files for reproducibility.

### 2.3. Deriving estimates of epigenetic clocks and methylation-based metrics

Epigenetic ages and related methylation metrics were computed from the cleaned beta matrix using validated implementations or author-provided code. Canonical clocks (Horvath multi-tissue [Horvath, 2013]), Horvath skin-and-blood (Horvath et al., 2018), Hannum (Hannum et al., 2013), PhenoAge, GrimAge (components and composite), (A.T. Lu et al., 2019) and DNA methylation telomere length (DNAmTL; [A.T. Lu et al., 2019]) were estimated; where applicable, principal-component (PC) implementations were used to enhance reliability (Higgins-Chen et al., 2022). System-specific aging (SystemsAge) was quantified by projecting blood DNAm PC implementations onto organ-system axes (blood, brain, inflammation, heart, hormone, immune, kidney, liver, metabolic, lung, musculoskeletal), scaling predictions to years, and converting to age-adjusted residuals using reference-cohort parameters before re-centering to absolute system ages (Sehgal et al., 2025). Additional causal/stochastic and other clocks (AdaptAge, CausAge, DamAge (Ying et al., 2024); stochastic Horvath/Hannum/PhenoAge/Zhang (Tong et al., 2024); IntrinClock with adult-age back-transformation (Tomusiak et al., 2024); and RetroClock v1/v2 [Ndhlovu et al., 2024]) were generated from published coefficient tables or locked models.

For downstream analyses, epigenetic metrics were age- and covariate-adjusted using linear models that included chronological age, sex, and cell-type fractions; SystemsAge followed its published calibration procedure. All scripts were written in R (key packages: minfi, sesame, EpiDISH, ENmix, glmnet, data.table, tidyverse), and model resources (PC-clock rotations, SystemsAge coefficients) were loaded from locked RData/CSV files. Final outputs included per-participant tables of clock values, cell-type estimates and ratios, SystemsAge domains, and ancillary predictors for statistical testing.

### 2.4. Analysis of changes in PM between bezisterim and placebo

Of the DNAm samples processed, 33 met quality control criteria and were retained for downstream analysis. Promoter region methylation levels were extracted, and the mean β value across CpG sites in promoter regions was calculated for each CpG. Average differences in PM between the bezisterim group (n = 17) and placebo group (n = 16) were computed for each gene based on UCSC_RefGene_Name annotations. Statistical significance of these differences was evaluated using parametric testing, and *P* values were corrected for multiple testing by the Benjamini-Hochberg false discovery rate (FDR) procedure, with a significance threshold of FDR *P* < 0.05. Between-group differences in bezisterim and placebo for epigenetic age acceleration (EAA) were estimated by calculating the residual of the clock to age and adjusting for sex, the first three potential components, and genotype. Statistical tests used were unpaired t-test with Welch correction.

Genes were further filtered to retain only those with an absolute mean methylation difference ≥ 5% (rounded to the nearest integer) between bezisterim and placebo. These genes were designated as DPM genes and underwent subsequent downstream analyses.

### 2.5. Analysis of PM correlations with clinical measures

The sum of promoter CpG β values for each gene with complete array data for all 33 patients was analyzed for correlations with 24 clinical measures (**ST35**). Seven measures were excluded from further analysis for technical reasons. Chronological age is a measure that cannot be modified in clinical studies. Both C-reactive protein (CRP) and tumor necrosis factor (TNF) measures had single patients with outliers that drove correlations. Updated Homeostatic Model Assessment for insulin resistance (HOMA2-IR) and %S are non-linear correlates of combined fasting glucose and insulin measures. *RANTES* is an X-linked gene, and correlates were based on gender patterns. Finally, waist-hip ratio was measured only at baseline, so correlations with study completion values were not possible.

### 2.6. Genes associated with EAA in biological clocks

Genes associated with CpGs were identified for 9 clocks with significant differences in EAA, and available CpG publications for these genes were cross-referenced for DPM matches and termed “DPM clock genes.”

### 2.7. Artificial intelligence (AI) disclosure

**Supplementary File 1** contains supplementary tables 1-40 (**ST1-ST40)**. Each gene in **ST2** was searched in Chrome using AI mode separately for the following traits: “Alzheimer,” “Nominated target,” “Microglial neuroinflammation,” “Inflammation,” “Cognition,” “Diabetes,” “Obesity,” “M1 polarization,” “Glycolysis,” “Kinase cascades,” “Phosphoprotein,” “Lipids,” “Chromatin remodeling,” “Transcription factor,” and “AD hub gene.” Potentially beneficial changes in genes were identified and curated from the published data, with specific links provided in **Supplementary File 2**.

## 3. Results

### 3.1. Epigenetic clocks

We assessed the effects of bezisterim on 13 independent EAA clocks that address various processes (**Figure 1**). These clocks have different foci: PCHorvath2 and PCHannum relate to principal components of biological aging; PCGrimAge links to principal components of mortality; IntrinClock is associated with aging independent of immune factors; DamAge relates to DNAm caused by injury, disease, and mortality; Stochastic Zang, Horvath, and PhenoAge are associated with random, aging-related DNAm; retroviral integration clocks (RetroClock and RetroClockV2) are associated with epigenetic changes in long interspersed transposable elements (e.g., LINE-1); and organ-specific SystemsAge clocks are associated with epigenetic aging of the heart, lungs, and SystemsAge overall. The rate of age acceleration across all 13 clocks showed significant trends were reduced in samples from bezisterim-treated participants with AD (n = 17) versus samples from placebo-treated participants with AD (n = 16) (**Figure 1**). This finding suggests that bezisterim may modulate the progression of aging-related epigenetic changes in AD.

**Figure 1.**
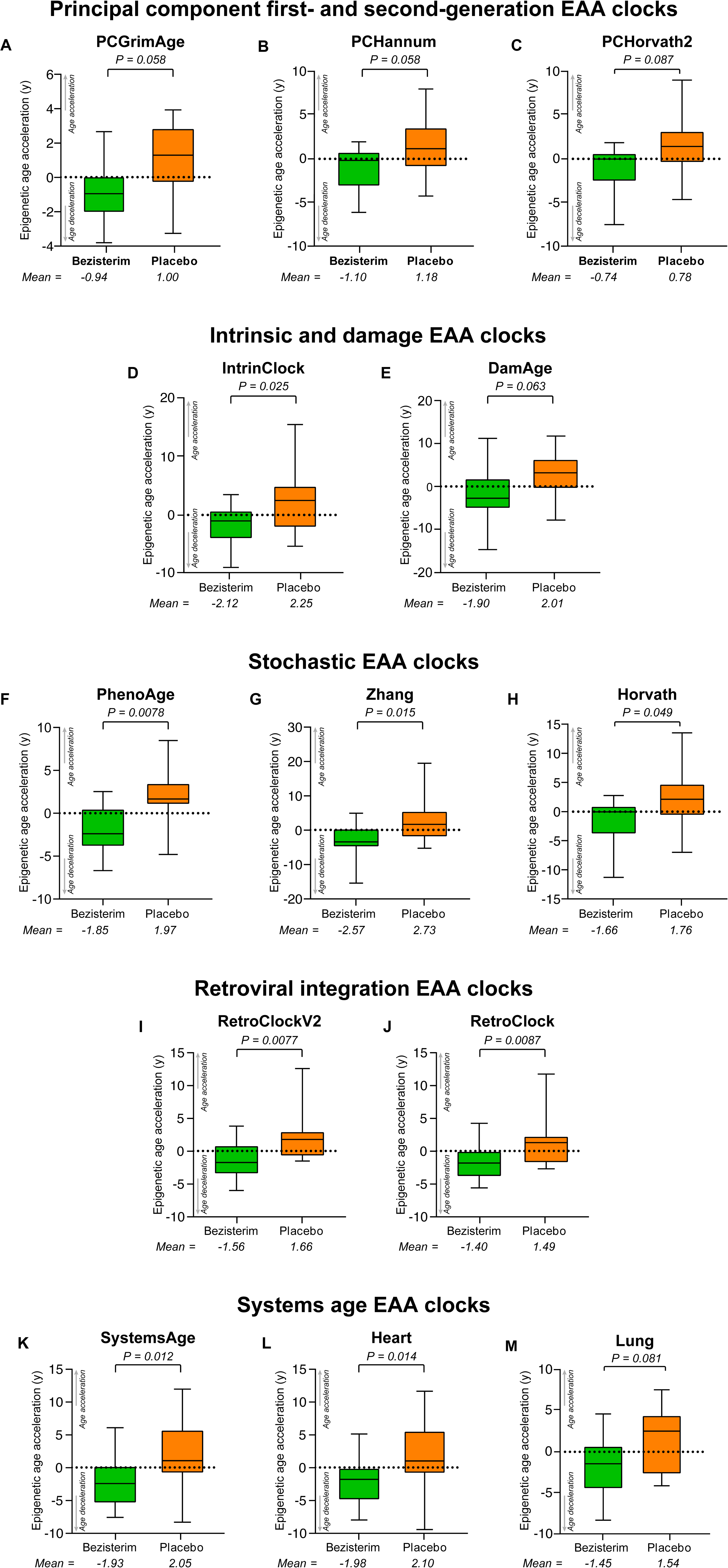
Decelerated Aging in First- and Second-Generation Aging Clocks After 30 Weeks of Treatment With Bezisterim. **(A-M)** Boxplots show the medians (solid lines) and ranges (whiskers) of EAA of DNA samples from patients with AD following 7 months of treatment with bezisterim (green, n=17) or placebo (orange, n=16), as predicted by established PC first- and second-generation clocks **(A-C)**, intrinsic and damage clocks **(D-E)**, stochastic clocks **(F-H)**, retroviral integration clocks **(I-J)**, and systems age clocks **(K-M)**. Positive and negative values surrounding the dashed reference lines indicate accelerated and decelerated aging, respectively. *P* values are by unpaired t-tests with Welch correction. AD, Alzheimer’s disease; EAA, epigenetic age acceleration; PC, principal component.

### 3.2. Gene investigations

**ST1** shows genes whose differential expressions might contribute to previously reported potential benefits of bezisterim on age deceleration, clinical assessments, and metabolic and inflammatory biomarkers. Various activity parameters were investigated, and they indicate significant differences between bezisterim and placebo.

### 3.3. Differences in DNA promoter methylation

Volcano plots (**Figure 2**) show the differences in DPM (absolute value > 5%; FDR *P* < 0.05) between treatment with bezisterim or placebo over 30 weeks for 2581 genes, using average promoter CpG methylation beta values as a surrogate for likely altered expression (**Figure 2A**). The abscissa shows bezisterim-treated genes with increased DPM as positive and decreased DPM as negative. Of 2581 bezisterim-associated DPM genes, 2559 had increased DPM and 22 had decreased DPM.

**Figure 2.**
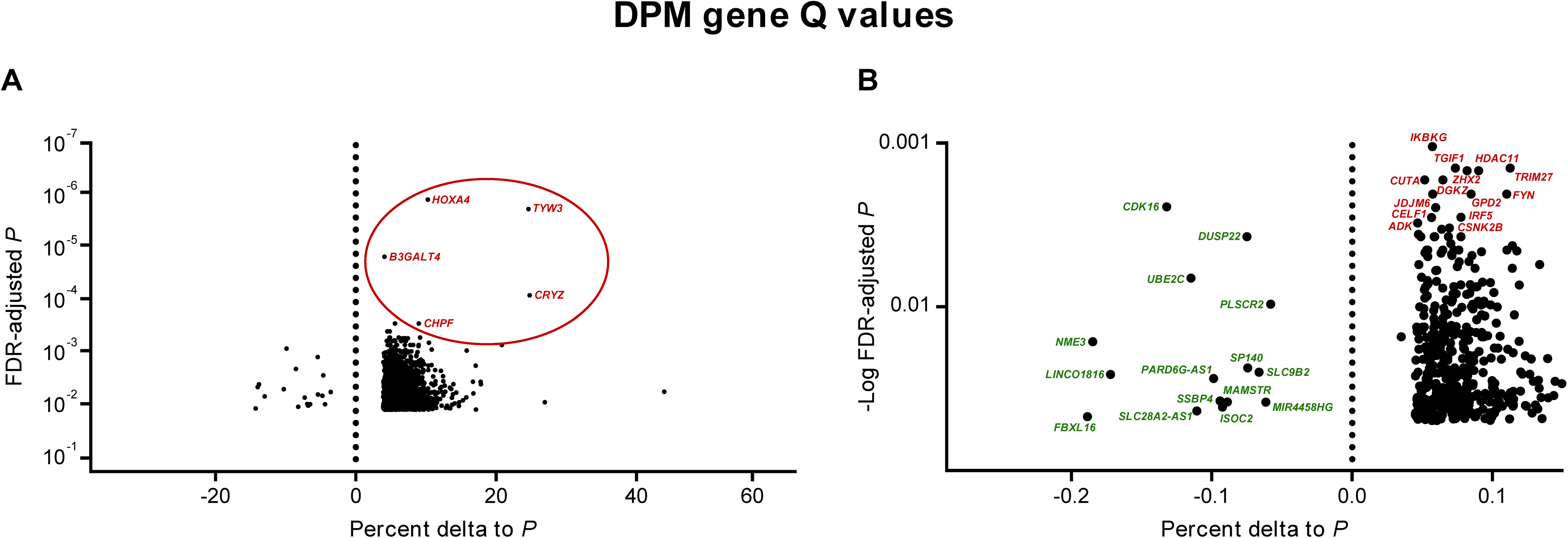
Differences in DPM Genes Following 7 Months of Treatment With Bezisterim or Placebo. **(A-B)** Treatment-dependent differences are shown for 2581 DPM genes with statistically significant FDR-adjusted *P* values below the *P* < 0.001 threshold **(A)** and 442 PBDPM genes within an adjusted threshold of *P* > 0.001 and *P* < 0.05 **(B)**. Average between-group differences in PM were computed for each of 447 genes. Positive and negative values indicate relative between-group decreases and increases in gene expression, respectively. DPM, differential promoter methylation; FDR, false discovery rate; PBDPM, potentially beneficial differential promoter methylation; PM, promoter methylation.

We identified 447 of these bezisterim-treated genes as potentially beneficial differential promoter methylation (PBDPM) genes based on inflammation and other traits associated with aging clocks and AD listed in **ST1**. Curated references supporting this designation are shown in **Supplementary File 2**. The top five PBDPM genes (FDR *P* < 0.001, circled in red in Figure 2A) had the highest increase in PM of all 2581 DPM genes.

Figure 2B shows only PBDPM genes on an expanded scale (FDP-adjusted *P* = 0.001 to 0.05) and includes the 14 genes (identified in red font) with the highest PBDPM increases, and all 15 genes with PBDPM decreases (identified in green font). Examples in **Table 1** describe activity associated with expression for the PBDPM genes. For the five identified in Figure 2A, *HOXA4* has epigenetic implications in neuropathy and cognitive decline; *TYW3/CRYZ* in insulin resistance (IR) and aberrant ubiquitination (in AD and amyotrophic lateral sclerosis); *B3GALT4* in AD is associated with dysregulated glycosylation, microglial neuroinflammation, and proinflammatory kinase cascades; and *CHPF* in prion disease, inflammation, and cognitive decline.

**Table 1.**
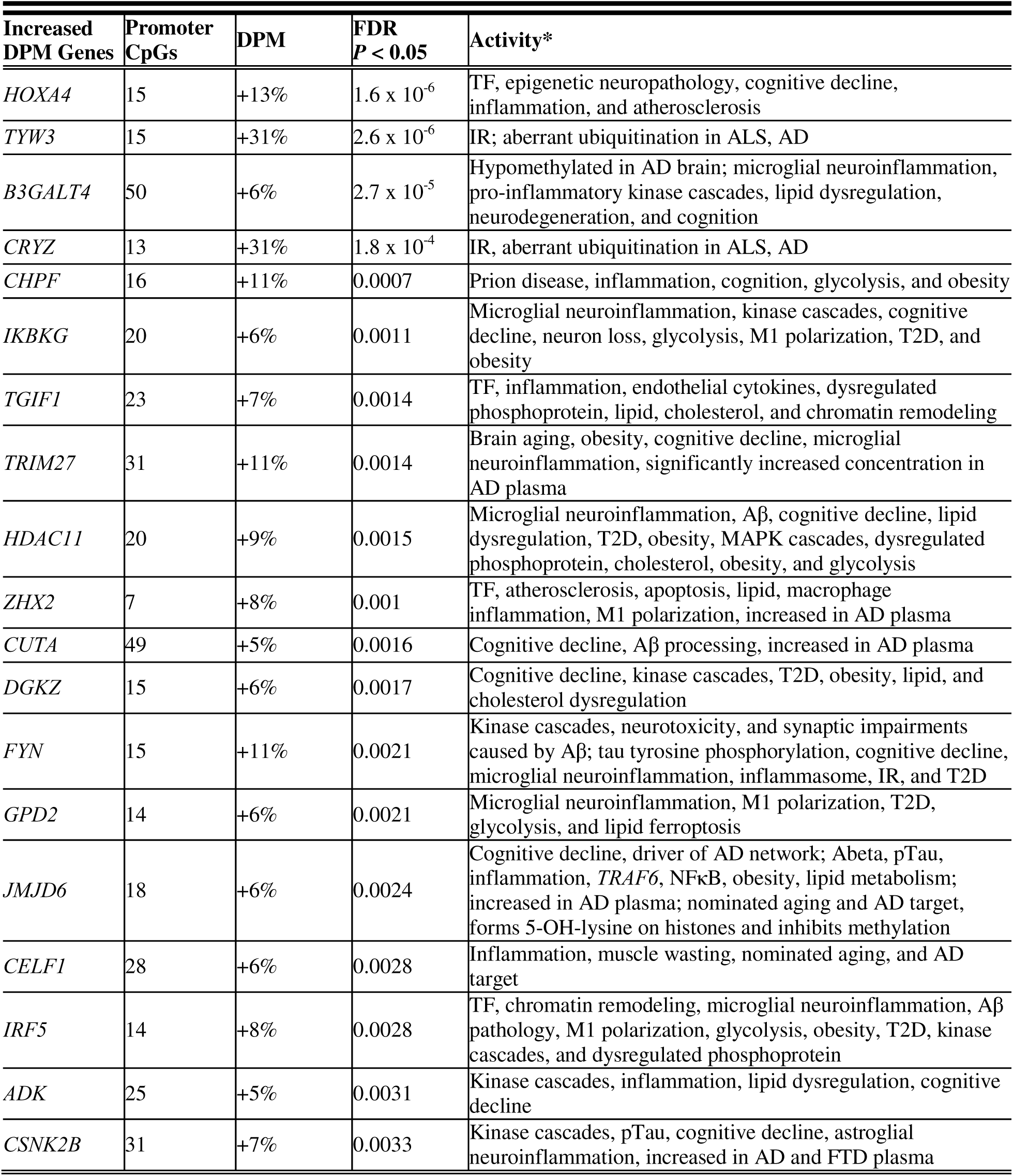
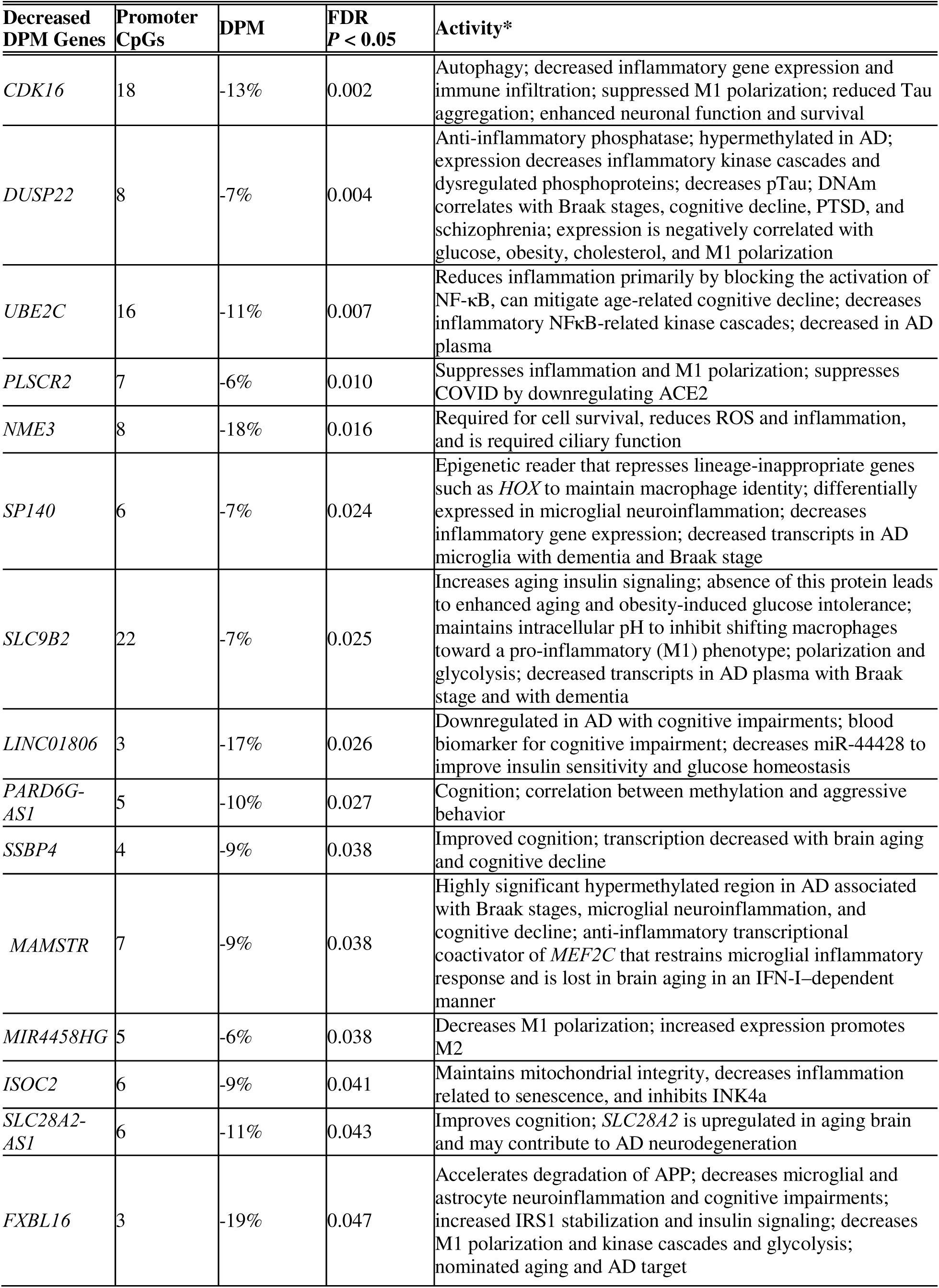
Genes With Potentially Beneficial Changes Identified in Figure 2. Potentially beneficial activities for genes with significantly increased or decreased DPM are identified and referenced, inclusive of the number of promoter CpGs identified, the DPM percentage, and the FDR-adjusted *P* value > 0.05 for each gene. *See Supplementary File 2 for gene activity Reference Catalog Aß, amyloid β; ACE2, angiotensin-converting enzyme 2; AD, Alzheimer’s disease; ALS, amyotrophic lateral sclerosis; APP, amyloid precursor protein; CpG, 5’-cytosine-phosphate-guanine-3’; DNAm, methylated DNA; DPM, differential promoter methylation; FDR, false discovery rate; FTD, frontotemporal dementia; IFN-I, interferon type 1; INK4a, inhibitors of CDK4 a; IR, insulin resistance; IRS1, insulin receptor substrate; MAPK, mitogen-activated protein kinase; NF-κB, nuclear factor kappa B; PTSD, post-traumatic stress disorder; ROS, reactive oxygen species; T2D, type 2 diabetes; TF, transcription factor.

The gene in Figure 2A with the highest increase in PBDPM, *IKBKG*, is essential for NF-κB activation, but it is also linked to microglial neuroinflammation and cognitive decline in AD.

Fifteen genes with significantly decreased PBDPM are associated with presumed increased expression (Figure 2B) based on their known functions (**Table 1**). The two genes in this group with the greatest decrease in DPM are *CDK16*, thought to enhance neuron survival through improved autophagy and decreased inflammatory gene expression, and *DUSP22*, an anti-inflammatory phosphatase that decreases kinase cascades.

Forty-five PBDPM genes were identified from nine EAA clocks with published CpGs and significant bezisterim-associated age deceleration, by identifying the promoter-associated CpGs. We found 249 (Figure 3) shared associations with AD, aging, and diseases of aging. These included cognition, diabetes, obesity, M1 polarization, kinase cascades, dysregulated lipids, glycolysis, dysregulated phosphoproteins, nominated aging and AD targets, chromatin remodeling, and TFs listed in **ST2**. Red circles in Figure 3 indicate PBDPM in genes with increased DPM and known associations to inflammation; gray circles indicate PBDPM in genes with increased DPM and no identified association to inflammation; and green circles indicate PBDPM in genes with decreased DPM. Of this latter group, 39 (87%) were associated with inflammation.

**Figure 3.**
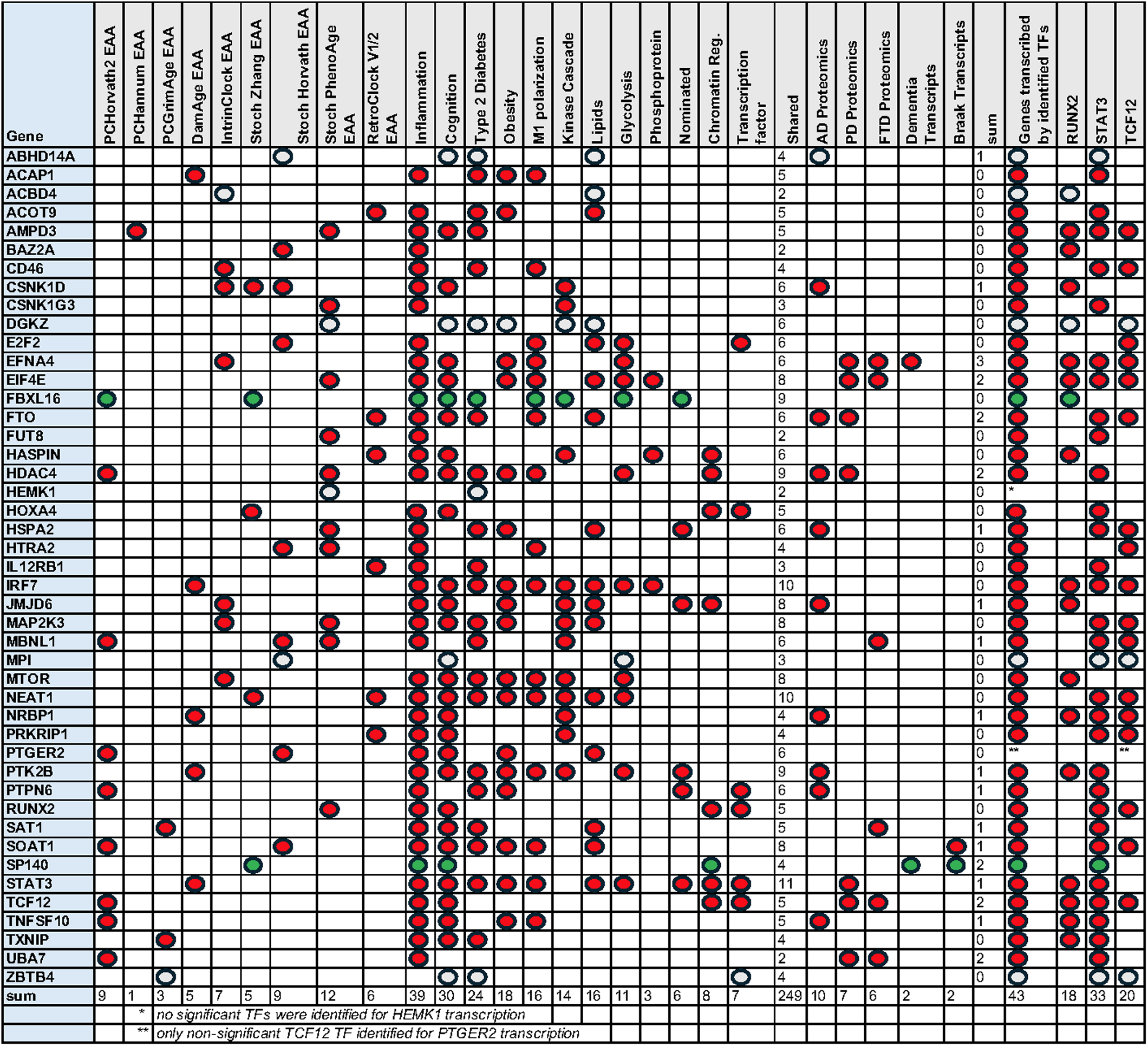
PBDPM Clock Genes. Forty-five clock PBDPM genes shared between nine EAA clocks and 12 reported aging and AD traits. In addition, expression of some of these genes were identified in AD, PD, and FTD proteomic, and in AD microglial dementia and Braak pathology score transcriptomic reports. Also included are details for PBDPM transcription factors that are known to transcribe the PBDPM genes. AD, Alzheimer’s disease; EAA, epigenetic age acceleration; FTD, frontotemporal dementia; PBDPM, potentially beneficial differential promoter methylation; PD, Parkinson’s disease; TF, transcription factor.

Bezisterim was associated with PBDPM clock genes linked to plasma proteomics in AD, Parkinson’s disease (PD), and frontotemporal dementia (FTD) (Ali et al., 2025). Also, microglial transcription levels were linked to PBDPM genes (Kosoy et al., 2025). Of 18 PBDPM genes linked to neurologic disease, higher DPM seen with bezisterim appears to translate to a dampening of those genes’ deleterious effects. In addition, bezisterim might provide beneficial effects via a decrease in expression of *EFN4A* (associated with dementia), *SOAT1* (associated with Braak pathology scores), and *SP140*. These three clock PBDPM genes have been reported to have alterations to AD microglial transcripts (Kosoy et al., 2025).

Seven clock EAA genes were identified as having increased PBDPM following bezisterim treatment associated with TFs. Three of those TFs (*RUNX2*, *STAT3*, and *TCF12*) were significantly associated with transcription of 43 of the 45 shared genes. This suggests that increases in DNAm of TF expression might compound the effects of decreased expression of target genes from bezisterim treatment.

Associations between all 447 bezisterim PBDPM genes and metabolic inflammation, aging, and AD are shown in **ST3**. These associations indicate a sharing of PBDPM genes among the risk factors in **Table 1**. Of the 447 shared genes, a total of 1670 shared risks were identified and 354 (79%) of the shared genes were associated with inflammation, . Similar to the findings in the shared clock gene matrix, shared PBDPM genes included 189 related to cognition, 156 to T2D and IR, 148 to obesity, 146 to M1 polarization, 143 to inflammatory kinase cascade genes, 120 to lipid dysregulation, 90 to glycolysis and Warburg effect genes, 76 to dysregulated phosphoproteins, 37 to nominated aging and AD risk targets, 75 to chromatin remodeling genes, and 78 to TFs. See **Supplementary File 2** for potentially beneficial activities associated with all 447 PBDPM genes.

There was evidence of PBDPM genes overlapping with neurodegenerative diseases. In AD, PD, and FTD plasma, 150 PBDPM genes had significantly increased plasma concentrations (Ali et al., 2025) but DPM was significantly increased with bezisterim; one (*UBE2C*) had significantly decreased AD and PD plasma concentrations but decreased DPM with bezisterim. Of these 151 genes, 106 were associated with potential improvements in AD plasma levels, 62 were associated with PD, and 34 were associated with FTD (**ST3, ST25, ST26, ST27**). In addition, 50 genes showed significantly increased microglial transcripts for AD dementia (30) or Braak pathology score (20) (Kosoy et al., 2025), but also significantly increased DPM (**ST3, ST28, ST29**).

The potential of TF-compounding effects on gene expression was also seen in the larger matrix. A total of 437 of the 447 PBDPM genes were significantly associated with transcription by 39 of the 78 identified PBDPM TFs. **ST2** and **ST3** provide a master table and figure denoting the gene name and DNAm information for each gene. **ST4-ST30** convey individual clock or trait sharing with other parameters. TF results for ChEA 2022 and ENCODE 2015 for all shared genes are noted in **ST31, ST32**, with shared clock genes in **ST33** and **ST34**.

### 3.4. AD hub gene and PBDPM gene sums of PM correlations with clinical measures

We investigated the relationships between the sum of PM for each of the 447 PBDPM genes for correlations with 24 clinical measures (**ST35**) and found that these include 179 known AD network hub genes (**ST36**). There were 1152 significant correlations of bezisterim-altered PM sums of PBDPM genes with clinical measures (**ST36**). We identified 426 significant correlations with AD hub genes. For placebo, there were 494 significant correlations with clinical measures and PBDPM genes, including 202 significant correlations with identified AD hub genes (**ST37**). We also found 107 significant correlations between PM sums of EAA clock–associated PBDPM genes and clinical measures (**ST38),** including 58 correlations with AD hub genes.

We examined bezisterim-associated PBDPM networks correlating to neurologic assessments (Alzheimer’s Disease Composite Score [ADCOMS], Alzheimer’s Disease Cooperative Study-Activities of Daily Living [ADL], Clinical Dementia Rating-Sum of Boxes [CDR-SB], Alzheimer’s Disease Cooperative Study-Clinical Global Impression of Change [CGIC], Alzheimer’s Disease Assessment Scale-Cognitive Subscale [Cog12], Global Statistical Test [GST], Neuropsychiatric Index [NPI]) and AD hub genes (**ST39,** Figure 4). There were 263 significant correlations of 157 PBDPM genes with 8 neurologic assessments, with a subset of 112 correlations of 62 AD hub genes. The networks for the genes correlated with neurologic assessments are shown in Figure 4, and the AD hub gene subset is shown in red bubbles.

**Figure 4.**
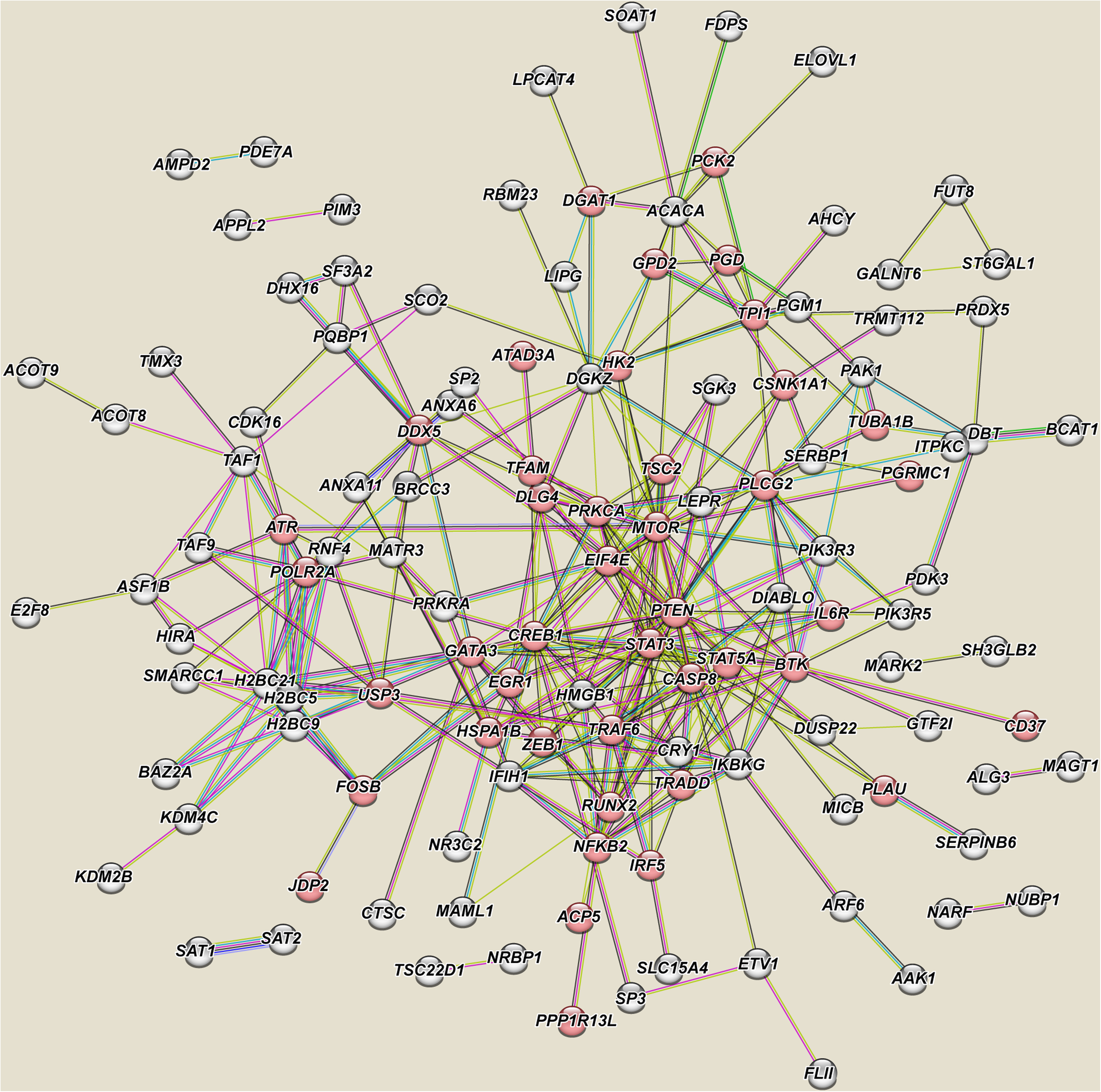
Bezisterim PBDPM Genes With Promoter Sum Correlations. Improved neurologic assessments in ADCOMS, ADL, CDR-SB, CGIC, Cog12, GST, MMSE, and NPI are associated with known AD network hub genes, identified in red. ADCOMS, Alzheimer’s Disease Composite Score; ADL, Alzheimer’s Disease Cooperative Study Activities of Daily Living Inventory; CDR-SB, Clinical Dementia Rating – Sum of Boxes; CGIC, Alzheimer’s Disease Cooperative Study Clinical Global Impression of Change; Cog12, Alzheimer’s Disease Assessment Scale–Cognitive Subscale 12; GST, Global Statistical Test; MMSE, Mini-Mental State Exam; NPI, Neuropsychiatric Index

Based on these correlations, we investigated the PBDPM clock genes’ associations with neurological measures. Clock-associated CpGs were available for nine EAA clocks identified in Figure 3 and **ST3**. **Table 2** identifies significant correlations of six neurologic assessment z scores with PM with the nine PBDPM EAA clock genes, among bezisterim-treated patients compared to placebo. Seven are identified AD hub genes. Four individual clock genes (*DGKZ*, *NRBP1*, *PTGER2*, and *SOAT1*) were associated with two to five individual neurologic assessment z scores. For each of the genes, there were significant increases in PM, significant correlations of PM sums with neurologic assessments consistent with improvements and known associations of the individual gene with cognitive decline.

**Table 2.**
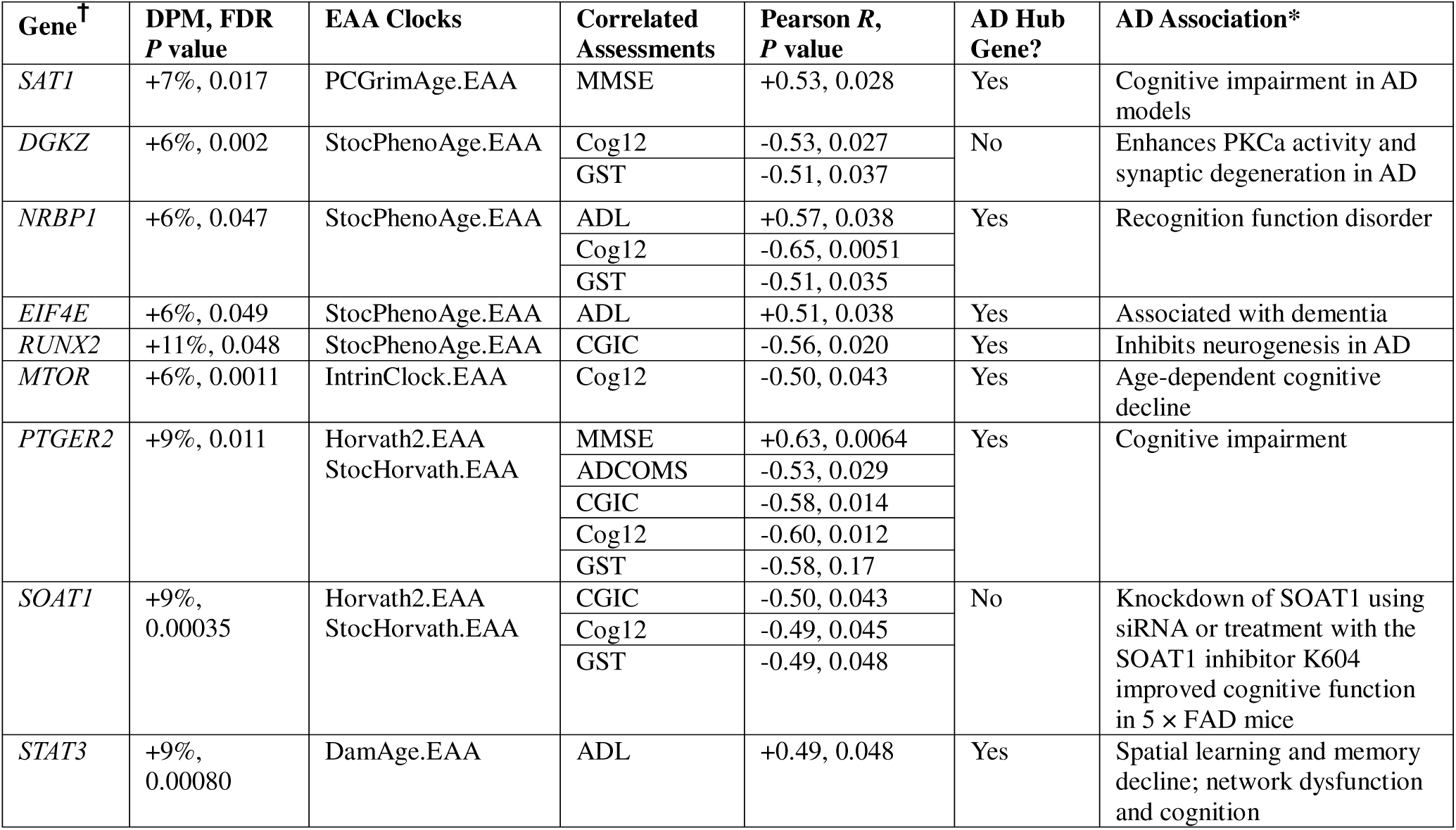
Clinical Neurologic and Cognition EEA Clock Genes. Higher bezisterim PM correlates to improvements in six neurologic assessment z scores. *See Supplementary File 2 for gene activity Reference Catalog ^†^Expanded gene names are available in ST3 AD, Alzheimer’s disease; ADCOMS, Alzheimer’s Disease Composite Score; ADL, Alzheimer’s Disease Cooperative Study-Activities of Daily Living Inventory; CGIC, Alzheimer’s Disease Cooperative Study-Clinical Global Impression of Change; Cog12, Alzheimer’s Disease Assessment Scale-Cognitive Subscale; DPM, differential promoter methylation; EAA, epigenetic age acceleration; FDR, false discovery rate; GST, Global Statistical Test; MMSE, Mini-Mental State Exam; PKCa, protein kinase C alpha; PM, promoter methylation; siRNA, small interfering RNA.

We further assessed the significance and the distributions of positive and negative correlations of genes versus clinical measures for bezisterim and placebo patients (**ST40**). There were significant (Fisher Exact *P* < 0.05) treatment-associated differences in distributions of correlations for 17 clinical measures with the 2581 DPM genes, and 11 of these were significant among the 447 PBDPM genes. The correlation direction with bezisterim subjects’ clinical measures were predominantly associated with potential improvements. The correlations with placebo subjects’ clinical measures were predominantly associated with potential decline.

Examples of PBDPM AD hub gene correlations with clinical measures are provided in Figure 5, with blue arrows indicating the direction of improvement and red arrows showing association with AD progression.

**Figure 5.**
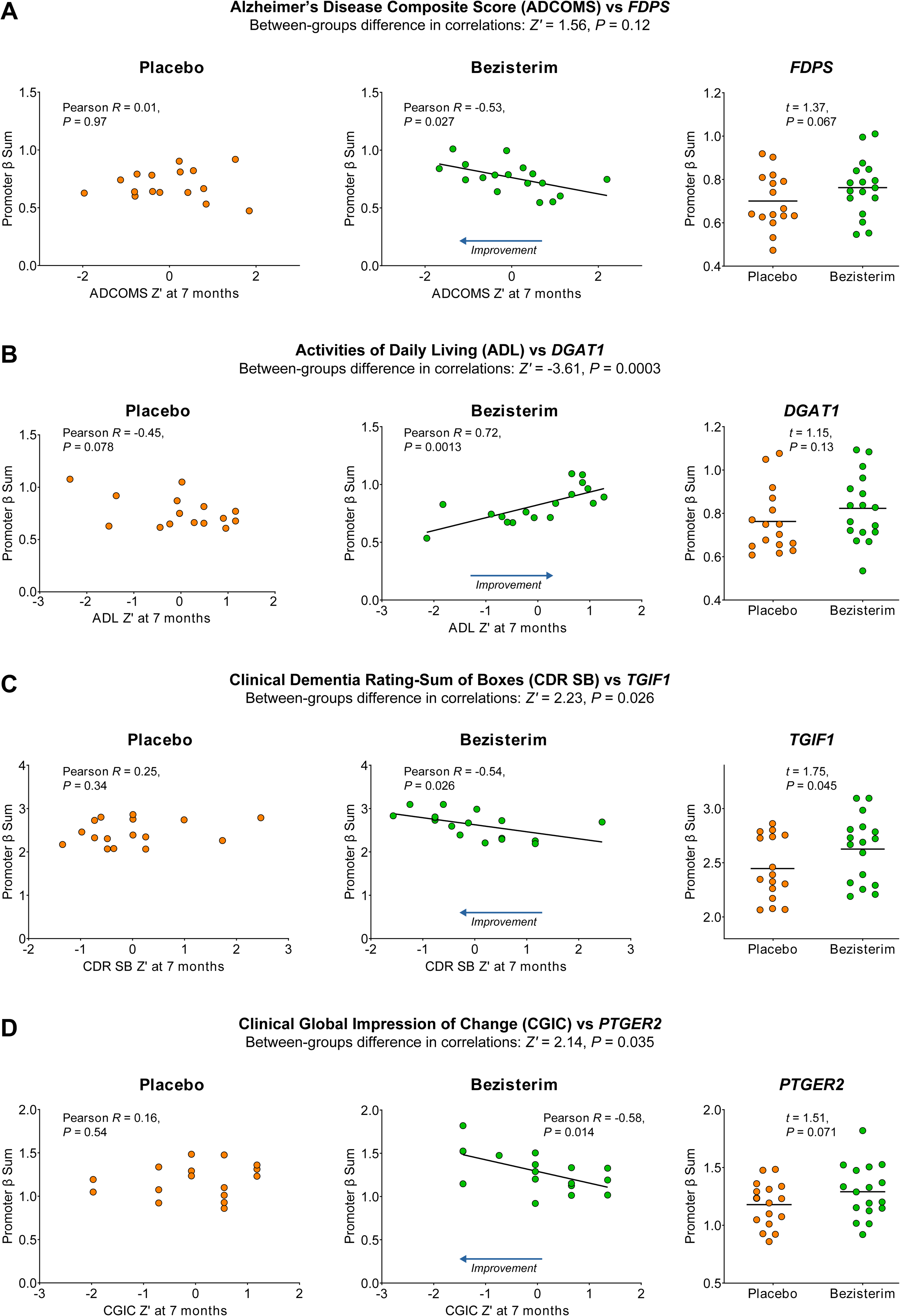
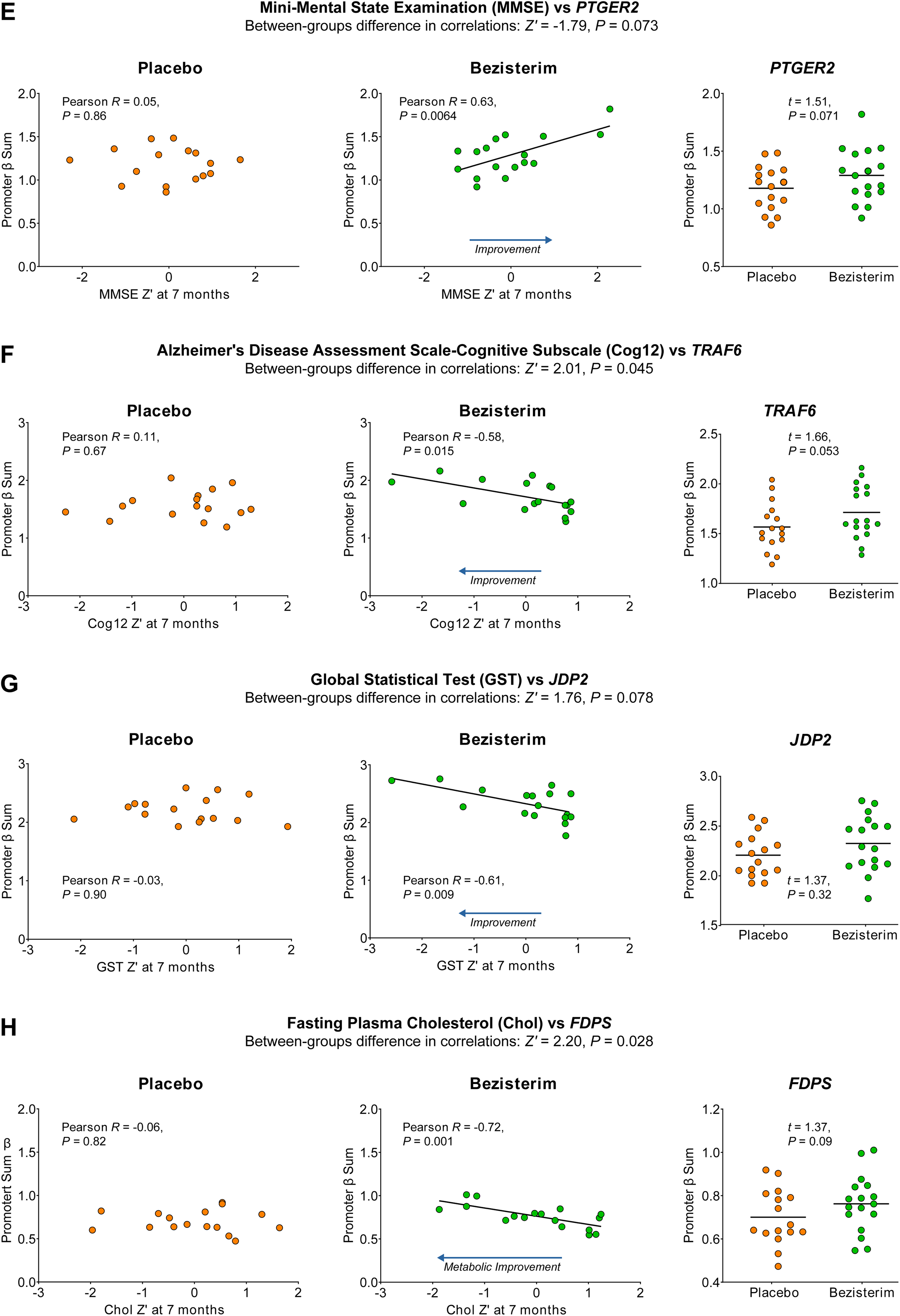
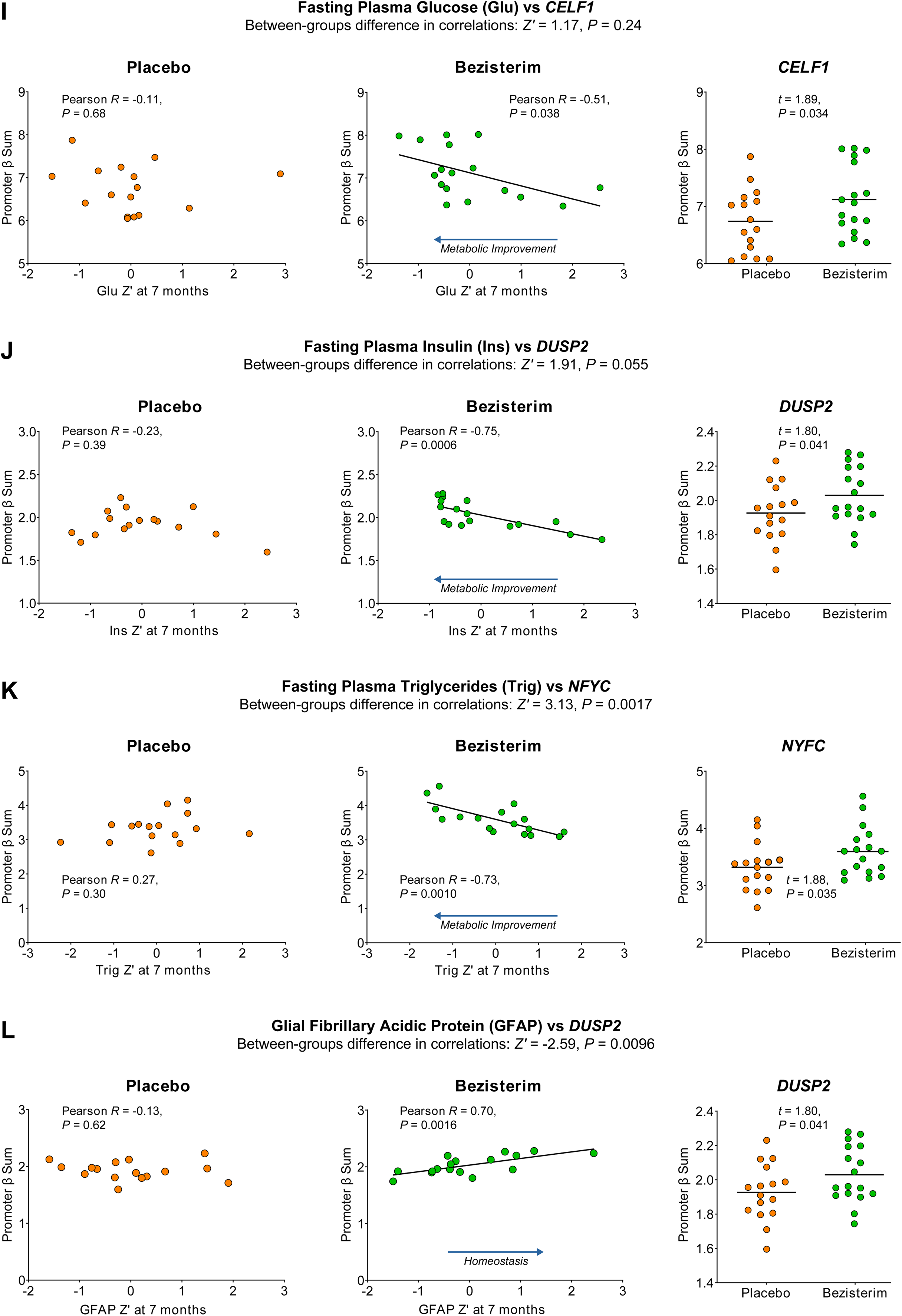
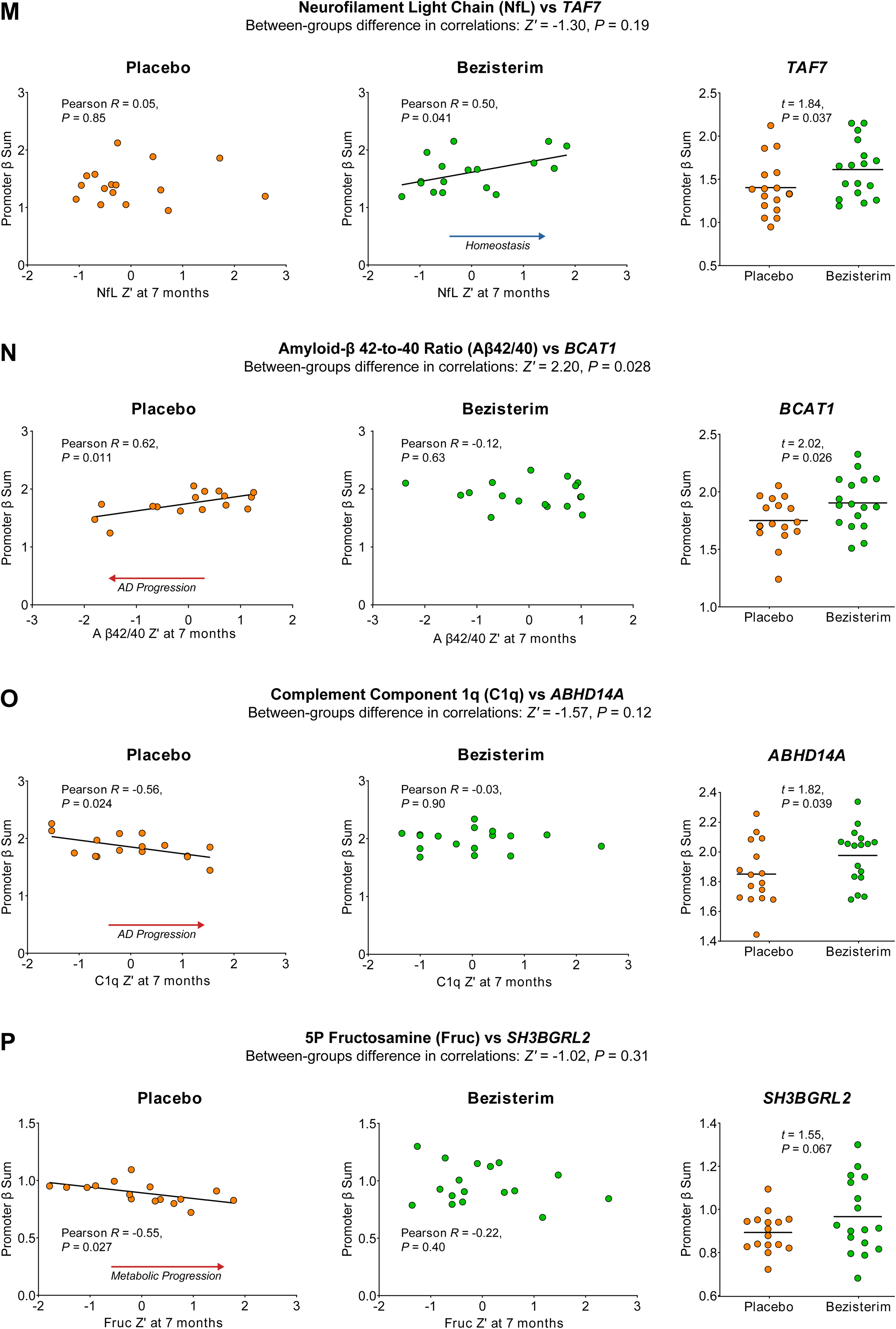
Correlations Between Genes and Clinical Measures. **(A-P)** Examples of significant correlations between clinical measures and PBDPM AD hub genes among AD patients treated with bezisterim or placebo. Pictured are graphs of placebo and bezisterim patient clinical measure z scores versus gene PM (β) sum Pearson correlations (*R* and *P*), and distribution differences of PM sums (t-test *P* values). Also included are z score *P* values for the significance of correlation differences. Differences in the sum of bezisterim PM compared to placebo are complementary to previously reported directional improvements in neurological assessments. Arrows indicate context for clinical measures. See the text for details. Aβ42/40, amyloid-β 42/40 ratio; AD, Alzheimer’s disease; ADCOMS, Alzheimer’s Disease Composite Score; ADL, Alzheimer’s Disease Cooperative Study Activities of Daily Living Inventory; C1q, component 1q; CDR-SB, Clinical Dementia Rating – Sum of Boxes; CGIC, Alzheimer’s Disease Cooperative Study Clinical Global Impression of Change; chol, fasting plasma cholesterol; Cog12, Alzheimer’s Disease Assessment Scale–Cognitive Subscale 12; fruc, 5P fructosamine; GFAP, glial fibrillary acidic protein; glu, fasting plasma glucose; GST, Global Statistical Test; ins, fasting plasma insulin; MMSE, Mini-Mental State Exam; NfL, neurofilament light chain; PBDPM, potentially beneficial differential promoter methylation; PM, promoter methylation; trig, fasting plasma triglycerides.

In addition to the nine clock genes with significant PBDPM correlated to neurologic assessments, improvements included associations with ADCOMS (8), ADL (89), CDR-SB (5), CGIC (36), cog12 (51), GST (41), Mini-Mental State Exam, or MMSE (17), and NPI (1) among the 447 PBDPM genes (**ST40**). Examples of seven of these are shown in Figures 5 **A to G**.

For bezisterim versus placebo, we found positive correlations of neurologic assessments with AD hub gene PM sums (absent in placebo: ADL with *DGAT1*, MMSE with *PTGER2*; Figures 5B **and 5E**), and negative correlations (absent in placebo: ADCOMS with *FDPS*, CDR-SB with *TGIF1,* CGIC with *PTGER2,* Cog12 with *TRAF6,* and GST with *JDP2*; Figures 5B**, 5C, 5D, 5F, and 5G**). Correlations were associated with significant directional improvements (z score *P* < 0.05) for ADL, CDR-SB, CGIC, and Cog12, and trends (*P* < 0.10) for MMSE and GST.

There were also significant positive correlations of metabolic measures and AD hub gene PM (cholesterol with *FDPS,* fasting plasma glucose with *CELF1,* insulin with *DUSP2,* and fasting triglycerides with *NYFC;* Figures 5I **to 5K**). These correlations were associated with significant improvements in the metabolic parameters.

Two clinical measures showed significant positive correlations for AD hub genes with AD biomarkers. Higher bezisterim PM of *DUSP2* was associated with higher glial fibrillary acidic protein (GFAP) levels (Figure 5L), and PM of *TAF7* was associated with higher neurofilament light chain (NfL) levels (Figure 5M). There were no significant differences in GFAP or NfL z scores between bezisterim and placebo, and potential interpretations of these correlations are addressed in the discussion. Further, there was a significant positive correlation with *BCAT1* and Aβ42/40 ratio (Figure 5N) and negative correlations of AD hub genes’ PM for *ABHD14A* and *SH3BGR2L* with complement component 1q (C1q) and fructosamine, respectively (Figures 5O **and 5P**). These correlations were associated with increased AD inflammatory and metabolic risk.

Figure 6 shows three examples of 270 PBDPM gene associations (including 110 AD hub genes) with significantly lower systolic blood pressure (SBP) among placebo subjects. Compared to bezisterim subjects, numerically lower placebo PM of *ABCA2* (Figure 6A) and *TUBA4A* (Figure 6B) and significantly lower placebo PM of *CTSL* (Figure 6C) were associated with lower SBP. The correlations by treatment were significant for all three, and potential interpretations of these correlations are addressed in the discussion.

**Figure 6.**
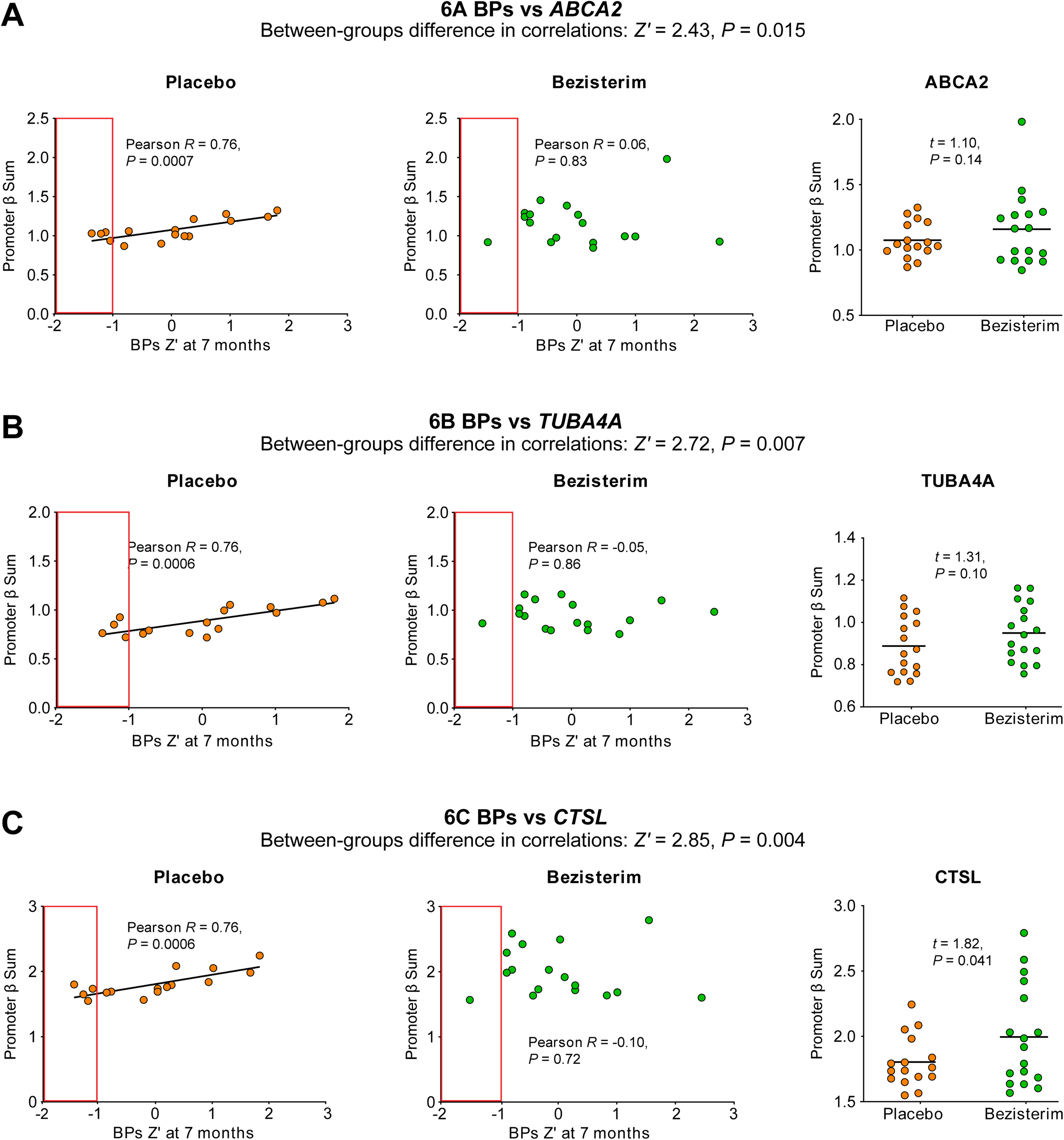
Correlations Between Genes and SBP. Examples of significant correlations between clinical measures and PBDPM AD hub genes among AD patients treated with bezisterim or placebo. Graphs of placebo- and bezisterim-treated patient SBP (termed BPs in the figures) z scores versus gene PM (β) sum Pearson correlations (R and P), distribution differences of PM sums (t-test *P* values), and distributions. Also included are z-score *P* values for the significance of correlation differences. Distributions of placebo- and bezisterim-treated patient BPs z scores include red boxes highlighting differences in lower SBP for placebo. See the text for details. AD, Alzheimer’s disease; PBDPM, potentially beneficial differential promoter methylation; PM, promoter methylation; SBP, systolic blood pressure.

There were significant correlations for individual clinical measures with the bezisterim-associated PM sums of genes known to be involved in dysregulation of the same clinical measure (citations listed in **Supplementary File 2**). For instance, *BTK* PM correlated with MMSE, and *NFKB2* PM with ADL related to cognitive function (Ekpenyong-Akiba et al., 2020); *BHLHE40* and *NFE2L2* PM with cholesterol (microglia, foam cells, and atherosclerosis), and *FYN* and *RAC1* PM with triglyceride levels.

By contrast, PM with placebo was significantly correlated with genes involved in dysregulation of associated clinical measures. For example, *BCAT1* PM correlated with Ab42/40 ratio (Aβ load) (Harris et al., 2020); *MAP2K3* PM with C1q (pTau, synapse loss) (Dejanovic et al., 2018); *RLIM* and *PGRMC1* PM with fructosamine (glycemic control); and *PTGER2* and *OCIAD1* PM with SBC (cerebral hypometabolism due to associated mitochondrial impairment).

## 4. Discussion

### 4.1. Epigenetic clocks

The finding that the rate of age acceleration across all 13 clocks showed significant trends for reduction in samples from bezisterim-treated AD participants versus samples from placebo-treated AD participants suggests that bezisterim can modulate the methylation changes predictive of biological aging. Emerging studies indicate that accelerated biological aging may not only serve as a biomarker for AD risk and progression (Li et al., 2021) but could also contribute to the pathogenesis of AD. Two second-generation epigenetic clocks, DNAmPhenoAge and DNAmGrimAge, predicted progression from cognitively normal aging to mild cognitive impairment or AD and worse longitudinal cognitive outcomes (Bonham et al, 2025). Epigenetic age was also strongly associated with cortical thinning in AD-relevant regions and white matter disease burden. Understanding how genes, pathways, or networks with differentially methylated CpGs influence the progression of AD is complex, since methylation within a specific gene can either enhance or reduce its expression. Since PM usually inversely correlates with gene expression, analyzing gene level aggregate PM can provide insight into potential transcriptional impact.

The genes associated with significant bezisterim-associated age deceleration clock CpGs were identified across 13 aging clocks (Figure 1) and cross-referenced with bezisterim-associated PBDPM genes that had significant average CpG methylation differences based on multiple PM sites. This DPM analysis may provide a higher probability of ascertaining the activity of various genes relevant to regulation of expression.

We identified 45 EAA clock genes with significant PBDPM from 9 clocks with available CpG identification. For the significant age deceleration of the PCHorvath2 EAA clock (Figure 1), there were nine PBDPM genes identified and, interestingly, all were associated with inflammation (Figure 3). Furthermore, there were FDR-adjusted *P* < 0.05 significant Pearson correlations between 40 of the 45 bezisterim PBDPM EAA clock genes (**ST38**), with 14 of the 24 clinical measures suggesting that bezisterim may be modulating the epigenetic regulation of genes predictive of biological aging that contribute to age-related neurodegeneration. There were similar patterns observed overall with the 447 PBDPM genes (**ST2)** that shared associations with inflammatory, metabolic, and transcriptional dysregulation.

### 4.3. Differences in promoter methylation

Of 11,322 genes, 2581 (23%) were found to have FDR-adjusted *P* < 0.05 significant DPM. Potentially beneficial effects of DPM were identified in 447 genes, including 432 with increased DPM and 15 with decreased DPM (Figure 2). The significance of these findings supports a bona fide activity of bezisterim to enrich PBDPM of AD patients’ genes in a potentially beneficial manner.

The data from Figure 3 and **Table S2** suggest broad functional associations between the DPM of specific genes and AD/aging risks, potentially identifying molecular changes associated with effects of bezisterim-associated systems biology. For instance, Aβ and pTau DAMPs activate kinases that inhibit insulin signaling (Berlanga-Acosta et al., 2020; Shaughness, Acs, Brabazon, Hockenbury, & Byrnes, 2020; Q. Zhang et al., 2018). Inflammation risk was the major shared trait (**Table S13**, 355 PBDPM genes) and may represent the teleologic basis for the associations. Inflammatory responses to pathogens can eliminate the pathogen, terminate the response, and then resolve and repair damage associated with the process. Age-associated neurologic diseases such as AD and PD create DAMPs (e.g., glycosylated proteins, prion-like protein aggregates) that trigger metabolic inflammation, but resolution of the process is lacking since inflammatory responses increase, rather than eliminate, the DAMPs. Based on the PBDPM associations evident with genes associated with decreased M1 microglial and macrophage responses, and based on the requisite Warburg glycolysis effect, we can hypothesize that bezisterim may influence homeostatic restoration in the face of metabolic and protein-aggregating inflammatory responses. Bezisterim decreased macrophage inflammatory responses, kinase cascades, and insulin resistance in diabetes models (M. Lu et al., 2010; T. Wang et al., 2010) and in clinical studies of impaired glucose tolerance and T2D (C. L. Reading, Stickney, et al., 2013) (C. L. Reading, Flores-Riveros et al., 2013). In AD, bezisterim decreased glucose, cholesterol, and monocyte chemoattractant protein 1, and increased HOMA2-IR and beta cell function with alterations in the monocyte methylome (C. Reading, J. Yan; Schmunk, L.; Martin-Herranz, D.; Ahlem, C.; Markham, P.; O’Quinn, S.; Palumbo, J., 2025). Data in Figures 3 and **4** and **Table 1** that address inflammation, kinase cascades, M1 polarization, and glycolysis suggest that bezisterim may alter the polarization of macrophages and microglia, inducing a shift from an M1 pro-inflammatory state to an M2 restorative state with altered monocyte methylome in AD patients (C. Reading, J Yan; Schmunk, L.; Martin-Herranz, D.; Ahlem, C.; Markham, P.; O’Quinn, S.; Palumbo, J., 2025).

After inflammation, the second-most prevalent gene sharing was with cognition risks. There were 189 out of 447 PBDPM genes associated with cognition (**ST14**). Our findings support the proposition that bezisterim may be altering the expression of genes impacting AD inflammation, cognition, and dysregulated lipid metabolism. In this study, there were potentially beneficial changes of approximately 8% overall in PM. This percentage may be an underestimation of the overall effect for five reasons: First, kinase cascades involved in signal transduction involve sequential phosphorylation of several kinases and targets (NF-κB, MAPK, p28, and JNK) followed by phosphoprotein targets, including TFs that may lead to compounding effects on the targets and TFs. Second, as shown in Figures 3 and **ST24**, TFs with PBDPM are involved in the transcription of genes with PBDPM, again leading to compounding effects. Third, chromatin-remodeling genes with PBDPM alter access to the TFs with multiple PBDPM. Fourth, TFs work in concert, suggesting that for multiple PBDPM gene TFs with potentially decreased expression, there may be a compounding effect on transcription at a promoter with multiple TF sites, with further impact if the site is for a PBDPM gene. Fifth, the inter-connections of neurologic PBDPM and AD network hub genes in Figure 4 indicate that the changes in PM of genes correlated with neurologic assessments are likely amplified in the pathways downstream of the AD hub genes. The modest differences in examples of Hub gene PM between bezisterim and placebo in Figures 5 and **6** may occur in more than 2500 AD hub genes and downstream risk genes, leading to potential compounding amplifications of expression effects.

Mechanisms of inflammatory processes in AD and aging may be elucidated by defining biomarkers of response to bezisterim. Since there were potentially beneficial responses in the AD study (C. Reading et al., 2025), we have focused on PBDPM genes. In the absence of clinically reported serious adverse events to date, we have not explored potentially deleterious DPM, and the definition would be tenuous since the context following bezisterim treatment is clearly different than that of placebo. In fact, the modulation of DPM may be involved in the safety profile, since an 8% change in DPM has limited impact compared to complete inhibition, mutation, or knockout of a gene.

Data for PBDPM genes (**ST2)** are similar to those for PBDPM clock genes (Figure 3). Patterns show sharing of associations with inflammatory, metabolic, and transcriptional dysregulation. For available plasma levels of PBDPM clock gene products, 10 of 45 gene products were reported to be significantly increased in AD plasma, but had higher DPM with bezisterim, which is likely to have decreased expression in our study. For the overall shared genes, 105 of 433 PBDPM genes with increased methylation have been reported to have significantly increased concentrations in plasma, and one product of the 15 PBDPM genes with decreased methylation was reported to be significantly decreased in plasma.

### 4.4. AD hub gene and PBDPM gene sum of PM correlations with clinical measures

The sum of CpG PM for each of the PBDPM genes was analyzed for correlations with clinical measure z scores from the previous clinical study (C. Reading et al., 2025). With bezisterim, we found evidence for 1152 significant associations of PBDPM genes that correlated with clinical neurologic assessments, biomarkers, and inflammatory and metabolic measures. For placebo, there were significantly fewer relationships correlated to clinical measures (494), compared with bezisterim.

While bezisterim correlates might assist in characterization of mechanism, placebo correlates may help to define disease progression pathways. This suggests that correlations between clinical measures and gene promoter expression may provide valuable prognostic information in a clinical setting and act as biomarkers of clinical response.

For neurologic assessments, PM sums of 157 genes showed significant correlations of bezisterim with directional improvements in the neurologic measures of ADCOMS, ADL, CDR-SB, CGIC, MMSE, and GST that were absent in placebo. Exploration of differential PM of EAA clock genes identified nine genes with significant correlations of PBDPM associated with the same neurologic assessments.

Mapping specific clinical measure effects of correlated genes may be complex considering the compounding effect of changes in methylation of many AD hub genes, which can affect expression of multiple downstream genes. However, we were able to identify potential effects of PM of individual genes with effects on their associated clinical measure for neurologic assessments, cholesterol, triglycerides, and GFAP for bezisterim patients, and for Aβ42/40 ratio, C1q, fructosamine, and systolic blood pressure for placebo.

For bezisterim patients, correlations with GFAP and NfL may be examples of context-dependent regulation. Both are biomarkers associated with AD progression, but they both have important homeostatic functions. There were no significant treatment differences in the plasma levels of GFAP or NfL, only potentially beneficial differences in the PM of correlated genes (172 for GFAP, 5 for NfL). GFAP is crucial for blood-brain barrier maintenance and neurotransmitter balance and is upregulated after brain injury, where it helps in wound healing and promoting repair (Tiwari, 2025). GFAP neural progenitors give rise to immature neurons via early intermediate progenitors that express both glial fibrillary acidic protein and neuronal markers in the adult hippocampus (Liu et al., 2010). Plasma GFAP z scores were also correlated with higher *MAP2K1* PM and *NFE2L2* PM. *MAP2K1* induces reactive (A1) astrocytes, suggesting that bezisterim might be involved in a restorative A1-to-A2 shift. *NFE2L2* decreases neurogenesis, and a reduced expression might be associated with GFAP regulation in neural stem cells and progenitors in adult hippocampus and subventricular brain regions.

NfL is a key structural protein in axons and acts as neuronal internal scaffolding that controls release and function in synapses, influencing neurotransmission (as with NMDA receptors) and reflecting neuronal health (Yuan, Rao, Veeranna, & Nixon, 2017). NfL protein expression is closely linked to the continued growth of normal axons and the regeneration of impaired axons, with some studies suggesting an increase in NfL content and synthesis during the developmental and regenerative growth phases (H. Wang et al., 2012). Our results are compatible with the hypothesis that for placebo, AD hub genes contribute to GFAP- and NfL-associated neurodegeneration over time, and bezisterim treatment may alter network interactions to improve clinical measures.

SBP correlations are also context-dependent. Higher SBP defines hypertension, but there were no differences in hypertension between bezisterim and placebo. Lower SBP in patients with dementia (Selbaek et al., 2022) is associated with cerebral hypometabolism (Cheng et al., 2025). For placebo in this study, lower SBP was associated with lower PM of 270 genes. We previously reported imaging data from a 13-week open-label study of 23 patients with mild cognitive impairment and mild AD (Haroon et al., 2024). Bezisterim was associated with clinician-rated improvements in relative cerebral blood flow (rCBF), using arterial spin label imaging and blood-oxygen–dependent functional connectivity within the brain. rCBF is strongly correlated with fluorodeoxyglucose-positron emission tomography (FDG-PET) brain imaging and metabolic coupling in AD. The significant positive placebo correlations of SBP (Figure 6) suggest that in AD the correlated genes may contribute to decreased central nervous system (CNS) perfusion, and lower methylation of *ABCA2*, *TUBA4A*, *CTSL*, and other genes (e.g., *PTGER2* and *OCIAD1*) may result in lower SBP and CNS perfusion. The higher PM among bezisterim subjects suggest that bezisterim might improve rCBF and brain hypometabolism compared to placebo by altering the expression of genes that are covariates of perfusion. Improvements in rCBF may be associated with improvements in brain glucose and energy utilization.

The relationships with clinical measures were strengthened by the identification of predicted expression differences in genes that have direct associations to correlated clinical measures for bezisterim (*BTK* for MMSE, *NFKB2* for ADL, *BHLHE40* and *NFE2L2* for cholesterol, *IKBKB* and *MAP2K6* for insulin, *FYN* and *RAC1* for triglycerides, and *MAP2K1* and *NFE2L2* for GFAP) and for placebo (*BCAT1* for Ab42/40, *MAP2K3* for C1q, *PGRMC1* and *RLIM* for fructosamine, and *PTGER2* and *OCIAD1* for SBC) (Haroon et al., 2024).

## 5. Conclusions

Based on the impact of bezisterim on EAA clocks and evidence of correlations between potential gene expression differences and clinical responses for neurologic and metabolic measures, it is tempting to speculate that bezisterim may increase healthspan. The confirmation of the exciting exploratory findings from this analysis in a larger randomized, placebo-controlled study might portend a new direction in AD therapeutics where reversing the methylation changes predictive of biological aging could not only increase longevity but also impact the progression of age-related diseases, including AD.

Aggregated promoter-wide methylation analysis used in this study may be beneficial when exploring conditions or treatments for healthspan or diseases of aging. While clocks based on differences in individual CpG methylation have been an immense advance in understanding aging, interpretations are limited without knowing the relationship of the individual CpGs to systems biology. Known potential markers of clinical benefit in bezisterim-treated patients were associated with modulation of promoter-wide methylation of 447 age- and disease-related genes. Most of these modulations were associated with potentially beneficial regulation of inflammatory pathways. Although potentially beneficial differences in PM of AD hub genes compared to placebo are exciting examples of bezisterim effects, the gestalt of the impact of all identified epigenetic differences between bezisterim and placebo underscores the need for a systems biology approach to understanding bezisterim activity. Bezisterim increased PM of AD hub genes that are associated with clinical measures. Other therapies that alter AD hub gene methylation or expression, such as exercise, are also correlated with clinical improvement in AD (Hill & Gammie, 2022; Liu et al., 2024; Zhou, 2025).

Bezisterim appears to decrease epigenetic age acceleration by potentially biasing gene regulation towards an anti-inflammatory state, with potential improvements that may translate to clinical benefit. The specifics of potential biomarkers that might be derived from this study await replication in a larger AD population. We are currently collecting DNAm data for participants in randomized, placebo-controlled trials evaluating bezisterim in PD (NCT06757010) and long COVID (NCT06847191), with the possibility of understanding the generalizability of these findings in other neuroinflammatory/neurodegenerative settings.

## Acknowledgements

Editorial support was provided by *p*-value communications and funded by BioVie Inc.

## Ethics Statement

The studies from which the DNAm data were obtained (NCT04669028; https://clinicaltrials.gov/study/NCT04669028?term=NCT04669028&rank=1) involved humans and received IRB approval from ADVARRA (MOD01862543). The studies were conducted in accordance with the local legislation and institutional requirements. The participants provided their written informed consent to participate in this study.

## Data Availability Statement

The data supporting the findings of this study are available in the main text and supplementary materials of the article.

## Author Contribution Statement

V.B.D., C.R., and J.Y. designed the study, performed analysis, and maintained quality control of data. All authors contributed to the draft and review of all components of the paper.

