## Supplemental File 2 for "Bezisterim-associated anti-inflammatory epigenetic modulation of age acceleration and Alzheimer’s disease genes"

**Reference Catalog**

| **General Topic** | **Resource (DOI or Website Link)** |
| --- | --- |
| Aging and Alzheimer’s Braak* Pathology Score* | EWAS Catalog β, 2024. [https://ewascatalog.org/?trait=Braak*%20stage](https://protect.checkpoint.com/v2/r01/___https://ewascatalog.org/?trait=braak%20stage___.YzJ1OmJpb3ZpZTE6YzpvOjljNzMyZTM5MjY4ZDg1MDA1MDcyODAxYjc4YTlmZjRhOjc6ZWI3MjpkODAyMjUyOGQ2ODczZDEzYTZkZjBlNWEwNjlkZGEzZDFmYmRhMjNlNTU1NmM0NmY3NjRiZjA0ZmRiNDllNDFhOnA6RjpG)  Smith RG et al, 2021. doi: 10.1038/s41467-021-23243-4  Zhang L et al, 2021. doi: 10.1186/s40478-021-01177-8 |
| Human Kinases | KinHub β, 2017. [http://www.kinhub.org/kinases.html](https://protect.checkpoint.com/v2/r01/___http://www.kinhub.org/kinases.html___.YzJ1OmJpb3ZpZTE6YzpvOjljNzMyZTM5MjY4ZDg1MDA1MDcyODAxYjc4YTlmZjRhOjc6MjZhNDo3NDQ5NWExNzcyNjk4YmZlN2Y5NzZhZTA1YTYxMzc1M2FiNTQ0ODdlMzZhNTU2MGYwMjRjZjYxM2M5MGVjMDE5OnA6RjpG)  [http://kinase.com/human/kinome/](https://protect.checkpoint.com/v2/r01/___http://kinase.com/human/kinome/___.YzJ1OmJpb3ZpZTE6YzpvOjljNzMyZTM5MjY4ZDg1MDA1MDcyODAxYjc4YTlmZjRhOjc6M2YzZDplMDExYjFiYzU2OWZjYzEyN2QzZmNkMzg3ZDJiNzljNmIwZDAyZTE2NjdkNzcwZDc5YzU5M2FhOTMzMWE1ZjYzOnA6RjpG) http://kinase.com/ and http://kinase.com/human/kinome/ |
| Epigenetic Inflammation Scoring | Smith HM et al, 2024. doi: 10.1186/s13148-024-01661-7  Stevenson AJ et al, 2020. doi: 10.1186/s13148-020-00903-8  Verschoor CP et al, 2023. doi: 10.1111/acel.13863  Gadd DA et al, 2022. doi: 10.7554/eLife.94481  Cited references below |
| Epigenetic Scoring Diseases in Aging | Gadd DA et al, 2022. doi: 10.7554/eLife.94481 |
| SOMAScan and OLINK | Gadd DA et al, 2022. doi: 10.7554/eLife.94481 |
| Aging and Cognition | Smith HM et al, 2024. doi: 10.1186/s13148-024-01661-7  Tin A et al, 2023. doi: 10.1038/s42003-023-05454-1  Caramaschi D et al, 2022. doi: 10.1038/s41380-022-01441-w |
| Obesity/Body Mass Index | EWAS Catalog β, 2024. [https://ewascatalog.org/?efo=EFO_0001073](https://protect.checkpoint.com/v2/r01/___https://ewascatalog.org/?efo=EFO_0001073___.YzJ1OmJpb3ZpZTE6YzpvOjljNzMyZTM5MjY4ZDg1MDA1MDcyODAxYjc4YTlmZjRhOjc6YTY2Zjo1MjkyNTM0MDE4Y2UyNmRmMWVlOTM5ZjRiNzA5NjM1OGE3ZDcyZTc2NGY3ZmQ3NGEwMzA3MTZkZTRlNTJmOWJmOnA6RjpG) and  [https://ewascatalog.org/?trait=body%20mass%20index](https://protect.checkpoint.com/v2/r01/___https://ewascatalog.org/?trait=body%20mass%20index___.YzJ1OmJpb3ZpZTE6YzpvOjljNzMyZTM5MjY4ZDg1MDA1MDcyODAxYjc4YTlmZjRhOjc6NWIyZTo3Yzk5ZWQzOGFhMTJhMGE3NmNlODEwZjQ4ZWVjZDdiNTY4ZjljOTI3MWZmODFjNGQ3ZjVmNDFhNDAzMTE3NmMwOnA6RjpG) |
| Alzheimer’s Disease – Proteomics | Ali M et al, 2025. doi: 10.1038/s41591-025-03833-1. Erratum: doi: 10.1038/s41591-025-03970-7 |
| Alzheimer’s Disease – Increased Microglial Transcripts | Kosoy R et al, 2025. doi: 10.1038/s41593-025-02020-2 |
| Alzheimer’s Disease Nominated** Target Genes (NOMINATED**)** | Agora. n.d. <https://agora.adknowledgeportal.org/genes/Nominated-targets> |
| Transcription Factors | Ma’ayan Laboratory, Computational Systems Biology Enrichr. https://maayanlab.cloud/Enrichr/#metadata  See [Libraries](https://maayanlab.cloud/Enrichr/#libraries): [**ChEA 2022**](https://maayanlab.cloud/chea3/) andENCODE **(various)** and ChEA Consensus TFs from ChIP-X |

*Braak staging scores indicate brain region involvement of neurofibrillary tangles.

**Nominated genes that may be good targets for new Alzheimer's disease treatment or prevention identified using computational analyses of high-dimensional genomic, proteomic, and/or metabolomic data derived from human samples by members of the National Institute on Aging’s Accelerating Medicines Partnership in Alzheimer’s Disease. (AMP-AD) consortium. **Specific Gene Library**

Genes below include those with increased transcripts shared with genes identified as having significant logFC in promoter DNA methylation difference between bezisterim vs placebo for AD dementia CDR or Braak* stage, proteomics FDR *P* < 0.05, and other genes identified in the general references above. In addition, specific references associated with pathways and mechanisms are indicated below.

| **Gene/**  **Protein** | **Pathway/ Mechanism** | **Resource (DOI or Website Link)** |
| --- | --- | --- |
| ***AAK1*** | Inflammation | Lian J et al, 2023. doi: 10.1016/j.ymthe.2023.01.025 |
|  | Kinase cascade | Siao W et al, 2023. doi: 10.1093/plcell/koad141 |
|  | Phosphoprotein | Ferrer I et al, 2021. doi: 10.1111/bpa.12996 |
|  | Cognition | DePrimo SE et al, 1998. doi: 10.1006/meth.1998.0644 |
|  | T2D | Kjaergaard J et al, 2025. doi: 10.1016/j.xcrm.2025.102163 |
| ***ABCA2*** | Polarization | Davis Jr W., Tew HD. 2018. doi: 10.1016/j.bcp.2017.11.018 |
|  | Inflammation |  |
|  | Lipids |  |
|  | Cholesterol | Calpe-Bierdiel L.et al, 2012. doi: 10.1016/j.atherosclerosis.2012.05.039 |
|  | Phosphoprotein | Storaczyk EI. et al, 2011. doi: 10.2174/138920111795164075 |
| ***ABHD14A*** | Cognition | Yang J. et al, 2025. doi: 10.1016/j.tjnut.2025.03.015 |
|  | Dementia |  |
|  | T2D | Lord CC. et al, 2013. doi: 10.1016/j.bbalip.2013.01.002 |
|  | Lipids |  |
|  | Significantly ↑ AD Plasma Proteomics | Ali M, et al, 2025. doi: 10.1038/s41591-025-03833-1. Erratum: doi: 10.1038/s41591-025-03970-7 |
| ***ABHD14B*** | Glycolysis | Rajendren A. et al 2022. doi: 10.1016/j.jbc.2022.102128 |
| ***ABR***  *a GTPase-activating protein (GAP) for Rac1; can influence Rac1 activity* | Cognition | Wu W. et al, 2019. doi: 10.1007/s13238-019-0641-0  Zhang H. et al, 2022. doi: 10.3389/fnagi.2022.914491 |
|  | Microglial neuroinflammation | D'Ambrosi N, et al, 2014. doi: 10.3389/fncel.2014.00279. |
|  | Glycolysis | Ganapathy-Kanniappan S., 2020. doi: 10.1080/15384047.2020.1809923 |
|  | M1 Polarization | Fu H. et al, 2023. doi: 10.1038/s41419-023-06150-y. |
|  | Mitochondrial damage | Kowluru RA. et al, 2021. doi: 10.1038/s41598-021-93420-4 |
|  | Obesity | Sun M, et al, 2012. doi: 10.1038/oby.2012.6 |
|  | Lipids | Pacia MZ. et al, 2022. doi: 10.1007/s00018-022-04362-7  Hasegawa K. et al, 2023. doi: 10.3390/ijms24054608 |
|  | Significantly ↑ AD & PD Plasma Proteomics | Ali M, et al, 2025. doi: 10.1038/s41591-025-03833-1. Erratum: doi: 10.1038/s41591-025-03970-7 |
| ***ACACA*** | Polarization | Yeudall S. et al, 2022. doi: 10.1126/sciadv.abq1984. |
|  | Inflammation |  |
|  | Carbohydrate |  |
|  | Lipids |  |
|  | T2D | Wei X. et al, 2016. doi: 10.1038/nature20117 |
| ***ACAP1*** | Inflammation | Lee S. et al, 2014. doi: 10.1016/j.cmet.2014.01.013 |
|  | Polarization |  |
|  | Obesity | MacDonnel PC. et al, 1975. doi: 10.1042/bj1500269. |
|  | T2D |  |
| ***ACAT2*** | Lipids | Rudel LL. et al, 2005. doi: 10.1161/01.ATV.0000166548.65753.1e  Valencia-Olvera AC. et al, 2023. doi: 10.1007/s13311-023-01375-3 |
|  | Inflammation |  |
|  | Cognition |  |
|  | T2D | Zhu Y. et al, 2021. doi: 10.1016/j.metabol.2021.154861 |
|  | Carbohydrate |  |
|  | Polarization | Brewer HB Jr., 2000. doi: 10.1172/JCI9664 |
| ***ACBD4*** | Lipids | NIH. MedlinePlus. [https://medlineplus.gov/genetics/gene/mmut/#:~:text=Methylmalonyl%20CoA%20mutase%20is%20responsible,fats%20(lipids)%20and%20cholesterol](https://protect.checkpoint.com/v2/r01/___https://medlineplus.gov/genetics/gene/mmut/___.YzJ1OmJpb3ZpZTE6YzpvOjljNzMyZTM5MjY4ZDg1MDA1MDcyODAxYjc4YTlmZjRhOjc6NzYyMzozMmVlNmNmNWQwYTBmZGZiNDdhNGZmYjZiNmE4ZTZhZTY3YTMyZjk3YTA3NzllYzU4M2U3ZWZjZTA2YzA2MjRhOnA6RjpG#:~:text=Methylmalonyl%20CoA%20mutase%20is%20responsible,fats%20(lipids)%20and%20cholesterol). |
| ***ACOT8*** | Inflammation | Li BR. et al, 2025. doi: 10.1038/s41401-025-01477-y  Tripathy D. et al, 2003. doi: 10.2337/diabetes.52.12.2882 |
|  | Ferroptosis |  |
|  | Lipids (FFA’s) |  |
|  | Polarization |  |
|  | T2D | Wang J. et al, 2023. doi: 10.7554/eLife.87419 |
| ***ACOT9*** | Inflammation | Jiang X, Ding WX, 2020. doi: 10.1002/hep.31450  Steensels S. et al, 2020. doi: 10.1002/hep.31409 |
|  | Lipids |  |
|  | Obesity |  |
|  | T2D |  |
| ***ACP5*** | Neuro-inflammation | Zhao YT. et al, 2022. doi: 10.1186/s12974-022-02503-0 |
|  | Cognition |  |
|  | Obesity | Lång P, et al, 2021. doi: 10.1002/1873-3468.14184 |
|  | Kinase cascade |  |
| ***ACP6***  *Hydrolyzes LPA to monoacyl-glycerol, which is cleaved by mono-glyceride lipase* | Aβ & Neuro-inflammation | Chen R. et al, 2012. doi: 10.1016/j.celrep.2012.09.030 |
|  | Lipids |  |
|  | Cognition |  |
|  | M1 Polarization | Gu C. et al, 2023. doi: 10.3892/mmr.2023.13004 |
|  | T2D | Taschler U. et al, 2011. doi: 10.1074/jbc.M110.215434 |
|  | Obesity | Yoshida K, et al, 2019. doi: 10.1096/fj.201801203R |
|  | Kinase Cascade | Zhong P. et al, 2014. doi: 10.1038/npp.2014.24. |
| ***ACVR1B*** | Inflammation | Gauthier T. et al, 2025. doi: 10.1172/JCI187063 |
|  | M1 Polarization | Sierra-Filardi E. et al, 2011. doi: 10.1182/blood-2010-09-306993 |
|  | Obesity | Lee ES. et al, 2023. doi: 10.1016/j.jbc.2022.102716 |
|  | Kinase cascade | Liu PP. et al, 2016. doi: 10.1530/REP-16-0262 |
|  | Phosphoprotein | Du R. et al, 2024. doi: 10.1016/j.bcp.2024.116061 |
| ***ADIPOR1*** | Glycolysis | Manly SJ. et al, 2021. doi : 10.1158/1538-7445.AM2021-2467  Manley SJ. et al, 2022. doi: 10.1038/s41419-022-04572-8. Erratum: doi: 10.1038/s41419-022-04615-0 |
| ***ADIPOR2*** | Obesity | Liu Y. et al, 2007. doi: 10.1210/en.2006-0708. |
|  | Glycolysis | Manley SJ. et al, 2022. doi: 10.1038/s41419-022-04572-8 |
| ***ADK*** | Inflammation | Ahmad S. et al, 2014. doi: 10.1016/j.jneuroim.2014.10.006  Sun S. et al, 2022. doi: 10.3389/fcell.2022.827714.  Xu Y. et al, 2017. doi: 10.1038/s41467-017-00986-7 |
|  | Lipids/ inflammation | Li H. et al, 2023. doi: 10.1053/j.gastro.2022.09.027 |
|  | Cognition | Yee BK. Et al, 2007. doi: 10.1111/j.1460-9568.2007.05897.x |
| ***AGAP3*** | Inflammation | Zhao K. et al, 2022. doi: 10.3389/fnagi.2022.901972 |
|  | Polarization |  |
|  | Cognition |  |
|  | AD Progression |  |
| ***AGGF1*** | M1 Polarization | Liu G, Zhang H. 2024. doi: 10.4149/gpb_2024001 |
|  | Kinase cascade | Wang J. et al, 2021. doi: 10.1161/ATVBAHA.121.316867 |
|  | ERK |  |
|  | Obesity | Shao J. et al, 2020. doi: 10.1016/j.bbrc.2020.04.016 |
| ***AHCY*** | Inflammation | Liu L. et al, 2024. doi: 10.1016/j.jdermsci.2024.10.004 |
|  | ROS |  |
|  | Obesity | Boczki P. et al, 2024. doi: 10.1080/21623945.2023.2290218 |
|  | Significantly ↑ AD microglial transcripts with Dementia | Kosoy R. et al, 2025. doi: 10.1038/s41593-025-02020-2 |
|  | Significantly ↑ AD Plasma Proteomics | Ali M, et al, 2025. doi: 10.1038/s41591-025-03833-1 Erratum: 2025. doi: 10.1038/s41591-025-03970-7 |
| ***AHCYL1*** | Cognition (tau) | Stahl E. et al, 1965. doi: 10.1002/ardp.19652980908  Goto JI. Et al, 2022. doi: 10.1101/lm.053542.121 |
|  | Inflammation (↓autophagy,  ↑ apoptosis) | Huang W. et al, 2022. doi: 10.1080/15548627.2021.1924038 |
|  | Phosphoprotein | Budnik N. et al, 2024. doi: 10.1016/j.bbamcr.2024.119819 |
|  | Oxidative stress | Kiefer H. et al, 2009. doi: 10.1074/jbc.M807136200 |
|  | NOMINATED** | Agora. [https://agora.adknowledgeportal.org/genes/ENSG00000168710](https://protect.checkpoint.com/v2/r01/___https://agora.adknowledgeportal.org/genes/ENSG00000168710___.YzJ1OmJpb3ZpZTE6YzpvOjljNzMyZTM5MjY4ZDg1MDA1MDcyODAxYjc4YTlmZjRhOjc6N2JhMDphZjhlYWZhNjMxMTFhMTRmZGRiNjI5NmM2YTNlMjkwNzk0ZmQ2NjI3YjYyNWM3MjE2N2U1OWFiNTM0NTg1NzY4OnA6RjpG) |
| ***ALDH4A1*** | Inflammation | Xia J. et al, 2020. doi: 10.1002/mco2.195 |
|  | ROS |  |
|  | Neuro-degeneration | Nagaoka A. et al, 2020. doi: 10.1016/j.jpsychires.2020.02.001 |
|  | Lipid | Lorenzo C. et al, 2021. doi: 10.1038/s41586-020-2993-2 |
|  | NOMINATED** | Agora. [https://agora.adknowledgeportal.org/genes/ENSG00000159423](https://protect.checkpoint.com/v2/r01/___https://agora.adknowledgeportal.org/genes/ENSG00000159423___.YzJ1OmJpb3ZpZTE6YzpvOjljNzMyZTM5MjY4ZDg1MDA1MDcyODAxYjc4YTlmZjRhOjc6MDBjNjoxOGIzOWE1NmNkYzUyNTI5YjMyYzYzZWNhN2FhMWU3MDlmNjU0NTQ0M2ZkMDJjMGYxMGYyMTI1MzcwYTNjZjYxOnA6RjpG) |
| ***ALG3*** | Inflammation | Loke I. et al, 2016. doi: 10.1016/j.mam.2016.04.004 |
|  | Immune infiltration |  |
|  | Vascular monocyte recruitment |  |
|  | Atherosclerosis |  |
|  | Significantly ↑ AD microglial transcripts with Dementia | Kosoy R. et al, 2025. doi: 10.1038/s41593-025-02020-2 |
| ***AMPD2*** | Obesity | Yang H. et al, 2023. doi: 10.1016/j.mce.2023.112039 |
|  | T2D |  |
| ***AMPD3*** | Neuro-degeneration | Sims B. et al, 1998. doi: 10.1016/s0197-4580(98)00083-9 |
|  | Inflammation | Li P. et al, 2007. doi: 10.1253/circj.71.591 |
|  | Reperfusion injury |  |
|  | Ferroptosis |  |
|  | Cognition | Xu H. et al, 2025. doi: 10.1186/s13040-025-00432-1 |
|  | T2D | Tatekoshi Y. et al, 2018. doi: 10.1016/j.yjmcc.2018.05.003 |
| ***ANKRA2*** | M1 Polarization | Zhang S. et al, 2023. doi: 10.1016/j.jtauto.2023.100228 |
|  | Obesity (aging) | Sun XY. et al, 2011. doi: 10.1371/journal.pone.0014605 |
|  | Lipids | Upadhyay RK, 2015. doi: 10.1155/2015/971453 |
|  | Cholesterol |  |
|  | Significantly ↑ AD Plasma Proteomics | Ali M, et al, 2025. doi: 10.1038/s41591-025-03833-1 Erratum: 2025. doi: 10.1038/s41591-025-03970-7 |
| ***ANP32E*** | Glycolysis | Liu J. et al, 2024. doi: 10.1515/biol-2022-0817  Huang J et al, 2020. doi: 10.1016/j.gene.2020.144681 |
|  | Phosphoprotein | NIH 2025. [https://www.ncbi.nlm.nih.gov/gene/81611](https://protect.checkpoint.com/v2/r01/___https://www.ncbi.nlm.nih.gov/gene/81611___.YzJ1OmJpb3ZpZTE6YzpvOjljNzMyZTM5MjY4ZDg1MDA1MDcyODAxYjc4YTlmZjRhOjc6MWZjNjoxOTZmNjYwN2ViNjQ0YTAyOTU1MTlmZjBjYjYxMTY0ODU3NzBlNDcwZjdmZmQ0MjIzMWI0Y2FjYWNjNTU1OTc1OnA6RjpG) |
| ***ANXA6*** | M1 Polarization | Huang B et al, 2023. doi: 10.1007/s13258-023-01410-9  Erratum: doi: 10.1007/s13258-023-01425-2 |
|  | Inflammation (MCP1) | Sakwe NI et al, 2024. doi: 10.1101/2024.10.22.619710 |
|  | Glycolysis | Williams SD et al, 2022. doi: 10.3390/cancers14051108  Williams SD et al, 2022. doi: 10.3390/cells11193007 |
|  | T2D | Stogbauer F et al, 2009. doi: 10.3858/emm.2009.41.7.055 |
|  | Obesity |  |
|  | Lipid |  |
|  | Cholesterol |  |
| ***ANXA11*** | Cognition (AD) | Robinson JL et al, 2024. doi: 10.1007/s00401-024-02753-7 |
|  | Neuro-degeneration |  |
|  | Significantly ↑ AD & PD Plasma Proteomics | Ali M, et al, 2025. doi: 10.1038/s41591-025-03833-1 Erratum: 2025. doi: 10.1038/s41591-025-03970-7 |
| ***APBB3*** | Cognition (Aβ) | Tanahashi H et al, 1999. doi: 10.1016/s0304-3940(98)00995-1 |
|  | Microglial neuro-inflammation |  |
|  | Glycolysis | Santangelo R et al, 2021. doi: 10.18632/aging.203330 |
|  | T3D | Nguyen TT et al, 2020. doi: 10.3390/ijms21093165 |
| ***APPL2*** | Microglial inflammation | Gao C et al, 2020. doi: 10.1007/s12264-020-00514-6 |
|  | Altered NSC differentiation |  |
|  | T2D | Cheng KK et al, 2014. doi: 10.2337/db14-0337 |
|  | Significantly ↑ FTD Plasma Proteomics | Ali M, et al, 2025. doi: 10.1038/s41591-025-03833-1 Erratum: 2025. doi: 10.1038/s41591-025-03970-7 |
| ***ARF6*** | Cognition (Aβ) | Tang W et al, 2015. doi: 10.1186/s13041-015-0129-7 |
|  | Inflammation (TLR signaling) | Wu JY et al, 2012. doi: 10.1074/jbc.M111.295113 |
|  | Microglial neuro-inflammation | D'Egidio F et al. 2024. doi: 10.1016/j.nbd.2024.106663 |
|  | M1 polarization | Jiménez-García L et al, 2016. doi: 10.18632/oncotarget.11652 |
|  | Ferroptosis | Geng D et al, 2022. doi: 10.21037/jgo-22-341 |
|  | Significantly ↑ AD Plasma Proteomics | Ali M, et al, 2025. doi: 10.1038/s41591-025-03833-1 Erratum: 2025. doi: 10.1038/s41591-025-03970-7 |
| ***ARHGAP25*** | Inflammation | Czárán D et al, 2023. doi: 10.3389/fimmu.2023.1182278  Czárán D et al, 2025. doi: 10.3389/fimmu.2025.1509713 |
|  | Kinase cascades | Thuault S et al, 2016. doi: 10.1091/mbc.E16-01-0041 |
|  | Phosphoprotein | Wisniewski É et al, 2022. doi: 10.1096/fj.202200689R |
| ***ARRB2*** | M1 polarization | Wei X et al, 2024. doi: 10.1016/j.cmet.2024.08.010 |
|  | Inflammation | Zeng LX et al, 2015. doi: 10.1038/mi.2014.104 |
|  | Aβ | Woo JA et al, 2020. doi: 10.1073/pnas.1917194117  Thathiah A et al, 2013. doi: 10.1038/nm.3023 |
|  | Glycolysis | Dong T, et al, 2024. doi: 10.1038/s41467-024-45167-5 |
|  | T2D | Pydi SP et al, 2019. doi: 10.1038/s41467-019-11003-4 |
|  | Obesity |  |
|  | Kinase cascade | Kahsai AW et al, 2023. doi: 10.1073/pnas.2303794120 |
|  | Phosphoprotein | Cassier E et al, 2017. doi: 10.7554/eLife.23777 |
|  | Lipid | Kim K et al, 2024. doi: 10.1038/s44319-024-00239-x |
| ***ARRDC3*** | T2D | Batista TM et al, 2020. doi: 10.1073/pnas.1922370117 |
|  | Obesity | Batista TM et al, 2020. doi: 10.1073/pnas.1922370117  Patwari P et al, 2011. doi: 10.1016/j.cmet.2011.08.011 |
|  | Inflammation | Liu YG et al, 2020. doi: 10.1172/jci.insight.135849 |
|  | Glycolysis | Dong T et al, 2024. doi: 10.1038/s41467-024-45167-5 |
|  | Phosphoprotein | Caplan M et al, 2025. doi: 10.1016/j.jbc.2025 |
|  | Lipid | Tang Y et al, 2025. doi: 10.1016/j.metabol.2025.156175 |
| ***ASF1B*** | Inflammation | Lin HY et al, 2024. doi: 10.1182/blood.2023022079 |
|  | Immune cell infiltration | Zhang S et al, 2022. doi: 10.3389/fgene.2022.842351 |
|  | Significantly ↑ AD Plasma Proteomics | Ali M, et al, 2025. doi: 10.1038/s41591-025-03833-1 Erratum: 2025. doi: 10.1038/s41591-025-03970-7 |
| ***ATAD3A*** | Neuropathology | Zhao Y et al, 2022. doi: 10.1038/s41467-022-28769-9 |
|  | Cognition |  |
|  | Cholesterol |  |
|  | Lipid | Tang Y et al, 2025. doi: 10.1016/j.metabol.2025.156175 |
|  | Senescence & aging | He Y et al, 2025. doi: 10.1002/advs.202404109 |
|  | Neuro-inflammation | Zhao Y et al, 2022. doi: 10.1038/s41467-022-28769-9 |
|  | Significantly ↑ FTD Plasma Proteomics | Ali M, et al, 2025. doi: 10.1038/s41591-025-03833-1 Erratum: 2025. doi: 10.1038/s41591-025-03970-7 |
| ***ATE1-AS1*** | Neuro-degeneration | Fina ME et al, 2021. doi: 10.1038/s41598-021-88628-3 |
|  | Cognition |  |
|  | Significantly ↑ AD Plasma Proteomics with Dementia | Kosoy R et al, 2025. doi: 10.1038/s41593-025-02020-2 |
| ***ATG16L2*** | AD | Don Wai Luu L et al, 2022. doi: 10.1080/15548627.2022.2042783  Caberlotto L et al, 2019. doi: 10.1038/s41598-019-39828-5 |
|  | T2D |  |
| ***ATIC*** | Cognition | Alonso-Andrés P et al, 2018. doi: 10.1111/bpa.12592 |
|  | Altered purine metabolism |  |
|  | Significantly ↑ AD Plasma Proteomics with Dementia | Kosoy R et al, 2025. doi: 10.1038/s41593-025-02020-2 |
| ***ATR*** | Inflammation-driven DNA vulnerability | Li C et al, 2024. doi: 10.1016/j.compbiomed.2024.108776  Ali MM et al, 2022. doi: 10.1080/15592294.2021.1876285  Madakashira BP et al, 2024. doi: 10.1093/nar/gkae031 |
|  | ↓ Inflammation restores ATR |  |
|  | Kinase cascade | Shigechi T et al, 2012. doi: 10.1158/0008-5472.CAN-11-2904  Zhao H et al, 2001. doi: 10.1128/MCB.21.13.4129-4139.2001 |
| ***AUP1*** | Microglial neuro-inflammation | Chang PC et al, 2023. doi : 10.1186/s12935-023-02912-y |
|  | Significantly ↑ AD Plasma Proteomics with Dementia | Kosoy R et al, 2025. doi: 10.1038/s41593-025-02020-2 |
| ***AZI2*** | TBK1 Kinase | Ujevic A et al, 2024. doi: 10.1038/s41467-024-54399-4 |
|  | Inflammation | Wei M et al, 2022. doi: 10.1093/femspd/ftac016 |
| ***B3GALT4*** | Lipid (GM1) | NIH 2025. [https://www.ncbi.nlm.nih.gov/gene/8705](https://protect.checkpoint.com/v2/r01/___https://www.ncbi.nlm.nih.gov/gene/8705___.YzJ1OmJpb3ZpZTE6YzpvOjljNzMyZTM5MjY4ZDg1MDA1MDcyODAxYjc4YTlmZjRhOjc6ZTk0ODpkYWU4MjFjNWRkNTU5MDNhZDc2ZmM0MmNlNzQ2ZGY3NjcwMjBiYzQxNWY5NGU3MDJlYzBhMTU2YzRjYmQwZGYzOnA6RjpG) |
|  | Aβ (via GM1) | Matsubara T et al, 201. doi: 10.1021/acs.langmuir.7b02091  Yanagisawa K. 2015. doi: 10.1007/s10719-015-9579-5  Yuyama K et al, 2010. doi: 10.1016/j.neulet.2010.06.080  Hayashi H et al, 2004. doi: 10.1523/JNEUROSCI.0861-04.2004  Oikawa N et al, 2015. doi: <https://doi.org/10.1371/journal.pone.0121356> |
|  | Cognition (LOAD) | Madrid A et al, 2018. doi: 10.3233/JAD-180592. |
|  | Microglial neuro-inflammation | Wang X et al, 2021. doi: 10.3389/fncel.2021.723308 |
|  | Kinase cascade | Sha YL et al, 2022. doi: 10.1186/s13046-022-02523-x |
| ***BAZ2A*** | Proteostasis | Gallrein C et al, 2023. doi: 10.1016/j.celrep.2023.113577 |
|  | Age acceleration |  |
|  | ↓ Choline acetyl-transferase |  |
|  | Inflammation | Jia Y et al, 2022. doi: 10.1016/j.stem.2022.01.001 |
| ***BCAT1*** | Inflammation | Papathanassiu AE et al, 2017. doi: 10.1038/ncomms16040 |
|  | Aβ | Harris M et al. 2020. doi : http://dx.doi.org/10.1016/j.freeradbiomed.2020.01.019 |
|  | T2D | Wang J et al, 2025. doi: 10.1167/iovs.66.6.59 |
|  | M1 Polarization | Huang H et al, 2024. doi: 10.1186/s10020-024-00894-9 |
|  | α-KG | Wang J et al, 2025. doi: 10.1167/iovs.66.6.59 |
|  | H3K4me3 |  |
|  | Kinase cascade | Shafei MA et al, 2020. doi: 10.18632/oncotarget.27607 |
| ***BCL3*** | Microglial neuro-inflammation | Ma J et al, 2024. doi: 10.1016/j.neuroscience.2023.11.025  Palmer S et al, 2008. doi: 10.1007/s12026-008-8075-4 |
|  | Inflammation | Liu H et al, 2022. doi: 10.3389/fimmu.2022.847699 |
|  | Lipid | Zhang S et al, 2021. doi: 10.2147/JIR.S327858 |
|  | Obesity |  |
|  | T2D |  |
|  | AD cognition | Smith JA et al, 2021. doi: 10.1016/j.socscimed.2018.11.019  Ding Y et al, 2023. doi: 10.1002/ibra.12106 |
|  | Aβ | Palmer S et al, 2008. doi: 10.1007/s12026-008-8075-4 |
|  | NFκB |  |
|  | TF |  |
| ***BMP8B*** | Tau | Affaneh A et al, 2025. doi: 10.1002/ana.27149 |
|  | Inflammation | Vacca M et al, 2020. doi: 10.1038/s42255-020-0214-9 |
| ***BTBD11*** | Cognition (anxiety, ↓exploratory drive*)* | Bygrave AM et al, 2023. doi: 10.1016/j.celrep.2023.112591 |
|  | Obesity | Watanabe K et al, 2019. doi: 10.1016/j.heliyon.2019.e02777 |
|  | Lipids |  |
|  | T2D |  |
| ***BHLHE40*** | Microglial neuro-inflammation | Ma C et al, 2025. doi: 10.1016/j.brainresbull.2024.111139 |
|  | Polarization |  |
|  | Glycolysis |  |
|  | Inflammation |  |
|  | T2D | Tsuyama T et al, 2023. doi: 10.15252/embr.202256227  Tian J et al, 2018. doi: 10.7554/eLife.36826 |
|  | Obesity |  |
|  | Carbohydrate |  |
|  | Cognition | Hamilton KA et al, 2018. doi: 10.1371/journal.pone.0196223 |
|  | Transcription factor | Cook ME et al, 2020. doi: 10.1016/j.it.2020.09.002 |
| ***BRCC3*** | Inflammation (NLRP6) | Huang X et al, 2024. doi: 10.1111/cns.14697  Wang H et al, 2025. doi: 10.1016/j.intimp.2024.113720 |
|  | Pyroptosis |  |
|  | AD/PD Inflammasome | Daune G et al, 1988. doi: 10.1042/bj2530481 |
| ***BRD2*** *Scaffolding protein to recruit transcription factors (e.g., E2F, RNA polymerase II) to promoters of target genes* | Neuro-inflammation | Liu L et al, 2021. doi: 10.3389/fmolb.2021.748449  Wang F et al, 2009. doi: 10.1042/BJ20090928 |
|  | T2D |  |
|  | Obesity |  |
|  | Cognition | Martella N et al, 2023. doi: 10.3390/biomedicines11030750 |
|  | Lipid | Lin Z et al, 2022. doi: 10.3389/fphar.2022.887991 |
|  | Glycolysis |  |
|  | Significantly ↑ AD Plasma Proteomics | Ali M, et al, 2025. doi: 10.1038/s41591-025-03833-1 Erratum: 2025. doi: 10.1038/s41591-025-03970-7 |
| ***BTK*** | Microglial neuro-inflammation | Hodges H et al, 1998. doi: 10.1016/s0166-4328(97)00108-3  Xu LL et al, 2024. doi: 10.1186/s12974-024-03187-4  Jiang Y et al, 2025. doi: 10.1186/s10020-025-01203-8 |
|  | Polarization |  |
|  | T2D | Zhao J et al, 2021. doi: 10.3892/ijmm.2021.5010  Li Y et al, 2021. doi: 10.1016/j.bbrc.2021.02.094 |
|  | Obesity |  |
|  | Cognitive aging | Ekpenyong-Akiba AE et al, 2020. doi: 10.1111/acel.13079 |
|  | Significantly ↑ AD & PD Plasma Proteomics | Ali M, et al, 2025. doi: 10.1038/s41591-025-03833-1 Erratum: 2025. doi: 10.1038/s41591-025-03970-7 |
|  | NOMINATED** | Agora. [https://agora.adknowledgeportal.org/genes/ENSG00000010671](https://protect.checkpoint.com/v2/r01/___https://agora.adknowledgeportal.org/genes/ENSG00000010671___.YzJ1OmJpb3ZpZTE6YzpvOjljNzMyZTM5MjY4ZDg1MDA1MDcyODAxYjc4YTlmZjRhOjc6MWIwYToyOGUxMmU3OTUyYzA5N2ZhMjg1ZWUwZjhmYjQ4NWRhZDI2YWM4NzkyMjk4Y2M3MGQwYjA2ZmJkNjdjOWY2MjlkOnA6RjpG) |
| ***C1RL*** | Proteolysis | Wicher KB, Fries E, 2004. doi: 10.1073/pnas.0405692101  Zhao H et al, 2025. doi: https://doi.org/10.1016/j.arr.2024.102636  Zhao H et al, 2025. doi: 10.1016/j.arr.2024.102636  Wang J et al, 2020. doi: 10.1186/s12885-020-07436-6 |
|  | Microglia |  |
|  | Inflammation |  |
|  | Complement activation |  |
|  | Brain aging | Yi F et al, 2025. doi: 10.1126/sciadv.adr3757  Inside Precision Medicine, 2025. [https://www.insideprecisionmedicine.com/topics/informatics/ai-accurately-estimates-brain-age-gap-and-confirms-druggable-genes/#:~:text=For%20their%20analysis%2C%20the%20team,drugs%20with%20strong%20genetic%20support](https://protect.checkpoint.com/v2/r01/___https://www.insideprecisionmedicine.com/topics/informatics/ai-accurately-estimates-brain-age-gap-and-confirms-druggable-genes/___.YzJ1OmJpb3ZpZTE6YzpvOjljNzMyZTM5MjY4ZDg1MDA1MDcyODAxYjc4YTlmZjRhOjc6ZmY1NjpmMDI4YWUzMGJkYTg4ZDczMjc2YTNhMjViMDU5ODlmNWM4ZGU3Njc3MmMzYjRiYTVmZjM4Y2RkZjM2NWYxMDJiOnA6RjpG" \l ":~:text=For%20their%20analysis%2C%20the%20team,drugs%20with%20strong%20genetic%20support) |
|  | Cognition | Broad Institute, 2024. https://www.broadinstitute.org/news/immune-factor-brain-plays-critical-roles-neuron-function-and-aging |
|  | Obesity | Kaye S et al, 2017. doi: 10.3389/fimmu.2017.00545  AHA/ASA, 2023. https://www.ahajournals.org/doi/10.1161/circ.147.suppl_1.P322#:~:text=We%20used%20established%20protocols%20and,with%20SevO%20in%20HL%20populations |
|  | T2D |  |
| ***C2*** | Microglial & astrocyte neuro-inflammation | Batista AF et al, 2024. doi: 10.3390/ijms25020817 |
|  | Cognition |  |
|  | M1 Polarization | Zhang G et al, 2024. doi: 10.3390/cancers16050908 |
|  | T2D | Moin ASM et al. 2021. doi: 10.1016/j.athplu.2021.11.002  Steffen BT, 2023. doi: 10.1007/s00125-022-05801-7 |
| ***C6orf120*** | Inflammation | Wu YN et al, 2022. doi: 10.1016/j.cellimm.2021.104467 |
|  | M1 Polarization |  |
|  | Kinase cascade | Wang X et al, 2024. doi: 10.1111/jgh.16538 |
| ***CACNA1C*** | Cognition | Zhang Q et al, 2012. doi: 10.1038/npp.2011.242  Loganathan S et al, 2024. doi: 10.1038/s41398-024-03140-2 |
|  | Neuro-inflammation | Jiang Y et al, 2018. doi: 10.12659/MSM.908765 |
|  | Tau phosphorylation |  |
|  | Synaptic loss | Song JY et al, 2017. <https://doi.org/10.1016/j.jalz.2017.06.769> |
|  | Lipid peroxidation | Michels S et al, 2018. doi: 10.1038/s41420-018-0061-6 |
|  | Obesity | Lima-Leopoldo AP et al, 2008. doi: 10.1590/s0100-879x2008000700011 |
|  | Microglial & nerve inflammation | Boller AL et al, 2025. doi: 10.1016/j.bbi.2025.07.004 |
| ***CANCB3*** | M1 polarization | Wu Y et al, 2024. doi: 10.1038/s41598-024-66918-w |
|  | Aging |  |
|  | Obesity |  |
|  | T2D | Lee K et al, 2018. doi: 10.1016/j.celrep.2018.06.086 |
|  | Significantly ↑ AD Plasma Proteomics | Ali M, et al, 2025. doi: 10.1038/s41591-025-03833-1  Erratum: 2025. doi: 10.1038/s41591-025-03970-7 |
| ***CAMK2D*** | Obesity  T2D | Dai W et al, 2021. doi: 10.1016/j.molmet.2021.101300  Daniels L et al, 2015. doi: 10.1007/s10741-015-9498-3  Chacar S et al, 2024. doi: [10.1007/s11154-023-09855-9](https://protect.checkpoint.com/v2/r01/___https://doi.org/10.1007/s11154-023-09855-9___.YzJ1OmJpb3ZpZTE6YzpvOjljNzMyZTM5MjY4ZDg1MDA1MDcyODAxYjc4YTlmZjRhOjc6ZjIzYTo2ZjQwODVkM2RkNjNlMTk5MjY2MWI5Yzc4OWNkMzdhZmNhYWU4MjkyMDgwYjhlZTczNGVkZWZjODRhM2RiMmU5OnA6RjpG) |
|  | Aβ | Wang YJ et al, 2005. doi: 10.1016/j.brainres.2004.10.061 |
|  | pTau |  |
|  | Neuro-degeneration |  |
|  | Cognition | Ghosh A et al, 2015. doi: 10.1186/s13041-015-0166-2 |
|  | Lipid droplets | Zhang J et al, 2025. doi: 10.1002/advs.202409513  Chai B et al, 2024. doi: 10.1002/advs.202310134 |
|  | Ferroptosis |  |
|  | Glycolysis | Sabbir MG et al, 2021. doi: 10.1186/s12964-021-00778-z |
|  | Inflammasome | Willeford A et al, 2018. doi: 10.1172/jci.insight.97054 |
|  | Microglial neuro-degeneration | Rigter PMF et al, 2024. doi: 10.1016/j.ajhg.2023.12.016.  Erratum: doi: 10.1016/j.ajhg.2025.08.001  Cheng J et al, 2021. doi: 10.1126/sciadv.abe3600 |
|  | Cognition |  |
|  | Excitotoxicity |  |
|  | Significantly ↑ FTD Plasma Proteomics | Ali M, et al, 2025. doi: 10.1038/s41591-025-03833-1  Erratum: 2025. doi: 10.1038/s41591-025-03970-7 |
| ***CAP1*** | Inflammation | Lee S et al, 2014. doi: 10.1016/j.cmet.2014.01.013  Zhong J et al, 2021. doi: 10.3389/fnagi.2021.696944  Rust MB et al, 2025. doi: 10.1016/j.tcb.2025.01.007 |
|  | M1 polarization |  |
|  | Cognition |  |
| ***CAPN2*** | Cognition | Baudry M et al, 2024. doi: 10.3389/fnmol.2024.1337850  Wang Y et al, 2020. doi: 10.3390/cells912269 |
|  | Neuro-inflammation | Gao A et al, 2022. doi: 10.31083/j.fbl2701020  Podbielska M et al, 2016. doi: 10.1111/jnc.13774 |
|  | T2D | Luo X et al, 2024. doi: 10.1186/s13048-024-01407-2  Huang CJ et al, 2010. doi: 10.1074/jbc.M109.024190 |
|  | Obesity | Muniappan L et al, 2017. doi: 10.1038/s41598-017-14719-9  Miyazaki T et al, 2023. doi: 10.3390/ijms242316782 |
|  | Inflammation |  |
|  | NOMINATED** | Agora. https://agora.adknowledgeportal.org/genes/ENSG00000162909 |
| ***CARD11*** | Microglial neuro-inflammation | Shi X et al, 2021. doi: 10.1371/journal.pone.0255293  Bedsaul JR et al, 2018. doi: 10.3389/fimmu.2018.02105 |
|  | Inflammation |  |
|  | Macrophage polarization | Lee HS et al, 2024. doi: 10.1038/s12276-024-01367-z  Jin X et al, 2015. doi: 10.1155/2015/732450 |
|  | Obesity | Jin X et al, 2015. doi: 10.1155/2015/732450 |
|  | Monocyte inflammation | Shi X et al, 2021. doi: 10.1371/journal.pone.0255293 |
|  | Glycolysis |  |
| ***CASP7*** | Cognition | Ayers KL et al, 2016. doi: 10.1186/s12864-016-2725-z |
|  | Microglial neuro-inflammation & polarization | Burguillos MA et al, 2011. doi: 10.1038/nature09788  Venero JL et al, 2011. doi: 10.1038/cdd.2011.107  Grabert K et al, 2023. doi: 10.1038/s41419-023-05714-2 |
|  | Significantly ↑ AD Plasma Proteomics | Ali M, et al, 2025. doi: 10.1038/s41591-025-03833-1  Erratum: 2025. doi: 10.1038/s41591-025-03970-7 |
| ***CASP8*** | Microglial neuro-inflammation & polarization | Burguillos MA et al, 2011. doi: 10.1038/nature09788  Venero JL et al, 2011. doi: 10.1038/cdd.2011.107  Grabert K et al, 2023. doi: 10.1038/s41419-023-05714-2 |
|  | Cognition | Ke DQ et al, 2020. doi: 10.4103/1673-5374.272613  Erratum: doi: 10.4103/1673-5374.282272 |
| ***CBR3*** | Induced by TNF, IL-1b, NFκB | Malátková P et al, 2012. doi: 10.1016/j.bbrc.2012.03.002 |
|  | Microglial polarization |  |
|  | Inflammation | Ali MM et al, 2022. doi: 10.1080/15592294.2021.1876285 |
| ***CBX3*** | Chromatin | Wahab MA et al, 2024. doi: 10.3390/cancers16173026 |
|  | Transcription factor ATR | Enrichr. https://maayanlab.cloud/Enrichr/#metadata |
|  | AD | Lampinen R et al, 2022. doi: 10.3390/cells11040676 |
|  | Glycolysis | Chen LY et al, 2018. doi: 10.1016/j.bbrc.2018.04.137 |
| ***CD37*** | Microglial neuro-inflammation | Gao X et al, 2025. doi: 10.1038/s41467-025-61348-2  Galloway DA et al, 2019. doi: 10.3389/fimmu.2019.00790 Erratum: doi: 10.3389/fimmu.2019.01575  Yeo YA et al, 2012. doi: 10.1186/1742-2094-9-173 |
|  | M1 polarization | Mao X et al, 2023. doi: 10.3389/fimmu.2023.1202725 |
| ***CD46*** | Complement lysis | Kolev MV et al, 2009. doi: 10.2174/157015909787602805 |
|  | Inflammation | Ni Choileain S et al, 2011. doi: 10.1007/s00005-010-0109-7  Wang X et al, 2017. doi: 10.1002/eji.20164639 |
|  | M1 polarization |  |
|  | Neuro-inflammation | Hernangómez M et al, 2014. doi: 10.2174/1381612820666140130202911 |
| ***CD63*** | Microglial exosomes | Yang Y et al, 2018. doi: 10.1186/s12974-018-1204-7  Zhao X et al, 2023. doi: 10.2147/DDDT.S417465 |
|  | M1 polarization | Shi Y et al, 2020. doi: 10.1007/s10753-020-01236-7 |
|  | Glycolysis | Najy AJ et al, 2021. doi: 10.3390/cells10102721  Khanal S et al, 2024. doi: 10.1126/sciadv.adn5228 |
|  | AD | Hansen DV et al, 2018. doi: 10.1083/jcb.201709069 |
|  | Cholesterol | Saint-Pol J et al, 2025. doi: 10.20517/evcna.2024.92  Palmulli R et al, 2024. doi: 10.1038/s41556-024-01432-9 |
| ***CD79b*** | Microglial neuroinflammation | Touil H et al, 2023. doi: 10.1016/j.ebiom.2023.104789 |
|  | AD | Biragyn A et al, 2017. doi: 10.1007/s00281-016-0615-8  Kim K et al, 2021. doi: 10.1038/s41467-021-22479-4 |
|  | Obesity | Matz AJ et al, 2022. doi: 10.1097/IN9.0000000000000005  Shaikh SR et al, 2015. doi: 10.1111/cei.12444 |
| ***CDC25A*** | Neuro-degeneration | Ding XL et al, 2000. doi: 10.1016/S0002-9440(10)64837-7 |
|  | Microglial neuroinflammation | Pramanik SK et al, 2023. doi: 10.1021/acschemneuro.2c0047 |
|  | Cognition |  |
|  | Glycolysis | Liang J et al, 2016. doi: 10.1038/ncomms12431 |
|  | Significantly ↑ AD Plasma Proteomics | Ali M, et al, 2025. doi: 10.1038/s41591-025-03833-1  Erratum: 2025. doi: 10.1038/s41591-025-03970-7 |
| ***CDC42*** | Cognition | Zhu M et al, 2023. doi: 10.1093/brain/awad184 |
|  | Inflammation | Puls A et al, 1999. doi: 10.1242/jcs.112.17.2983 |
|  | M1 polarization | He J et al. 2024. doi: 10.1016/j.jcmgh.2024.01.023 |
| ***CDK12*** | Neuro-inflammation | Kabadi SV et al, 2012. doi: 10.1038/jcbfm.2011.117 |
|  | Polarization | Henry KL et al, 2018. doi: 10.1126/scisignal.aam8216 |
|  | Significantly ↑ FTD Plasma Proteomics | Ali M, et al, 2025. doi: 10.1038/s41591-025-03833-1  Erratum: 2025. doi: 10.1038/s41591-025-03970-7 |
| ***CDK16***  *(Promoter DNAm decreased [potentially ↑ expression] in bezisterim vs placebo)* | M2 Polarization | Qi J et al, 2024. doi: 10.18632/aging.205465 |
|  | ↓ Tau aggregation | Garwain O et al, 2020. doi: 10.1016/j.cellsig.2020.109620 |
|  | CDK5 interaction | Vervoorts J et al, 2020. doi: 10.1080/15548627.2020.1795423 |
|  | RIPK1 | Wang X et al, 2023. doi: 10.1016/j.biopha.2023.114929 |
| ***CDK19*** | Microglial neuroinflammation | Chen M et al, 2023. doi: 10.1093/nar/gkad538 |
|  | Inflammation | Cozzolino K et al, doi: 10.1101/2023.07.05.547813. Update: doi: 10.7554/eLife.100197  Steinparzer I et al, 2019. doi: 10.1016/j.molcel.2019.07.034  Kokinos EK et al, 2023. doi: 10.3390/v15061292 |
|  | Glycolysis | Butler M et al, 2020. doi: 10.3390/v12060654 |
|  | Lipid droplets |  |
|  | T2D | Sakuma K et al, 2023. doi: 10.1186/s13287-022-03220-4 |
|  | Obesity | Rabhi N et al, 2021. doi: 10.1038/s41598-021-99871-z |
|  | Total inhibition (Warning) | ICR. <https://www.icr.ac.uk/about-us/icr-news/detail/scientists-raise-warning-over-drugs-targeting-key-cancer-mechanism> |
|  | NOMINATED** | Agora. [https://agora.adknowledgeportal.org/genes/ENSG00000155111](https://protect.checkpoint.com/v2/r01/___https://agora.adknowledgeportal.org/genes/ENSG00000155111___.YzJ1OmJpb3ZpZTE6YzpvOjljNzMyZTM5MjY4ZDg1MDA1MDcyODAxYjc4YTlmZjRhOjc6ODJjODo2YWUzYjJmZDU5NDIxN2U0YTBiOTBlMWYwNTdiYjBlMzU1MTlmMTc4OTM1MDljNTViYmVhYjBiMTExMWM4MDlhOnA6RjpG) |
| ***CDK2AP1*** | Inflammation | Che Y et al, 2022. doi: 10.3389/fgene.2022.937310 |
|  | Significantly ↑ AD microglia with dementia | Kosoy R et al, 2025. doi: 10.1038/s41593-025-02020-2 |
|  | Significantly ↑ AD Plasma Proteomics | Ali M, et al, 2025. doi: 10.1038/s41591-025-03833-1  Erratum: 2025. doi: 10.1038/s41591-025-03970-7 |
| ***CELF1*** | Inflammation | Ward AJ et al, 2010. doi: 10.1093/hmg/ddq277 |
|  | Muscle wasting |  |
|  | NOMINATED** | Agora. [https://agora.adknowledgeportal.org/genes/ENSG00000149187](https://protect.checkpoint.com/v2/r01/___https://agora.adknowledgeportal.org/genes/ENSG00000149187___.YzJ1OmJpb3ZpZTE6YzpvOjljNzMyZTM5MjY4ZDg1MDA1MDcyODAxYjc4YTlmZjRhOjc6MzhmNzpkMGRlNTU1MjUyNzA3MTlhMTllZmZkNTgxZTkzZjE4OTA1NWE5NDUzNjNmMmNkOTQwODJjOWVjMmMyYmFjYzJlOnA6RjpG) |
| ***CENPM*** | Glycolysis | Valone FH et al, 1985. doi: 10.1016/0006-291x(85)90634-5  Stanke KM et al, 2021. doi: 10.3389/fmolb.2021.752404 |
| ***CEP55*** | Glycolysis | Wang G et al, 2016. doi: 10.7150/jca.15497 |
| ***CFL2*** | Inflammation | Zeng Q et al, 2021. doi: 10.3389/fnmol.2021.728184 |
|  | Significantly ↑ AD Plasma Proteomics | Ali M, et al, 2025. doi: 10.1038/s41591-025-03833-1  Erratum: 2025. doi: 10.1038/s41591-025-03970-7 |
| ***CHAC1***  *Bezisterim increased brain glutathione in MCI/mild AD subjects* | Inflammation | Sun J et al, 2024. doi: 10.3389/fcell.2024.1458716 |
|  | Glutathione depletion |  |
|  | MCI/AD | Haroon J et al, 2024. doi: 10.1097/MD.0000000000039027 |
|  | Cognition | Yadav S et al, 2019. doi: 10.1042/BCJ20190077. Erratum: 2020. doi: 10.1042/BCJ-2019-0077_COR |
|  | Calcium signaling |  |
| ***CHEK2*** | Neuron death | Mula A et al, 2023. doi: 10.14715/cmb/2023.69.9.17 |
|  | Kinase cascade | Ditch S et al, 2012. doi: 10.1016/j.tibs.2011.10.002 |
|  | pTau |  |
|  | M1 polarization | Li C et al, 2024. doi: 10.1016/j.redox.2024.103059 |
|  | Glycolysis |  |
| ***CHD1L*** | Inflammation | Zhang X et al, 2022. doi: 10.1080/21655979.2021.2019869 |
|  | ROS |  |
|  | NOMINATED** | Agora. [https://agora.adknowledgeportal.org/genes/ENSG00000149187](https://protect.checkpoint.com/v2/r01/___https://agora.adknowledgeportal.org/genes/ENSG00000149187___.YzJ1OmJpb3ZpZTE6YzpvOjljNzMyZTM5MjY4ZDg1MDA1MDcyODAxYjc4YTlmZjRhOjc6MzhmNzpkMGRlNTU1MjUyNzA3MTlhMTllZmZkNTgxZTkzZjE4OTA1NWE5NDUzNjNmMmNkOTQwODJjOWVjMmMyYmFjYzJlOnA6RjpG) |
| ***CHPF*** | Prion disease | Barret A et al, 2005. doi: 10.1074/jbc.M412635200 |
|  | Inflammation | Liao WC et al, 2021. PMID: 33791155; PMCID: PMC7994168 |
|  | ↓ Neurogenesis | Avram S et al, 2014. doi: 10.1155/2014/642798  Huynh MB et al, 2019. doi: <https://doi.org/10.1371/journal.pone.0209573> |
|  | Ferroptosis | Zhou M et al, 2025. doi: <https://doi.org/10.1101/2025.09.08.25335383> |
|  | Glycolysis | Zhong Q et al, 2023. PMID: 37424829; PMCID: PMC10326591. |
|  | Obesity | Meen AJ et al, 2023. doi: 10.3390/ijms24086884 |
| ***CLIC5*** | Inflammation | Tang T et al, 2017. doi: 10.1038/s41467-017-00227-x |
|  | Obesity | Bradford EM et al, 2010. doi: 10.1152/ajpregu.00849.2009 |
| ***CMPK2*** | Microglial neuro-inflammation | Guan X et al, 2024. doi: 10.1016/j.xcrm.2024.101522 |
|  | AD | Hoffmann A et al, 2023. doi: 10.3390/ijms24086884 |
|  | M1 polarization | Arumugam P et al, 2022. doi: 10.3389/fimmu.2022.935710 |
| ***CPEB3*** | Cognition | Qu WR et al, 2020. doi: 10.18632/aging.103404  Babu AT et al, 2025. doi: 10.1021/acs.jpcb.4c06423  Vogler C et al, 2009. doi: 10.3389/neuro.08.004.2009  Qu WR et, 2020. doi: 10.18632/aging.103404 |
|  | Glycolysis | Mejias M et al, 2020. doi: 10.1053/j.gastro.2020.03.008  TaoTt et al, 2024. <https://doi.org/10.1093/eurheartj/ehae666.3718> |
|  | Polarization | Winkler I et al,2023. doi: 10.1126/sciadv.adi6855 |
| ***CREB1*** | Inflammation | Chowdhury MAR et al, 2023. doi: 10.14348/molcells.2023.2193  Ichiki T, 2006. doi: 10.1161/01.ATV.0000196747.79349.d1 |
|  | Obesity | Qi L et al, 2009. doi: 10.1016/j.cmet.2009.01.006  Benchoula K et al, 2021. doi: 10.1016/j.bcp.2021.114531 |
|  | T2D |  |
|  | Kinase cascade | Xing J et al, 1996. doi: 10.1126/science.273.5277.959 |
|  | Phosphoprotein | Mayr B et al, 2001. doi: 10.1038/35085068 |
|  | Lipid | Yao D et al, 2020. doi: 10.3390/ani10101871 |
|  | Transcription factor | NIH, 2025. https://www.ncbi.nlm.nih.gov/gene/1385#:~:text=CREB1%20Facilitates%20GABAergic%20Neural%20Differentiation,type%20calcium%20channels%20by%20Shank3 |
| ***CREB3L2*** | pTau | Gouveia Roque C et al, 2023. doi: 10.1126/sciadv.add2671 |
|  | Aβ |  |
|  | ATF4 Transcription factor |  |
|  | Microglial neuroinflammation |  |
|  | Lipids/FFAs | Ma X et al, 2011. doi: 10.1042/BJ20101475 |
| ***CREBZF*** | Inflammation | Liu Y et al, 2024. doi: 10.1002/advs.202306685 |
|  | M1 polarization |  |
|  | T2D |  |
|  | Obesity | Zhang F et al, 2018. doi: 10.1002/hep.29926 |
|  | Transcription factor (STAT3, p53, HCF-1) | Liu YX et al, 2021. Chinese. PMID: 34708233  https://en.wikipedia.org/wiki/Transcriptional_regulation |
| ***CRY1*** | Obesity | Griebel G et al, 2014. doi: 10.3389/fendo.2014.00049 |
|  | Lipid | Gnocchi D et al, 2015. doi: 10.3390/biology4010104 |
| ***CRYZ*** | T2D (TYW3 gene) | Qi Q et al, 2012. doi: 10.1093/hmg/dds300 |
|  | PD | Akrioti E et al, 2022. doi: 10.3390/biom12070876 |
|  | ALS | Wei L et al, 2019. doi: 10.1212/NXG.0000000000000375 |
| ***CSNK1A1*** | Kinase cascade | Flajolet M et al, 2007. doi: 10.1073/pnas.0611236104 |
| ***CSNK1D*** | Kinase cascade | Flajolet M et al, 2007. doi: 10.1073/pnas.0611236104  Sharma V et al, 2024. doi: 10.1016/j.bioorg.2024.107378  Benn CL et al, 2020. doi: 10.3389/fnagi.2020.00242  Xu P et al, 2019. doi: 10.1016/j.gene.2019.144005 |
|  | Cognition | Adler P et al, 2019. doi: 10.1021/acs.jproteome.9b00312 |
|  | Microglial inflammation | Martínez-González L et al, 2020. doi: 10.1038/s41598-020-61265-y |
|  | Significantly ↑ AD Plasma Proteomics | Ali M, et al, 2025. doi: 10.1038/s41591-025-03833-1  Erratum: 2025. doi: 10.1038/s41591-025-03970-7 |
| ***CSNK1G3*** | Inflammation | Wang Q et al, 2025. doi: 10.1038/s41598-025-89275-8  Lee SY et al, 2019. doi: 10.1038/s41419-019-2146-4 |
|  | Necroptosis |  |
|  | TNF |  |
|  | Significantly ↑ FTD Plasma Proteomics | Ali M, et al, 2025. doi: 10.1038/s41591-025-03833-1  Erratum: 2025. doi: 10.1038/s41591-025-03970-7 |
| ***CSNK2B*** | pTau | Zhang Q et al, 2018. doi: 10.3389/fnmol.2018.00146 |
|  | Cognition |  |
|  | T3D | Rorbach-Dolata A, Piwowar A, 2019. doi: <https://doi.org/10.1155/2019/1435276> |
|  | Astroglial neuro-inflammation | Rosenberger AF et al, 2016. doi: 10.1186/s12974-015-0470-x |
|  | Significantly ↑ AD & FTD Plasma Proteomics | Ali M, et al, 2025. doi: 10.1038/s41591-025-03833-1  Erratum: 2025. doi: 10.1038/s41591-025-03970-7 |
| ***CSRP1*** | Neuro-inflammation | Gong Y et al. 2025. doi: 10.1038/s41467-024-54715-y  Narayanan M et al, 2014. doi: 10.15252/msb.20145304 |
|  | Glycolysis | Han L et al, 2025. doi: 10.1007/s12013-024-01610-4 |
|  | NOMINATED** | Agora. [https://agora.adknowledgeportal.org/genes/ENSG00000159176](https://protect.checkpoint.com/v2/r01/___https://agora.adknowledgeportal.org/genes/ENSG00000159176___.YzJ1OmJpb3ZpZTE6YzpvOjljNzMyZTM5MjY4ZDg1MDA1MDcyODAxYjc4YTlmZjRhOjc6YTgwMDo5ZGJhMGFiN2E2Mzc5MWEyNTY3Y2IzMWQ2OWM3NTVlMjRiZmU4NzM0ODFmOGMzMjkzNDEwMzAwNmVlNjcxNGE1OnA6RjpG) |
| ***CTSC*** | Microglial neuroinflammation | Liu Q et al, 2019. doi: 10.1186/s12974-019-1398-3  Alam S et al, 2019. doi: 10.1016/j.yexcr.2019.06.017 |
|  | M1 Polarization |  |
|  | Kinase cascade |  |
|  | Cognition | Zhang Y et al, 2018. doi: 10.1007/s11064-017-2320-y. Erratum: doi: 10.1007/s11064-018-2564-1 |
|  | Significantly ↑AD microglial transcripts with dementia | Kosoy R et al 2025. doi: 10.1038/s41593-025-02020-2 |
| ***CTSL*** | Microglial neuroinflammation | Zhang H et al, 2025. doi: 10.1096/fj.202403101R. |
|  | M1 polarization |  |
|  | Casp8 | Xu S et al, 2018. doi: 10.1016/j.neurobiolaging.2017.09.030 |
|  | NFκB |  |
|  | Kinase GSK2β | Tang Q et al, 2009. doi: 10.1007/s00109-008-0423-2  Fei Y et al, 2018. doi: 10.1016/j.cellsig.2018.01.012 |
| ***CUTA*** | Cognition | Hou P et al, 2015. doi: 10.1016/j.neurobiolaging.2014.12.005 |
|  | Aβ |  |
|  | Significantly ↑ AD Plasma Proteomics | Ali M, et al, 2025. doi: 10.1038/s41591-025-03833-1  Erratum: 2025. doi: 10.1038/s41591-025-03970-7 |
| ***DAPK2*** | Inflammation | Elbadawy M et al, 2018. doi: 10.3390/ijms19103031. Erratum: doi: 10.3390/ijms21124560 |
|  | Kinase cascade |  |
|  | Phosphoprotein |  |
|  | EPO |  |
|  | NFκB | Jiang Y et al, 2021. doi: 10.21037/atm-21-2062  Geering B, 2015. doi: 10.1016/j.biocel.2015.06.001 |
| ***DBI*** | Aging | Montégut L et al, 2023. doi: 10.1111/acel.13910 |
|  | Dementia | Ferrarese C et al, 1990. doi: 10.1212/wnl.40.4.632 |
|  | Inflammation | Nogueira-Recalde U et al, 2025. doi: 10.1038/s41418-025-01474-y |
|  | Significantly ↑AD microglial transcripts with dementia | Kosoy R et al 2025. doi: 10.1038/s41593-025-02020-2 |
|  | Braak* |  |
| ***DBT*** | Neuro-inflammation | Hwang RD et al, 2024. doi: 10.1101/2023.09.12.556394. Update: doi: 10.7554/eLife.91002 |
|  | Cognition | Salcedo C et al, 2021. doi: 10.3389/fnagi.2021.736580 |
|  | T2D | Vanweert F et al, 2022. doi: 10.1038/s41387-022-00213-3 |
|  | Obesity |  |
|  | Significantly ↑ AD Plasma Proteomics | Ali M, et al, 2025. doi: 10.1038/s41591-025-03833-1  Erratum: doi: 10.1038/s41591-025-03970-7 |
|  | NOMINATED** | Agora. [https://agora.adknowledgeportal.org/genes/ENSG00000137992](https://protect.checkpoint.com/v2/r01/___https://agora.adknowledgeportal.org/genes/ENSG00000137992___.YzJ1OmJpb3ZpZTE6YzpvOjljNzMyZTM5MjY4ZDg1MDA1MDcyODAxYjc4YTlmZjRhOjc6NGU0YjoxNjdhNWE0ZjYxNTFjNWEwMzZhYTA5YTA4ZjJhODdhNjBhYjRiNzEwOWM4NTI4ZDU3NTI3MDk2ZDZmYTU3YTg4OnA6RjpG) |
| ***DCAF6*** | Inflammation | Tseng C et al, 2023. doi: 10.1007/s00109-022-02277-1  Xu C et al, 2024. doi: 10.1016/j.jbc.2023.105566 |
| ***DDB2*** | ROS | Roy N et al, 2012. doi: 10.3390/ijms130911012 |
|  | Cognition | Borgesius NZ et al, 2011. doi: 10.1523/JNEUROSCI.1589-11.2011 |
|  | Phosphoprotein | Zhao Q et al, 2008. doi: 10.1074/jbc.M803963200 |
|  | Chromatin | Luijsterburg MS et al, 2012. doi: 10.1083/jcb.201106074 |
|  | Transcription factor |  |
| ***DDX20*** | Interferon | Chen Z et al, 2024. doi: 10.1016/j.antiviral.2024.105875 |
|  | Kinase cascade | Simankova A et al, 2021. doi: 10.1002/glia.24058 |
|  | Phosphoprotein | Hobani YH, et al, 2022. doi: 10.3390/genes13081404 |
| ***DDX41*** | Microglial inflammation | Ma J et al, 2024. doi: 10.3389/fimmu.2024.1451705  Winstone L et al, 2024. doi: 10.1042/BST20230725  Tan HY et al, 2022. doi: 10.1016/j.isci.2022.104404  Wang D et al, 2024. doi: 10.1186/s12967-024-04881-w |
|  | M1 Polarization | Tharshan Jeyakanesh J et al, 2024. doi: 10.1016/j.fsi.2024.109365 |
|  | Cognition | Weinreb JT et al, 2022. doi: 10.1002/1873-3468.14487 |
|  | Significantly ↑ FTD Plasma Proteomics | Ali M, et al, 2025. doi: 10.1038/s41591-025-03833-1  Erratum: 2025. doi: 10.1038/s41591-025-03970-7 |
| ***DDX46*** | Inflammation | Yang G et al, 2024. doi: 10.4149/neo_2024_230904N469 |
|  | Kinase cascade (JMJD6) |  |
|  | Phosphoprotein | Latourelle JC et al, 2010. doi: 10.1186/1471-2377-10-23 |
| ***DDX5*** | AD | Harel I et al, 2024. doi: 10.1016/j.celrep.2023.112787  Kar A et al, 2011. doi: 10.1128/MCB.01149-10 |
|  | Prion |  |
|  | pTau |  |
|  | Inflammation | Abbasi N et al, 2020. doi: 10.26508/lsa.202000772 |
|  | Cognition | Harel I et al, 2024. doi: 10.1016/j.celrep.2023.112787 |
|  | Glycolysis | Xing Z et al, 2017. doi: 10.1261/rna.060335.116 |
|  | Obesity | Ramanathan N et al, 2015. doi: 10.1186/s12944-015-0163-6 |
|  | TF | Zuo Q et al, 2024. doi: 10.3390/genes15070841 |
| ***DEDD2*** | Neurogenesis | Zhang W et al, 2014. doi: 10.1038/ncomms4330 |
|  | Neuro-inflammation | Roth W et al, 2002. doi: 10.1074/jbc.M110749200  Arai S et al, 2007. doi: 10.1073/pnas.0611167104 |
| ***DENND1B*** | Cognition | Cheng Z et al, 2025. https://doi.org/10.1002/alz.086245 |
|  | Obesity | Wallis NJ et al, 2025. doi: 10.1126/science.ads2145 |
|  | T2D | Chen J et al, 2013. doi: 10.1155/2013/970435 |
|  | NOMINATED** | Agora. [https://agora.adknowledgeportal.org/genes/ENSG00000137992](https://protect.checkpoint.com/v2/r01/___https://agora.adknowledgeportal.org/genes/ENSG00000137992___.YzJ1OmJpb3ZpZTE6YzpvOjljNzMyZTM5MjY4ZDg1MDA1MDcyODAxYjc4YTlmZjRhOjc6NGU0YjoxNjdhNWE0ZjYxNTFjNWEwMzZhYTA5YTA4ZjJhODdhNjBhYjRiNzEwOWM4NTI4ZDU3NTI3MDk2ZDZmYTU3YTg4OnA6RjpG) |
| ***DGAT1*** | Lipid droplet | Prakash P et al, 2025. doi: 10.1016/j.immuni.2025.04.029 |
|  | Microglial neuroinflammation | Yadav A et al, 2025. doi: 10.1101/2025.02.18.638929 |
|  | Cognition |  |
|  | T2D | Subauste A et al, 2003. doi: 10.2174/1568008033340081 |
|  | Obesity |  |
|  | M1 polarization | Morgan PK et al, 2021. doi: 10.1016/j.jbc.2021.101341 |
| ***DGKZ*** | Cognition | Mérida I et al, 2019. doi: 10.1042/BCJ20180620. |
|  | Kinase cascade |  |
|  | T2D | Benziane B et al, 2017. doi: 10.1194/jlr.M079723 |
|  | Obesity | Begin S et al, 2016. doi: 10.15761/IOD.1000148 |
|  | Cholesterol |  |
| ***DHX16*** | Inflammation | Hage A et al, 2022. doi: 10.1016/j.celrep.2022.110434 |
|  | COVID |  |
|  | Neurologic gene dysregulation | Hage A et al, 2022. doi: 10.1016/j.celrep.2022.110434  Drackley A et al, 2024. doi: 10.1002/ajmg.a.63392  Lederbauer J et al, 2024. doi: 10.3389/fnmol.2024.1414949 |
| ***DIABLO*** | Inflammation | Pandey SK et al, 2022. doi: 10.3389/fonc.2022.992260  Paul A et al, 2018. doi: 10.1016/j.ymthe.2017.12.020  Silke J et al, 2015. doi: 10.1038/ni.3206 |
|  | Apoptosis |  |
|  | NFκB |  |
|  | TNF |  |
|  | Significantly ↑ AD Plasma Proteomics | Ali M, et al, 2025. doi: 10.1038/s41591-025-03833-1  Erratum: 2025. doi: 10.1038/s41591-025-03970-7 |
| ***DLG4*** | Neuro-inflammation | Krishnan ML et al, 2017. doi: 10.1038/s41467-017-00422-w |
|  | AD |  |
| ***DLX2*** | AD | Liu MH et al, 2024. doi: 10.1002/dneu.22951  Lee SY et al, 2011. doi: 10.1186/1476-4598-10-113 |
|  | Metabolic stress |  |
|  | Necrosis |  |
|  | Glycolysis | Lee SY et al, 2015. doi: 10.3892/ijo.2015.2874 |
|  | Transcription factor | Harmonize 3, 2024. <https://maayanlab.cloud/Harmonizome/gene/DLX2#:~:text=DLX2%20is%20a%20key%20homeobox,5> |
|  | Chromatin |  |
| ***DPP4*** | Microglial neuroinflammation | Zheng Z et al, 2021. doi: 10.1186/s12974-021-02133-y |
|  | M1 polarization | Zhuge F et al, 2016. doi: 10.2337/db16-0317 |
|  | T2D | Omar B et al, 2014. doi: 10.2337/db14-0052 |
|  | Obesity | Sell H et al, 2013. doi: 10.2337/dc13-0496 |
|  | Cognition | Valverde A et al, 2021. doi: 10.1016/j.jbc.2021.100963 |
|  | Lipids | Preinfalk V et al, 2024. doi: 10.1016/j.heliyon.2024.e30329  Yuan W et al, 2021. doi: 10.3389/fonc.2021 |
|  | Fatty acids |  |
|  | Cholesterol |  |
| ***DUSP2*** | Inflammation | Hamamura K et al, 2015. doi: 10.1016/j.cellsig.2015.01.010 |
|  | Kinase cascade |  |
| ***DUSP22***  *Bezisterim subjects had lower promoter DNA methylation than placebo subjects* | Inflammation | Sanchez-Mut JV, et al, 2015. doi: 10.3389/fnbeh.2015.00347 |
|  | Kinase STAT, JNK & ERK |  |
|  | Phosphoprotein |  |
|  | Glucose | Ge C et al, 2022. doi: 10.1038/s41467-022-33493-5 |
|  | Obesity |  |
|  | Cholesterol |  |
|  | M1 polarization | Patysheva MR et al, 2023 doi: 10.3390/ijms242417542 |
|  | Cognition | An N et al, 2021. doi: 10.1016/j.pneurobio.2020.101906 |
|  | Neurologic disorders |  |
|  | AD | Sanchez-Mut JV et al, 2014. doi: 10.1002/hipo.22245 |
|  | Schizophrenia | Boks MP et al, 2018. doi: 10.1038/s41537-018-0058-4 |
|  | PTSD | Howie H et al, 2019. doi: 10.31887/DCNS.2019.21.4/kressler |
| ***E2F2*** | Microglial neuroinflammation | Xin J et al, 2025. doi: 10.1007/s10616-025-00730-w  Liu Z et al, 2024. doi: 10.1002/advs.202410880. Erratum: doi: 10.1002/advs.202502901 |
|  | M1 polarization |  |
|  | Cholesterol | Apodaka-Biguri M et al, 2025. doi: 10.1097/HEP.0000000000001461 |
|  | Glycolysis | Li XQ et al, 2024. doi: 10.1007/s10616-024-00655-w |
|  | Transcription factor | Fisher E, 1988. PMID: 3395280 |
| ***E2F7*** | Inflammation (H1F1α) | Zhou Y et al, 2022. doi: 10.1007/s00011-022-01544-8 |
|  | Myeloid cells | Cui H et al, 2024. doi: 10.1097/MD.0000000000038574 |
|  | Lipid | Tang W et al, 2025. doi: 10.1016/j.prostaglandins.2025.106988 |
|  | Phosphoprotein | Yuan R et al, 2018. doi: 10.15252/embj.201797877 |
|  | Transcription factor (p53) | Carvajal LA et al, 2012. doi: 10.1101/gad.184911.111 |
| ***E2F8*** | Glycolysis | Li XQ et al, 2024. doi: 10.1007/s10616-024-00655-w |
|  | T2D | Chen Y et al, 2019. doi: 10.1134/S0006297919120125 |
|  | Phosphoprotein | Subtil-Rodríguez A et al, 2014. doi: 10.1093/nar/gkt1161 |
|  | Chromatin |  |
| ***EARS2*** | M1 polarization | Wang L et al, 2025. doi: 10.2147/ITT.S499680 |
|  | Cholesterol |  |
|  | Neurogenic hypertension | Chan SHH et al, 2017. doi: 10.1152/physiol.00006.2017 |
|  | Cognition | Poirier R et al, 2007. doi: 10.3389/neuro.08.006.2007 |
|  | T2D | Lu L et al, 2018. doi: 10.1016/j.ygcen.2017.08.023 |
|  | Lipid |  |
|  | Obesity | Zhang J et al, 2013. doi: 10.1038/srep01476  Boyle KB et al, 2009. doi: 10.1038/cdd.2009.11 |
| ***ECHDC1*** | Lipid | Ma Y et al, 2022. doi: 10.1016/j.bbrc.2022.03.055 |
|  | Inflammation |  |
|  | T2D |  |
|  | Cognition | Fu AK et al, 2014. doi: 10.1073/pnas.1405803111 |
|  | Significantly ↑ FTD Plasma Proteomics | Ali M, et al, 2025. doi: 10.1038/s41591-025-03833-1  Erratum: 2025. doi: 10.1038/s41591-025-03970-7 |
| ***EFNA4*** | Cognition | Fu AK et al, 2014. doi: 10.1073/pnas.1405803111 |
|  | Synapse |  |
|  | Microglial neuroinflammation | Wei HX et al, 2023. doi: 10.1016/j.heliyon.2023.e18429 |
|  | M1 polarization |  |
|  | Glycolysis | Zhao X et al, 2022. doi: 10.3390/cancers14174226 |
|  | Obesity | Huang J et al, 2025. doi: 10.1016/j.ebiom.2025.105579 |
|  | Significantly ↑AD microglial transcripts with dementia | Kosoy R et al, 2025. doi: 10.1038/s41593-025-02020-2 |
|  | Significantly ↑ FTD & PD Plasma Proteomics | Ali M, et al, 2025. doi: 10.1038/s41591-025-03833-1  Erratum: 2025. doi: 10.1038/s41591-025-03970-7 |
| ***EGR1*** | Aβ | Lu Y et al, 2011. doi: 10.1074/jbc.M111.220962 |
|  | pTau |  |
|  | Kinase cascade (CDK5) |  |
|  | Microglial neuroinflammation | Yu Q et al, 2018. doi: 10.1016/j.expneurol.2018.01.009 |
|  | Obesity | Milet C et al, 2017. doi: 10.1038/s41598-017-16543-7 |
|  | Chromatin | Rocks D et, 2023 doi: 10.1101/2023.12.20.572697 |
| ***EGR2*** | Inflammation | Bo Z et al, 2022. doi: 10.1186/s12872-022-02814-3 |
|  | Microglial neuroinflammation | Yan Y et al, 2013. doi: 10.1007/s10735-013-9482-y |
|  | M1 polarization | Veremeyko T et al, 2028. doi: 10.3389/fimmu.2018.02515. Erratum: doi: 10.3389/fimmu.2018.02923 |
|  | Cognition | Chen YR et al, 2022. doi: 10.1038/s41401-022-00915-5 |
|  | Obesity | Boyle KB et al, 2009. doi: 10.1038/cdd.2009.11 |
|  | Lipids | Lu L et al, 2018. doi: 10.1016/j.ygcen.2017.08.023 |
|  | IR |  |
|  | TF | Li S et al, 2012. doi: [10.1016/j.immuni.2012.08.001](https://protect.checkpoint.com/v2/r01/___https://doi.org/10.1016/j.immuni.2012.08.001___.YzJ1OmJpb3ZpZTE6YzpvOjljNzMyZTM5MjY4ZDg1MDA1MDcyODAxYjc4YTlmZjRhOjc6M2VmNTplNGZiYmVmM2Y4YTNmOTk1Nzk1NmMzYTBlMTRkYWE4MjRhZTg4ZTYyZjFlYWJlZTZmNDAzNDE0YzMwNDY0MzE2OnA6RjpG" \t "_blank) |
|  | Chromatin | Martinez-Moreno, et al, 2022. doi: <https://doi.org/10.1101/2022.05.02.490330> |
| ***EHMT1*** | Cognition | Zheng Y et al, 2019. doi: 10.1093/brain/awy354  Zhang Z et al, 2024. doi: 10.1007/s00018-024-05176-5. |
|  | AD (AMPAR) | Lin L et al, 2019. doi: 10.3233/JAD-190190 |
|  | Epigenetics | NIH, 2025. https://www.ncbi.nlm.nih.gov/gene/79813 |
|  | Inflammation (H3K9me3) | Villeneuve LM et al, 2008. doi: 10.1073/pnas.0803623105 |
|  | T2D |  |
|  | Transcription |  |
| ***EHMT2*** | Cognition | Lin L et al, 2019. doi: 10.3233/JAD-190190  Zheng Y et al, 2019. doi: 10.1093/brain/awy354  Xiong F et al, 2023. doi: 10.1007/s12031-023-02107-0 |
|  | Aβ | Jana A et al, 2023. doi: 10.1021/acsmedchemlett.3c00344 |
|  | Microglial neuroinflammation | Yang H et al, 2023. doi: 10.1515/tnsci-2022-0276  Bellver-Sanchis A et al, 2024. doi: 10.14336/AD.2023.0424-2 |
|  | M1 polarization |  |
|  | Glycolysis | Yang LN et al, 2019. PMID: 31497345; PMCID: PMC6726997. |
|  | Obesity | Zhang W et al, 2020. doi: 10.2337/db20-0437 |
|  | Significantly ↑AD microglial transcripts with dementia | Kosoy R et al 2025. doi: 10.1038/s41593-025-02020-2 |
|  | Significantly ↑ AD & PD Plasma Proteomics | Ali M, et al, 2025. doi: 10.1038/s41591-025-03833-1  Erratum: 2025. doi: 10.1038/s41591-025-03970-7 |
|  | pTau | Wang W et al, 2021. doi: 10.1111/acel.13456 |
|  | Chromatin | Chatterjee K et al,2024. doi: 10.1101/2024.12.18.629181 |
| ***EIF1AD*** | Macrophage | De Leon-Oliva D et al, 2023. doi: 10.3390/biology12050694 |
|  | Significantly ↑ AD Plasma Proteomics | Ali M, et al, 2025. doi: 10.1038/s41591-025-03833-1  Erratum: 2025. doi: 10.1038/s41591-025-03970-7 |
| ***EIF2AK3*** | Neuro-inflammation | Guthrie LN et al, 2016. doi :10.1074/jbc.M116.738021 |
|  | Cognition | Akay-Espinoza C et al, 2024. doi: 10.1007/s11481-024-10125-x |
|  | M1 polarization | Yang F et al, 2019. doi: 10.1016/j.cellimm.2018.12.008 |
| ***EIF4E*** | Dementia | Pérez RF et al, 2022. doi: 10.1093/gerona/glac068 |
|  | Phosphoprotein |  |
|  | Microglial neuroinflammation | Amorim IS et al, 2018. doi: 10.3389/fgene.2018.00561 |
|  | M1 polarization | Chatterjee S et al, 2021. doi: 10.1016/j.isci.2021 |
|  | Obesity | Conn CS et al, 2021. doi: 10.1038/s42255-021-00349-z |
|  | Lipids |  |
| ***EIF5A*** | pTau | Desai R et al, 2025. doi: 10.1016/j.bbadis.2025.167991 |
|  | Inflammation | Anderson-Baucum E et al, 2021. doi: 10.1016/j.cmet.2021.08.003.  Tauc M et al, 2021. doi: 10.1186/s13578-021-00733-y |
|  | T2D |  |
|  | Obesity |  |
|  | M1 polarization |  |
|  | Lipids | Zhou J et al, 2022. doi: 10.1038/s41467-022-32788-x |
|  | Cognition | Liang Y et al, 2021. doi: 10.1016/j.celrep.2021.108941 |
|  | Glycolysis | Cao TT et al, 2017. doi: 10.1093/carcin/bgw119 |
| ***ELF1*** | TF | NIH, 2025. <https://www.ncbi.nlm.nih.gov/gene/1997> |
|  | Inflammation | Varghese P et al, 2025. doi: 10.1016/j.exphem.2025.104864 |
|  | NFκB |  |
|  | Kinase cascade |  |
|  | Glycolysis | Petrosino S, Grumati P, 2026. doi: 10.1152/ajpcell.00551.2025 |
| ***ELOVL1*** | Lipids | Guttenplan KA et al, 2021. doi: 10.1038/s41586-021-03960-y |
|  | Cognition | Hou SJ et al, 2024. doi: 10.1186/s40478-024-01771-6 |
|  | Neuro-inflammation | Zhang X et al, 2024. doi: 10.1186/s13195-024-01430-x |
|  | Lipids | Wang X et al, 2023. doi: 10.1186/s40001-023-01523-7 |
|  | T2D | Oh YS et al, 2018. doi: 10.3389/fendo.2018.00384 |
| ***EML2*** | Microtubules | Marasi V et al, 2025. doi: 10.1016/j.jbc.2025.110252 |
|  | Inflammation | Sun Y et al, 2024. doi: 10.62347/PALH4103 |
| ***ENO3*** | Glycolysis | Liu S et al, 2024. doi: 10.1155/2024/5200222 |
|  | Inflammation | Lu D et al, 2021. doi: 10.21037/atm-21-471 |
|  | Lipids |  |
|  | T2D | Liu S et al, 2024. doi: 10.1155/2024/5200222 |
|  | AD |  |
|  | NOMINATED** | Agora. [https://agora.adknowledgeportal.org/genes/ENSG00000108515](https://protect.checkpoint.com/v2/r01/___https://agora.adknowledgeportal.org/genes/ENSG00000108515___.YzJ1OmJpb3ZpZTE6YzpvOjljNzMyZTM5MjY4ZDg1MDA1MDcyODAxYjc4YTlmZjRhOjc6MzZkNDpjZDkzNzM0NWE1OTFjNzM2YTc2MWI5NGYzMmJiMjIxNjA0ZmQ1MDczZDg4NjYzMDllYjU4OWM3YzRjNWM4MTZhOnA6RjpG) |
| ***EPHA4*** | Microglial neuroinflammation | Kowalski EA et al, 2019. doi: 10.1186/s12974-019-1605-2 |
|  | AD | Fu AK et al, 2014. doi: 10.1073/pnas.1405803111 |
|  | M1 polarization | Wei HX et al, 2023. doi: 10.1016/j.heliyon.2023.e18429 |
|  | Kinase cascade | Zhou L et al, 2007. doi: 10.1523/JNEUROSCI.1170-07.2007 |
| ***ERBB2*** | Inflammation | Madson JG et al, 2006. doi: 10.2353/ajpath.2006.060082  Rahman A et al, 2019. doi: 10.7554/eLife.50990 |
|  | Cognition | Wang BJ et al, 2017. doi: 10.1073/pnas.1618804114 |
|  | M1 polarization | Lv C et al, 2022. doi: 10.1021/acsomega.2c00571 |
|  | Glycolysis | Zhou L et al, 2016. doi: 10.3892/or.2016.4627  Zhao YH et al, 2009. doi: 10.1038/onc.2009.229  Ma X et al, 2023. doi: 10.1007/s12032-023-02008-7 |
|  | T2D | Muhammad IF et al, 2019. doi: 10.2337/dc18-2556 |
|  | Obesity | Jian W et al, 2020. doi: 10.1186/s12967-020-02292-1 |
|  | Lipids | Gopal SM et al, 2020. doi: 10.1016/j.chemphyslip.2020.104911 |
|  | Significantly ↑ AD Plasma Proteomics | Ali M, et al, 2025. doi: 10.1038/s41591-025-03833-1  Erratum: 2025. doi: 10.1038/s41591-025-03970-7 |
| ***ETV1*** | Inflammation | Shen X et al, 2022. doi: 10.3389/fimmu.2022.939806 |
|  | Phosphoprotein | Abe H et al, 2012. doi: 10.1073/pnas.1206418109 |
|  | Chromatin |  |
|  | Demethylase |  |
|  | TF | NIH, 2025. [https://www.ncbi.nlm.nih.gov/gene/2115](https://protect.checkpoint.com/v2/r01/___https://www.ncbi.nlm.nih.gov/gene/2115___.YzJ1OmJpb3ZpZTE6YzpvOjljNzMyZTM5MjY4ZDg1MDA1MDcyODAxYjc4YTlmZjRhOjc6ZmM4Mjo3OGM5MmE1Yzk1M2M2MzdiM2M0ZWI2OGNiYjdmNzE0YjkyZmE1NmI0YWI3NDYzNmVhNTc4NWZhNDM4NTRiNmRkOnA6RjpG) |
| ***EXPH5*** | Cognition | Sell GL et al, 2017. doi: 10.1172/JCI85504 |
|  | Braak* score |  |
| ***FAM163A*** | Kinase cascade | Liu N et al, 2019. doi: 10.2147/OTT.S214731 |
|  | NOMINATED** | Agora. [https://agora.adknowledgeportal.org/genes/ENSG00000143340](https://protect.checkpoint.com/v2/r01/___https://agora.adknowledgeportal.org/genes/ENSG00000143340___.YzJ1OmJpb3ZpZTE6YzpvOjljNzMyZTM5MjY4ZDg1MDA1MDcyODAxYjc4YTlmZjRhOjc6OWViMTo2YTU1OTBhYzFlMTJlYjczYzY4MDMyODc5MjJkNTBiMTdmOGI2NTU5OTc5ZTk0YjkyZWVmNzk3ZmE0MDcyOTIyOnA6RjpG) |
| ***FBXL16***  *Promoter DNAm was decreased (potentially increased expression) in bezisterim subjects* | APP | Qu L et al, 2024. doi: 10.1186/s40364-024-00691-w |
|  | Neuro-inflammation |  |
|  | Cognition |  |
|  | T2D | Morel M, Long W, 2024. doi: 10.1002/1878-0261.13554 |
|  | Glycolysis | Kim YJ et al, 2021. doi: 10.1016/j.celrep.2021.109996 |
|  | M1 polarization | Sugimoto-Ishige A et al, 2025. doi: 10.3389/fimmu.2025.1524110 |
|  | NOMINATED** | Agora. [https://agora.adknowledgeportal.org/genes/ENSG00000127585](https://protect.checkpoint.com/v2/r01/___https://agora.adknowledgeportal.org/genes/ENSG00000127585___.YzJ1OmJpb3ZpZTE6YzpvOjljNzMyZTM5MjY4ZDg1MDA1MDcyODAxYjc4YTlmZjRhOjc6ZjY3NjpiNzljYjhkNzQ4MjM4NzEzZDA2ZDVmNTQ0MmJlZGI1ZDZhYWJjMWI1NzVkNzM3M2I2YWE0ZThiZjk5ODFhYWQ1OnA6RjpG) |
| ***FDPS*** | Inflammation | Pelleieux S et al, 2018. doi: 10.1016/j.neurobiolaging.2018.01.012  Chen Z et al, 2020. doi: 10.1111/jcmm.15542  Liu J et al, 2023. doi: 10.1096/fj.202300433RR |
|  | pTau |  |
|  | AD |  |
|  | Lipids | Wang L et al, 2023. doi: 10.3892/ol.2023.13731 |
|  | Cognition | Cheng S et al, 2013. doi: 10.1074/jbc.M113.503904 |
|  | Significantly ↑ AD & PD Plasma Proteomics | Ali M, et al, 2025. doi: 10.1038/s41591-025-03833-1  Erratum: 2025. doi: 10.1038/s41591-025-03970-7 |
| ***FGD6*** | Inflammation | Zeng J et al, 2021. doi: 10.3389/fmed.2021.672595 |
|  | Braak* score |  |
| ***FKBP1A*** | Microglial neuroinflammation | Agam G et al, 2024. doi: 10.3390/cells13100801 |
|  | pTau |  |
|  | Cognition |  |
|  | T2D |  |
|  | Obesity | Häusl AS et al, 2019. doi: 10.1016/j.molmet.2019.09.003 |
|  | T2D |  |
|  | Significantly ↑ AD Plasma Proteomics | Ali M, et al, 2025. doi: 10.1038/s41591-025-03833-1  Erratum: 2025. doi: 10.1038/s41591-025-03970-7 |
| ***FLI-1*** | Cognition | Li P et al, 2022. doi: 10.1016/j.ymthe.2022.01.023 |
|  | Microglial neuroinflammation |  |
|  | Inflammation | Wang X, Zhang XK, 2025. doi: 10.3390/biom15040480 |
|  | Significantly ↑ AD & FTD Plasma Proteomics | Ali M, et al, 2025. doi: 10.1038/s41591-025-03833-1  Erratum: 2025. doi: 10.1038/s41591-025-03970-7 |
| ***FLNA*** | pTau | Aumont E et al, 2022. doi: 10.3389/fnagi.2022.1038343 |
|  | Neuro-inflammation |  |
|  | Macrophage | Bandaru S et al, 2019. doi: 10.1161/CIRCULATIONAHA.119.039697 |
|  | Lipids |  |
|  | Cholesterol |  |
|  | TF | Loy CJ et al, 2003. doi: 10.1073/pnas.0736237100 |
|  | Significantly ↑ AD Plasma Proteomics | Ali M, et al, 2025. doi: 10.1038/s41591-025-03833-1  Erratum: 2025. doi: 10.1038/s41591-025-03970-7 |
| ***FOSB*** | TF | Corbett BF et al, 2017. doi: 10.1016/j.celrep.2017.06.040 |
|  | AD |  |
|  | Cognition |  |
|  | Inflammaging | Karakaslar EO et al, 2023. doi: 10.1111/acel.13792 |
|  | M1 Polarization | Fontana MF et al, 2015 Sep;37(9):470-8. doi: 10.1111/pim.12215 |
|  | Kinase cascades | Bhosale PB et al, 2022. doi: 10.1155/2022/9797929 |
|  | Obesity | White UA et al, 2010. doi: 10.1016/j.mce.2009.08.023 |
|  | T2D | Khan AW et al, 2024. doi: 10.2337/db23-0167 |
|  | Phosphoprotein | Ulery-Reynolds PG et al, 2009. doi: 10.1016/j.neuroscience.2008.10.059 |
|  | Chromatin | Yeh SY et al, 2023. doi: 10.1016/j.biopsych.2022.12.021 |
| ***FOXJ2*** | TF | Tan Y et al, 2024. doi: 10.1002/advs.202309140 |
|  | Inflammation |  |
|  | Kinase cascade |  |
|  | NFκB |  |
|  | Chromatin |  |
|  | Epigenetics |  |
|  | Monocytes |  |
| ***FOXO4*** | TF | Du S, Zheng H, 2021. doi: 10.1186/s13578-021-00700-7 |
|  | T2D |  |
|  | AD |  |
|  | Microglial neuroinflammation | Asadi Y et al, 2025. doi: https://doi.org/10.1101/2025.03.13.643180 |
|  | Cognition | Maiese K, 2017. doi: 10.2174/1567202614666171116102911 |
|  | Chromatin | Riedel CG et al, 2013. doi: 10.1038/ncb2720 |
|  | Significantly ↑ AD & PD Plasma Proteomics | Ali M, et al, 2025. doi: 10.1038/s41591-025-03833-1  Erratum: 2025. doi: 10.1038/s41591-025-03970-7 |
| ***FTO*** | Dementia | Keller L et al, 2011. doi: 10.3233/JAD-2010-101068  Benedict C et al, 2011. doi: 10.1016/j.neurobiolaging.2011.02.006 |
|  | Cognition |  |
|  | Obesity |  |
|  | M1 polarization | Hu F et al, 2019. doi: 10.1007/s00424-019-02316-w  Ren X et al, 2023. doi: 10.3389/fonc.2023.1241357 |
|  | Lipids | Yang Z et al, 2021. doi: 10.1016/j.gendis.2021.01.005 |
|  | Inflammation | Zhou C et al, 2023. doi: 10.1172/JCI160517 |
| ***FUT8*** | Microglial neuroinflammation | Jin LW et al, 2023. doi: 10.1002/glia.24345 |
| ***FXBL16*** | HIF1α | Kim YJ et al, 2021. doi: [http://dx.doi.org/10.1016/j.celrep.2021.109996](https://protect.checkpoint.com/v2/r01/___http:/dx.doi.org/10.1016/j.celrep.2021.109996___.YzJ1OmJpb3ZpZTE6YzpvOjFjZTBjMWI5N2VkOWI3NmJkYWM3N2E2ZjJlM2MxYmE4Ojc6NzJlMTpjNGRhM2I0Nzg5MzQ4MDY2M2E0ZGRmODE2ZTMxMTk3MWJiZTMxNGNiNjcyYTQwYjcyN2JmNzE1MjJkNTA4OWYzOnA6RjpG) |
|  | IRS1 | Morel M, Long W, 2023. doi: <https://doi.org/10.1002/1878-0261.13554> |
|  | Neuro-inflammation | Qu L et al, 2024. doi: <http://dx.doi.org/10.1186/s40364-024-00691-w> |
|  | NF-κB | Sugimoto-Ishige A et al, 2025. doi: https://doi.org/10.3389/fimmu.2025.1524110 |
|  | NOMINATED | Agora. n.d. [https://agora.adknowledgeportal.org/genes/ENSG00000127585](https://protect.checkpoint.com/v2/r01/___https:/agora.adknowledgeportal.org/genes/ENSG00000127585___.YzJ1OmJpb3ZpZTE6YzpvOjFjZTBjMWI5N2VkOWI3NmJkYWM3N2E2ZjJlM2MxYmE4Ojc6MTZiODozZDFhY2MyNDA3ODM5MjU2NTAyYTYxY2JhYTJjZDhmMTcyNjFhMjY5NmRjYzBhYjQwNTAzZDQ1NmI2ZWVhZGNkOnA6RjpG) |
| ***FYN***  *Kinase mediates neurotoxicity* | Aβ | Meur S et al, 2025. doi: 10.1007/s12035-024-04286-2 |
|  | pTau |  |
|  | Cognition | Guglietti B et al, 2021. doi: 10.1007/s12035-021-02518-3 |
|  | Microglial neuroinflammation | Panicker N et al, 2019. doi: 10.1084/jem.20182191 |
|  | Obesity | Lee TW et al, 2013. doi: 10.2337/db12-0920 |
|  | Triglycerides | Kazi JU, Rönnstrand L, 2019. doi. http://dx.doi.org/10.1016/j.biocel.2018.12.007 |
|  | T2D | Bastie CC et al, 2007. doi: 10.1016/j.cmet.2007.04.005 |
| ***GAB2*** | Microglial neuroinflammation | Kondreddy V et al, 2021. doi: 10.1161/ATVBAHA.121.316153  Kondreddy V et al, 2022. doi: 10.1182/blood.2022016424  Byeon JW et al, 2021. doi: 10.1016/j.bbrc.2021.06.028 |
|  | T2D | Wang X et al, 2021. doi: 10.1038/s41419-021-03519-9 |
|  | Obesity |  |
|  | Lipids |  |
|  | Phosphoproteins | Ding CB et al, 2015. doi: 10.3892/mmr.2015.3951 |
| ***GADD45B*** | PD | Ravel-Godreuil C et al, 2021. doi: 10.1016/j.isci.2021.102756 |
|  | Neuro-degeneration |  |
|  | Inflammation | Yang J et al, 2001. doi: 10.1038/84264 |
| ***GALNT6*** | Microglial neuroinflammation | Gurdon B et al, 2023. doi: 10.1101/2023.02.27.530226. Update, 2024. doi: 10.1038/s42003-024-06242-1 |
|  | Aβ |  |
|  | Cognition | Kato K et al, 2021. doi: 10.3390/molecules26185504 |
|  | Significantly ↑ AD microglial transcripts with Dementia | Kosoy R. et al, 2025. doi: 10.1038/s41593-025-02020-2 |
| ***GATA3*** | TF | Al-Mansoori L et al, 2020. doi: 10.1016/j.cellsig.2020.109735 |
|  | Inflammation |  |
|  | T2D |  |
|  | Obesity | El-Arabey AA et al, 2022. doi: 10.1016/j.cbi.2022.110141 |
|  | IR |  |
|  | M1 polarization |  |
|  | Kinase cascades | Yamashita M et al, 2005. doi: 10.1074/jbc.M502333200 |
|  | Phosphoprotein | Hosokawa H et al, 2016. doi: 10.1038/ncomms11289 |
|  | Chromatin | Swinstead EE et al, 2016. doi: 10.1002/bies.201600137 |
| ***GCA*** | Cognition | Zhou R et al, 2023. doi: 10.1002/advs.202303402  Liu ZT et al, 2024. doi: 10.1002/alz.13864 |
|  | T2D | Su T et al, 2024. doi: 10.1038/s41467-023-43787-x  Su T et al, 2024. doi: 10.1002/advs.202406500 |
|  | Inflammation |  |
|  | IR |  |
|  | Obesity |  |
|  | Kinase cascade |  |
|  | M1 polarization | Li CJ et al, 2021. doi: 10.1016/j.cmet.2021.08.009 |
| ***GDP2*** | T2D | OMIM, 2024. <https://omim.org/entry/138430> |
| ***GLUD1*** | Obesity | Vetterli L et al, 2016. doi: 10.1074/jbc.M115.707448 |
| ***GNA12*** | Neuro-inflammation | Galea E et al, 2022. doi: 10.1016/j.nbd.2022.105655 |
|  | Braak* score | Yang YM et al, 2020. doi: 10.1038/s12276-020-0454-5 |
|  | T2D |  |
|  | Obesity |  |
| ***GNA13*** | Microglial neuroinflammation | Bettegazzi B et al, 2021. doi: 10.1007/s12035-021-02553-0 |
|  | Inflammation | Rasheed SAK et al, 2018. doi: 10.1038/s41388-017-0038-6 |
|  | Kinase cascades |  |
|  | Lipids | Grimm M et al, 2016. doi: 10.1161/ATVBAHA.115.306066 |
|  | Macrophage |  |
|  | T2D | Yang YM et al, 2020. doi: 10.1038/s12276-020-0454-5 |
|  | Obesity | Lee Y, 2024 Abstract LB-R-29-01. FEBS Congress. [https://2024.febscongress.org/](https://protect.checkpoint.com/v2/r01/___https://2024.febscongress.org/___.YzJ1OmJpb3ZpZTE6YzpvOjljNzMyZTM5MjY4ZDg1MDA1MDcyODAxYjc4YTlmZjRhOjc6ZGNjNDo1NTljZDUyZjY4MmNhMzg3ZGQ3M2FhOTc4ODI5MTYzMDVkMzU1OGRjNDE1YTU2ZjU2MTcwZDQ0OWI3N2E4ZGU4OnA6RjpG)  Tokgozoglu J et al, 2025. doi: 10.1038/s41419-025-08004-1 |
| ***GOT2*** | Glutamate | Bukke VN et al, 2020. doi: 10.3390/ijms21207452  Wang R, Reddy PH, 2017. doi: 10.3233/JAD-160763  Holton KF et al, 2021. doi: 10.3389/fnins.2021.726457  Zhou XH et al, 2022. doi: 10.1515/tnsci-2022-0220  Castillo-Vazquez SK et al, 2024. doi: 10.1016/j.arcmed.2024.103039 |
|  | Neuro-inflammation |  |
|  | Cognition |  |
| ***GPAT3*** | Microglial neuroinflammation | Fan G et al, 2023. doi: 10.1038/s41419-023-05741-z  Haney MS et al, 2023. doi: 10.1101/2023.07.21.549930. Update, 2024. doi: 10.1038/s41586-024-07185-7 |
|  | Lipids |  |
|  | ERK |  |
|  | Obesity | Fan G et al, 2024. doi: 10.1016/j.bbadis.2023.167007  Ma S et al, 2015. doi: 10.1038/srep07633  Kuhajda FP et al, 2011. doi: 10.1152/ajpregu.00147.2011 |
|  | Cholesterol | Yu J et al, 2018. doi: 10.1038/s41387-018-0045-x |
|  | T2D |  |
|  | M1 polarization | Quiroga IY et al, 2021. doi: 10.1016/j.atherosclerosis.2020.11.022 |
| ***GPCPD1*** | Aging | Cikes D et al, 2024. doi: 10.1038/s43587-023-00551-6. |
|  | T2D |  |
|  | Lipids |  |
| ***GPD2*** | Microglial neuroinflammation | You JE et al, 2024. doi: 10.3390/biomedicines12051130  Langston PK et al, 2019. doi: 10.1038/s41590-019-0453-7 |
|  | M1 polarization |  |
|  | T2D | Omim, 2024. [https://omim.org/entry/138430](https://protect.checkpoint.com/v2/r01/___https://omim.org/entry/138430___.YzJ1OmJpb3ZpZTE6YzpvOjljNzMyZTM5MjY4ZDg1MDA1MDcyODAxYjc4YTlmZjRhOjc6YjEwYTplZGY0Y2RiMTQ0ZmU5NDRhMzE5MjI5NjRiZGE3OTQ4NDIxMTA5MDY4NjA1ZmQzOWU2ZWE5ZjJiNDgyYjhmNzczOnA6RjpG)  Qu H et al, 2021. doi: 10.2337/db20-1157 |
|  | Glycolysis | Lu J et al, 2020. doi: 10.1111/cas.14408  Oh S et al, 2024. doi: 10.1038/s12276-024-01222-1 |
|  | Lipids | Wu S et al, 2022. doi: 10.1073/pnas.2121987119  Oh S et al, 2023. doi: 10.7150/thno.75973 |
|  | Ferroptosis |  |
| ***GTF2I*** | TF | Kim DW et al, 2000. doi: 10.1128/MCB.20.4.1140-1148.2000 |
|  | Kinase cascades |  |
|  | ERK |  |
|  | Phosphoprotein | NIH, 2025. [https://www.ncbi.nlm.nih.gov/gene/2969](https://protect.checkpoint.com/v2/r01/___https://www.ncbi.nlm.nih.gov/gene/2969___.YzJ1OmJpb3ZpZTE6YzpvOjljNzMyZTM5MjY4ZDg1MDA1MDcyODAxYjc4YTlmZjRhOjc6ZDk0Mjo3YWQ3YWFjYTQ5N2Q4YmIwY2Q5NTJjNDEyNmU2MjNkODYxNWY0OWRlNzFkMWYzYTMyODBiMDUwYTVjNmJkYWI5OnA6RjpG) |
|  | Significantly ↑ AD Plasma Proteomics | Ali M, et al, 2025. doi: 10.1038/s41591-025-03833-1  Erratum: 2025. doi: 10.1038/s41591-025-03970-7 |
| ***GTF3C2*** | Vascular inflammation | Sun W et al, 2024. doi: 10.1007/s00018-024-05282-4 |
|  | TF | NIH, 2025. [https://www.ncbi.nlm.nih.gov/gene/2976#:~:text=Summary,the%20specific%20induction%20of%20TFIIIC110](https://protect.checkpoint.com/v2/r01/___https://www.ncbi.nlm.nih.gov/gene/2976___.YzJ1OmJpb3ZpZTE6YzpvOjljNzMyZTM5MjY4ZDg1MDA1MDcyODAxYjc4YTlmZjRhOjc6MDBmNTpkMzI3YjA2Yzc2N2RlZmFkZTZkMDQ5ZjVjYzEwNjgyZGIyMGNmNjcxYmU3NGJlOTJkYzJhZTVjYjA2NjY4YTAxOnA6RjpG" \l ":~:text=Summary,the%20specific%20induction%20of%20TFIIIC110) |
|  | Chromatin | Kundu TK et al, 1999. doi: 10.1128/MCB.19.2.1605 |
| ***H1-10*** | Microglial neuroinflammation | Gilthorpe JD et al, 2013. doi: 10.12688/f1000research.2-148.v1  Tarcic O et al, 2016. doi: 10.1016/j.celrep.2016.01.020 |
|  | Nuclear histone | Sollberger G et al, 2020. doi: 10.7554/eLife.52563 |
|  | Significantly ↑ AD & PD Plasma Proteomics | Ali M, et al, 2025. doi: 10.1038/s41591-025-03833-1  Erratum: 2025. doi: 10.1038/s41591-025-03970-7 |
| ***H2BC21*** | Microglial neuroinflammation | Zheng SZ et al, 2022. doi: 10.1016/j.phrs.2022.106093 |
|  | Histone expression | Jia J et al, 2022. doi: 10.3389/fonc.2022.966817 |
|  | T2D | Luo R et al, 2025. doi: 10.1371/journal.pone.0320061 |
|  | Periodontitis |  |
|  | Significantly ↑ AD & PD Plasma Proteomics | Ali M, et al, 2025. doi: 10.1038/s41591-025-03833-1  Erratum: 2025. doi: 10.1038/s41591-025-03970-7 |
| ***H2BC5*** | Histone-associated inflammation | Jia J et al, 2022. doi: 10.3389/fonc.2022.966817 |
| ***H2BC9*** | Histone-associated inflammation | Jia J et al, 2022. doi: 10.3389/fonc.2022.966817 |
| ***H3-3B*** | Vascular inflammation | Zhang X et al, 2023. doi: 10.1016/j.ygeno.2023.110685 |
|  | Phosphoprotein | Armache A et al, 2020. doi: 10.1038/s41586-020-2533-0 |
|  | Kinase cascades |  |
|  | Chromatin | Masuzawa R et al, 2024. doi: 10.1186/s12576-024-00935-2 |
| ***HASPIN*** | Neuro-inflammation | Tanaka H et al, 2023. doi: 10.3390/biology12020320  Melms JC et al, 2020. doi: 10.1158/0008-5472.CAN-19-2330 |
|  | pTau |  |
|  | Cognition |  |
|  | Kinase cascades | Ghenoiu C et al, 2013. doi: 10.1016/j.molcel.2013.10.002 |
|  | Phosphoprotein |  |
|  | Chromatin |  |
| ***HCFC1*** | Inflammation | Wang H et al, 2023. doi: 10.7150/jca.8457 |
|  | Chromatin | Vogel JL, Kristie TM, 2013. doi: 10.3390/v5051272 |
|  | TF | NIH, 2025. [https://www.ncbi.nlm.nih.gov/gene/3054](https://protect.checkpoint.com/v2/r01/___https://www.ncbi.nlm.nih.gov/gene/3054___.YzJ1OmJpb3ZpZTE6YzpvOjljNzMyZTM5MjY4ZDg1MDA1MDcyODAxYjc4YTlmZjRhOjc6ODYyNjo5MzIxNTAxMjkwZTgwNzIxODc1ZTE5YzY5NzgyYzQ0NjViZTAxMmYyODg0MTYzODJhN2I0YzQ0NWE3NzI2MmZlOnA6RjpG) |
| ***HDAC11*** | Microglial neuroinflammation | Bai P et al, 2025. doi: 10.1002/alz.14616 |
|  | Aβ |  |
|  | Cognition |  |
|  | Lipids | Cao J et al, 2019. doi: 10.1073/pnas.1815365116  Núñez-Álvarez Y et al, 2022. doi: 10.1111/febs.15895  Bai P et al, 2025. doi: 10.1002/alz.14616 |
|  | T2D | Sun L et al, 2018. doi: 10.1016/j.ebiom.2018.06.025 |
|  | Obesity |  |
|  | Cholesterol |  |
|  | MAPK | Zhou Z et al, 2023. doi: 10.1007/s12032-023-02196-2 |
|  | Phosphoprotein | Bahl S, Seto E, 2021. doi: 10.1007/s00018-020-03599-4 |
|  | T2D | Chen H et al, 2022. doi: 10.3389/fendo.2022.989305 |
|  | Glycolysis | Bi L et al, 2021. doi: 10.1158/0008-5472.CAN-20-3044 |
| ***HDAC4*** | Cognition | Colussi C et al, 2023. doi: 10.1111/nan.12861  Wu Y et al, 2016. doi: 10.3389/fnmol.2016.00114  Main P et al, 2021. doi: 10.3389/fnmol.2021.616642. |
|  | Microglial neuroinflammation | Jayaraj K et al, 2025. doi: 10.1002/glia.70035  Wang B et al, 2014. doi: 10.1091/mbc.E13-12-0757 |
|  | T2D | Lin Y et al, 2025. doi: 10.1186/s40001-025-02697-y |
|  | Obesity | Ozcan L et al, 2022. doi: 10.1016/j.celrep.2022.111015. Erratum, 2016. doi: 10.1016/j.celrep.2016.05.006 |
|  | Glycolysis | Wang B et al, 2014. doi: 10.1091/mbc.E13-12-0757 |
|  | M1 polarization | Ding Y et al, 2025. doi: 10.3389/fphar.2025.1515787 |
|  | Chromatin | Wang Z et al, 2014. doi: 10.2217/epi.13.73 |
|  | Significantly ↑ AD & PD Plasma Proteomics | Ali M, et al, 2025. doi: 10.1038/s41591-025-03833-1  Erratum: 2025. doi: 10.1038/s41591-025-03970-7 |
| ***HEBP2*** | Microglial neuroinflammation | Block ML et al, 2007. doi: 10.1038/nrn2038  GeneCards, 2025. [https://www.genecards.org/cgi-bin/carddisp.pl?gene=HEBP2](https://protect.checkpoint.com/v2/r01/___https://www.genecards.org/cgi-bin/carddisp.pl?gene=HEBP2___.YzJ1OmJpb3ZpZTE6YzpvOjljNzMyZTM5MjY4ZDg1MDA1MDcyODAxYjc4YTlmZjRhOjc6MDZiNjo4N2VlMmI4MWY0MWEwMjZlYTU0NzVlNGMzM2U2ODhiYjA4NGExZjllZWU1MGIxODhmMWE3NTg2Nzk3NWE2NTYwOnA6RjpG) |
|  | Cognition | Yagensky O et al, 2019. doi: 10.7554/eLife.47498 |
|  | T2D | Takematsu E et al, 2020. doi: 10.1371/journal.pone.0225267 |
|  | Lipids | Yang B, Dai M, 2024. doi: 10.1016/j.tranon.2024.101917 |
|  | NOMINATED** | Agora. [https://agora.adknowledgeportal.org/genes/ENSG00000051620](https://protect.checkpoint.com/v2/r01/___https://agora.adknowledgeportal.org/genes/ENSG00000051620___.YzJ1OmJpb3ZpZTE6YzpvOjljNzMyZTM5MjY4ZDg1MDA1MDcyODAxYjc4YTlmZjRhOjc6NjUwZjpiOGI2NzRiZGEwNTBhZTYwZjBhNDgzODc1ZDA4N2Y1NDRkODU4ZmQ2NWI3MTFkNGUyODMxNjlhYmQzZGJkMTY3OnA6RjpG) |
| ***HEMK1*** | Aβ | Yagensky O et al, 2019. doi: 10.7554/eLife.47498 |
|  | Hebp1 |  |
|  | T2D | Ishihara S et al, 1988. doi: 10.1111/j.1447-0756.1988.tb00081.x |
| ***HEPACAM2*** | Inflammation | Wang S et al, 2025. doi: 10.2174/0115665232325395241018103006 |
|  | Significantly ↑ AD Plasma Proteomics | Ali M, et al, 2025. doi: 10.1038/s41591-025-03833-1  Erratum: 2025. doi: 10.1038/s41591-025-03970-7 |
| ***HIRA*** | Chromatin | Esteves de Lima J et al, 2021. doi: 10.1038/s41467-021-23775-9 |
|  | Inflammation | Dasgupta N et al, 2024. doi: 10.1016/j.molcel.2024.08.006 |
|  | Obesity | Wan D et al, 2025. doi: 10.1101/2025.03.21.644577 |
|  | Phosphoprotein |  |
| ***HK2*** | Microglial neuroinflammation | Grantome, 2024. [https://grantome.com/grant/NIH/RF1-AG068400-01#:~:text=The%20initial%20and%20rate%20limiting,intervention%20strategies%20attenuate%20disease%20pathogenesis](https://protect.checkpoint.com/v2/r01/___https://grantome.com/grant/NIH/RF1-AG068400-01___.YzJ1OmJpb3ZpZTE6YzpvOjljNzMyZTM5MjY4ZDg1MDA1MDcyODAxYjc4YTlmZjRhOjc6ZmY3Mjo1NTI2ZTY2ZjQ5YTVmMGZiYjkyNzYxYjVhOWMyNjYxNDliZDljMWFhZTljYjU5M2ZmYzdjMjRlYzczMGRhODU0OnA6RjpG#:~:text=The%20initial%20and%20rate%20limiting,intervention%20strategies%20attenuate%20disease%20pathogenesis) |
|  | Glycolysis |  |
|  | Cognition | Codocedo JF et al, 2024. doi: 10.1016/j.celrep.2024.114488 |
|  | T2D | Rabbani N et al, 2024. doi: 10.3389/fendo.2023.1268308  Rabbani N, Thornalley PJ, 2019. doi: 10.1016/j.tem.2019.04.011 |
|  | M1 polarization | Li J et al, 2025. doi: 10.1016/j.celrep.2025.115350 |
|  | Significantly ↑ AD Plasma Proteomics | Ali M, et al, 2025. doi: 10.1038/s41591-025-03833-1  Erratum: 2025. doi: 10.1038/s41591-025-03970-7 |
| ***HMGB1*** | TF | Paudel YN et al, 2020. doi: 10.3390/cells9020383  Gaikwad S et al, 2021. doi: 10.1016/j.celrep.2021.109419 |
|  | Microglial neuroinflammation |  |
|  | Neuropathology |  |
|  | Cognition | Ngadimon IW et al, 2025. doi: 10.1002/epi4.70001 |
|  | T2D | Zhu Z et al, 2020. doi: 10.1096/fj.202000242R  Dash UK et al, 2024. doi: 10.1007/s12035-024-04081-z |
|  | Obesity | Zhang J et al, 2017. doi: 10.1016/j.mce.2017.06.012  Arrigo T et al, 2013. doi: 10.1530/EJE-13-003  Chen L et al, 2020. doi: 10.1161/ATVBAHA.120.314599 |
|  | M1 polarization | Hu C et al, 2025. doi: 10.1016/j.intimp.2025.114192  Li YF et al, 2022. doi: 10.3389/fnagi.2022.901117  Mo J et al, 2023. doi: 10.1016/j.biopha.2023.114541 |
|  | Glycolysis | Chen R et al, 2018. doi: 10.1002/hep.29663 |
|  | Lipids | Montes VN et al, 2015. doi: 10.1038/nutd.2015.11 |
|  | Chromatin | Joshi SR et al, 2012. doi: 10.1093/nar/gks815 |
| ***HMGB2*** | TF | Lee S et al, 2014. doi: 10.1038/nchembio.1669  UPenn, 2025. [https://www.med.upenn.edu/aging/the-science-of-aging-summer-2025.html#:~:text=HMGB2%2C%20which%20may%20play%20a%20key%20role,HMGB2%20levels%20are%20much%20higher%20and%20can](https://protect.checkpoint.com/v2/r01/___https://www.med.upenn.edu/aging/the-science-of-aging-summer-2025.html___.YzJ1OmJpb3ZpZTE6YzpvOjljNzMyZTM5MjY4ZDg1MDA1MDcyODAxYjc4YTlmZjRhOjc6Yjc2OTo5MTE5N2ZhZmQ1MWRkYjM0YmY3NjZlMjQ4MDM2OTc2Y2FmNmI2NjJlMjZjYjA5NTIzMzEzNmNjNDVlYWE3YWUyOnA6RjpG#:~:text=HMGB2%2C%20which%20may%20play%20a%20key%20role,HMGB2%20levels%20are%20much%20higher%20and%20can) |
|  | Microglial neuroinflammation |  |
|  | Ferroptosis | Blair I et al, 2024. doi: 10.21203/rs.3.rs-4009459/v2. Update, 2024. doi: 10.1038/s42003-024-06930-y |
|  | Inflammation | Wan J et al, 2022. doi: 10.1177/20406223221135011 |
|  | Hypertension |  |
|  | Retinal degeneration | Zhang Y et al, 2022. doi: 10.1016/j.freeradbiomed.2022.01.018 |
|  | Glycolysis | Fu D et al, 2018. doi: 10.1186/s12964-018-0219-0 |
|  | Obesity | Morinaga H et al, 2021. doi: 10.1016/j.bbrc.2021.03.149  Choijookhuu N et al, 2024. doi: 10.1369/00221554241241569 |
|  | Lipids |  |
|  | Chromatin | Aird KM et al, 2016. doi: 10.1083/jcb.201608026. |
|  | Significantly ↑ AD Plasma Proteomics | Ali M, et al, 2025. doi: 10.1038/s41591-025-03833-1  Erratum: 2025. doi: 10.1038/s41591-025-03970-7 |
| ***HMGN1*** | Inflammation | Yang D et al, 2012. doi: 10.1084/jem.20101354 |
|  | Microglial neuroinflammation | Gao HM et al, 2011. doi: 10.1523/JNEUROSCI.3732-10.2011 |
|  | M1 polarization | Mu Q et al, 2025. doi: 10.1002/cbin.12252 |
|  | Obesity | Nanduri R et al, 2022. doi: 10.1038/s41467-022-34964-5 |
|  | Cholesterol | Postnikov YV et al, 2014. doi: 10.1158/1541-7786.MCR-13-0392 |
|  | Lipids |  |
|  | TF | Murphy KJ et al, 2017. doi: 10.1093/nar/gkx579 |
|  | Chromatin |  |
|  | Significantly ↑ AD Plasma Proteomics | Ali M, et al, 2025. doi: 10.1038/s41591-025-03833-1  Erratum: 2025. doi: 10.1038/s41591-025-03970-7 |
| ***HNRNPA2B1*** | Cognition | Xia T et al, 2022. doi: 10.3389/fnagi.2022.1034041  Wang J et al, 2020. doi: 10.3389/fnagi.2020.00086 |
|  | Inflammation | Meng M et al, 2023. doi: 10.3390/nu15071555 |
|  | M1 polarization |  |
|  | Obesity |  |
|  | Glycolysis | Yin M et al, 2021. doi: 10.1016/j.isci.2021.103345 |
|  | TF | Nguyen ED et al, 2018. doi: 10.1080/15476286.2018.1474072 |
|  | Chromatin |  |
|  | Significantly ↑ AD Plasma Proteomics | Ali M, et al, 2025. doi: 10.1038/s41591-025-03833-1  Erratum: 2025. doi: 10.1038/s41591-025-03970-7 |
| ***HNRNPK*** | Cognition | Sidhu R et al, 2022. doi: 10.1111/nan.12793  Kavanagh T et al, 2025. doi: 10.1111/bpa.13305  MedlinePlus, 2019. [https://medlineplus.gov/genetics/gene/hnrnpk/#:~:text=The%20HNRNPK%20gene%20provides%20instructions,critical%20for%20learning%20and%20memory](https://protect.checkpoint.com/v2/r01/___https://medlineplus.gov/genetics/gene/hnrnpk/___.YzJ1OmJpb3ZpZTE6YzpvOjljNzMyZTM5MjY4ZDg1MDA1MDcyODAxYjc4YTlmZjRhOjc6OWMxOTpjZjczNDk0NjgxZGRmNThiYTY2YWJiOWFjZGRiYzYzN2MyYjgzMTk0MWJhNDJjMWUwYjU5ZGZlOTlkMmEwYTNmOnA6RjpG#:~:text=The%20HNRNPK%20gene%20provides%20instructions,critical%20for%20learning%20and%20memory) |
|  | Inflammation | Liu S et al, 2023. doi: 10.1177/15353702221110649  Feng J et al, 2020. doi: 10.1080/13510002.2020.1857157 |
|  | T2D | Chen X et al, 2022. doi: 10.1016/j.molmet.2022.101515 |
|  | Lipids | Zhang M et al, 2025. doi: 10.1016/j.jbc.2025.110500 |
|  | Obesity |  |
|  | Glycolysis | Sun Z et al, 2018. doi: 10.1016/j.stemcr.2018.08.001 |
|  | TF | Li H et al, 2010. doi: 10.1074/jbc.M109.082057 |
|  | Chromatin |  |
| ***HOXA4*** | Inflammation | Pai P, Sukumar S, 2020. doi: 10.1016/j.bbcan.2020.188450 |
|  | Atherosclerosis | Zhou Y et al, 2023. doi: 10.3892/ijmm.2023.5317 |
|  | Cognition | Li QS et al, 2021. doi: 10.1186/s13148-021-01179-2 |
|  | TF | Mazzoni EO et al, 2013. doi: 10.1038/nn.3490 |
|  | Chromatin |  |
| ***HSPA1A*** | Inflammation | Chen Y et al, 2007. doi: 10.2174/187152807780832274  Ogbodo E et al, 2023. doi: 10.1002/2211-5463.13695 |
|  | M1 polarization |  |
|  | AD | Wang Y et al, 2024. doi: 10.3390/ijms25136934  Dong Y et al, 2022. doi: 10.1155/2022/9480398 |
|  | Cognition |  |
|  | T2D | Ochoa Mendoza V et al, 2025. doi: 10.3390/cells14060424 |
|  | Obesity | Timofeev YS et al, 2023. doi: 10.3390/cimb45120588 |
|  | TLR | Klink M et al, 2012. doi: 10.1007/s12192-012-0338-2 |
|  | Lipids | Zhang J et al, 2018. doi: 10.1186/s12944-018-0722-8 |
| ***HSPA1B*** | Inflammation | Gu Y et al, 2025. doi: 10.3390/brainsci15020205 |
|  | Cognition |  |
|  | M1 polarization | Chen Y et al, 2007. doi: 10.2174/187152807780832274  Ogbodo E et al, 2023. doi: 10.1002/2211-5463.13695 |
|  | T2D | Ochoa Mendoza V et al, 2025. doi: 10.3390/cells14060424 |
|  | Obesity | Timofeev YS et al, 2023. doi: 10.3390/cimb45120588  Ruppert Z et al, 2025. doi: 10.1186/s13293-025-00746-z |
|  | Lipids | Zhang J et al, 2018. doi: 10.1186/s12944-018-0722-8 |
| ***HSPA2*** | AD | Petyuk VA et al, 2018. doi: 10.1093/brain/awy215 |
|  | Aβ |  |
|  | pTau |  |
|  | Inflammation | Dong Y et al, 2022. doi: 10.1155/2022/9480398 |
|  | Obesity | Zhang W et al, 2022. doi: 10.3389/fendo.2022.1041027 |
|  | Lipids |  |
|  | DM | Conning-Rowland MS et al, 2024. doi: 10.1093/cvr/cvae181 |
|  | CVD |  |
|  | Significantly ↑ AD Plasma Proteomics | Ali M, et al, 2025. doi: 10.1038/s41591-025-03833-1  Erratum: 2025. doi: 10.1038/s41591-025-03970-7 |
|  | NOMINATED** | Agora. [https://agora.adknowledgeportal.org/genes/ENSG00000126803](https://protect.checkpoint.com/v2/r01/___https://agora.adknowledgeportal.org/genes/ENSG00000126803___.YzJ1OmJpb3ZpZTE6YzpvOjljNzMyZTM5MjY4ZDg1MDA1MDcyODAxYjc4YTlmZjRhOjc6MjViNjoyZTQ4MjQyMTdiOWY3MzQwZDFkMzUxMWU1NTM0NDJmNDY5NDdmZGFiOGE0ZGI2YzYxODBjMzZhYzNmZGMyOTU4OnA6RjpG) |
| ***HSPBAP1*** | Microglial neuroinflammation | Dukay B et al, 2021. doi: 10.1186/s12974-020-02070-2 |
|  | T2D | Atalay M et al, 2009. doi: 10.2174/138920309787315202 |
|  | M1 polarization | Essalemi RK et al, 2025. doi:10.33696/cancerimmunol.7.104 |
|  | *HSPB1* association | NIH, 2025. [https://www.ncbi.nlm.nih.gov/gene/79663#:~:text=This%20gene%20encodes%20a%20protein%20that%20binds,is%20involved%20with%20cell%20growth%20and%20](https://protect.checkpoint.com/v2/r01/___https://www.ncbi.nlm.nih.gov/gene/79663___.YzJ1OmJpb3ZpZTE6YzpvOjljNzMyZTM5MjY4ZDg1MDA1MDcyODAxYjc4YTlmZjRhOjc6YzcxNzpiMjEyNDZmNDhlZWZiYTJmMWUwNWIwNGRjZWM3ZTg4YjM3N2MxNTU4NmZmM2FmYTYyNzE0MTNhMjY4NTViZjlkOnA6RjpG" \l ":~:text=This%20gene%20encodes%20a%20protein%20that%20binds,is%20involved%20with%20cell%20growth%20and%20) |
| ***HTRA2*** | Inflammation | Liu Q et al, 2024. doi: 10.3390/ijms25031577  Xu Z et al, 2021. doi: 10.1016/j.molimm.2020.10.024 |
| ***IFIH1*** | Inflammation | Zhang S et al, 2024. doi: 10.1002/ctm2.70027 |
|  | M1 polarization |  |
|  | Cognition | Guney MH et al, 2023. doi: 10.1101/2023.11.17.567619. Update, 2024. doi: 10.1073/pnas.2404349121 |
|  | AD | Neher JJ, 2022. doi: 10.1016/j.immuni.2022.04.008 |
|  | Microglial neuroinflammation |  |
|  | Kinase cascades | Lee NR et al, 2015. doi: 10.14348/molcells.2015.0047 |
|  | NOMINATED** | Agora. [https://agora.adknowledgeportal.org/genes/ENSG00000115267](https://protect.checkpoint.com/v2/r01/___https://agora.adknowledgeportal.org/genes/ENSG00000115267___.YzJ1OmJpb3ZpZTE6YzpvOjljNzMyZTM5MjY4ZDg1MDA1MDcyODAxYjc4YTlmZjRhOjc6ZmRiNToxODA4NzFlMjFjZTc1MzQzMzQyNzVkZDVkMWFiZDA2OGQ0NzNkZmVlNDU5OGZhNzg1MjE1Zjg5NjliMDlkYWNmOnA6RjpG) |
| ***IGF2BP3*** | AD | Miao J et al, 2025. doi: 10.1007/s12035-024-04457-1 |
|  | pTau |  |
|  | Cognition |  |
|  | Kinase activation | Suvasini R et al, 2011. doi: 10.1074/jbc.M110.178012 |
|  | Brain diseases | Alberini CM, 2023. doi: 10.1016/j.tins.2023.03.007 |
|  | Inflammation | lcheva IA et al, 2023. doi: 10.3389/fimmu.2023.1224516  Cheng K et al, 2024. doi: 10.1016/j.intimp.2024.113030 |
|  | Glycolysis | Wang B et al, 2024. doi: 10.1038/s41419-024-06564-2 |
|  | T2D | Wu X et al, 2023. doi: 10.1111/1753-0407.13378  Rattanapan Y et al, 2024. doi: 10.14740/jocmr6099 |
| ***IKBKB*** | Cognition | Schnöder L et al, 2023. doi: 10.1096/fj.202201512R  Sun E et al, 2022. doi: 10.3390/ijms23168972  Liu Y et al, 2014. doi: 10.1523/JNEUROSCI.1348-14.2014  Yang S et al, 2021. doi: 10.3390/cells10102669 |
|  | Microglial neuroinflammation |  |
|  | M1 polarization |  |
|  | pTau |  |
|  | Aβ |  |
|  | Kinase cascade |  |
|  | Glycolysis | Kawauchi K et al, 2008. doi: 10.1038/ncb1724 |
|  | Obesity | Wunderlich FT et al, 2008. doi: 10.1073/pnas.0707849104 |
|  | NOMINATED** | Agora. [https://agora.adknowledgeportal.org/genes/ENSG00000104365](https://protect.checkpoint.com/v2/r01/___https://agora.adknowledgeportal.org/genes/ENSG00000104365___.YzJ1OmJpb3ZpZTE6YzpvOjljNzMyZTM5MjY4ZDg1MDA1MDcyODAxYjc4YTlmZjRhOjc6MWY0NzplZTU0MDc5N2EyYzZlZTAzOTc4ZTUzNjAwYzU4ZTRhYWYzODg2NmVhZjMwZmExMjc0MTk3NGFmMjQyNmRjYTU1OnA6RjpG) |
| ***IKBKG*** | Microglial neuroinflammation | Maqbool A et al, 2013. 10.1186/1750-1326-8-40 |
|  | Kinase cascade |  |
|  | Cognition |  |
|  | Glycolysis | Londhe P et al, 2018. doi: 10.3389/fonc.2018.00104 |
|  | M1 polarization | Wang N et al, 2014. doi: 10.3389/fimmu.2014.00614  Alanazi FJ et al, 2025. doi: 10.1016/j.prp.2025.155903 |
|  | T2D | Kracht M et al, 2020. doi: 10.1016/j.jaci.2020.07.027 |
|  | Obesity | Wunderlich FT et al, 2008. doi: 10.1073/pnas.0707849104 |
| ***IKZF1*** | Inflammation | Oh KS et al, 2018. doi: 10.4049/jimmunol.1800158  Ballasch I et al, 2023. doi: 10.1016/j.bbi.2023.01.016 |
|  | Microglial neuroinflammation |  |
|  | AD |  |
|  | M1 polarization | Liu G et al, 2025. doi: 10.1016/j.bbadis.2025.167690 |
|  | Chromatin | Ding Y et al, 2025. doi: 10.1038/s41375-025-02651-  NIH, 2025. [https://www.ncbi.nlm.nih.gov/gene/10320](https://protect.checkpoint.com/v2/r01/___https://www.ncbi.nlm.nih.gov/gene/10320___.YzJ1OmJpb3ZpZTE6YzpvOjljNzMyZTM5MjY4ZDg1MDA1MDcyODAxYjc4YTlmZjRhOjc6NzgyYTo3NzU0YzJjM2JiY2Q2NmEwYjYzZTc1N2YwYTRhOTE1MWY5ODBlMWNhM2E5NzVhY2JlNzYxMTI5ODViZDk3MmZhOnA6RjpG) |
|  | HDAC1 |  |
|  | TF |  |
| ***IL12RB1*** | Inflammation | Ford NR et al, 2012. doi: 10.4049/jimmunol.1200606 |
|  | T2D | Taylor-Fishwick DA et al, 2013. doi: 10.1007/s00125-012-2732-9 |
|  | AD | Schneeberger S et al, 2025. doi: 10.1038/s43587-025-00816-2  Medical Xpress, 2025. file:///C:/Users/KristinDeBellis/Downloads/2025-03-microglia-inflammatory-shift-il-alzheimer.pdf |
|  | Neuro-inflammation |  |
| ***IL15*** | Inflammation | Waldmann TA, 2004. doi: 10.1186/ar1202 |
|  | Cognition | Di Castro MA et al, 2024. doi: 10.1016/j.bbi.2023.11.015 |
|  | Significantly ↑ AD microglial transcripts with Dementia | Kosoy R. et al, 2025. doi: 10.1038/s41593-025-02020-2 |
|  | Braak* stage |  |
|  | Significantly ↑ AD Plasma Proteomics | Ali M, et al, 2025. doi: 10.1038/s41591-025-03833-1 Erratum: 2025. doi: 10.1038/s41591-025-03970-7 |
|  | NOMINATED** | Agora. [https://agora.adknowledgeportal.org/genes/ENSG00000164136](https://protect.checkpoint.com/v2/r01/___https://agora.adknowledgeportal.org/genes/ENSG00000164136___.YzJ1OmJpb3ZpZTE6YzpvOjljNzMyZTM5MjY4ZDg1MDA1MDcyODAxYjc4YTlmZjRhOjc6MjJhYjoxOTA4MWY0MTE3MWJjYTBlMmE1NjM4ZWUxNmZlZDA4MmIzYjA1N2NlY2FiZDZjMzc3NTFiZWRmNzYxZDgwZWQ0OnA6RjpG) |
| ***IL6R*** | Microglial neuroinflammation | Lyra E Silva NM et al, 2021. doi: 10.1038/s41398-021-01349-z |
|  | Cognition |  |
|  | M1 polarization | Chen X et al, 2022. doi: 10.1590/1678-7757-2022-0316 |
|  | T2D | Rehman K et al, 2017. doi: 10.1615/CritRevEukaryotGeneExpr.2017019712 |
|  | Obesity | Sindhu S et al, 2015. doi: 10.1371/journal.pone.0133494 |
|  | Lipids |  |
| ***IMPDH2*** | Microglial neuroinflammation | Liao LX et al, 2017. doi: 10.1073/pnas.1706778114 |
|  | Obesity | Hu J et al, 2024. doi: 10.1016/j.bbrc.2024.149998 |
|  | Significantly ↑ AD Plasma Proteomics | Ali M, et al, 2025. doi: 10.1038/s41591-025-03833-1 Erratum: 2025. doi: 10.1038/s41591-025-03970-7 |
| ***IRF1*** | Microglial neuroinflammation | Zegarra-Valdivia J et al, 2025. doi: 10.3390/cimb47040233  Fan X et al, 2025. doi: 10.3390/ijms26072906 |
|  | Cognition |  |
|  | M1 polarization | Chu YB et al, 2021. doi: 10.1016/j.intimp.2021.108072 |
|  | T2D | Zhou Y et al, 2025. doi: 10.1016/j.avsg.2025.02.018 |
|  | Obesity | Friesen M et al, 2017. doi: 10.1016/j.stemcr.2017.03.014 |
|  | Lipids | Friesen M et al, 2017. doi: 10.1016/j.stemcr.2017.03.014  Zhang F et al, 2025. doi: 10.1186/s40164-025-00612-z |
|  | TF | Feng H et al, 2021. doi: 10.1371/journal.ppat.1009220 |
|  | Chromatin | Song R et al, 2021. doi: 10.1016/j.celrep.2021.108891 |
|  | Significantly ↑ AD & PD Plasma Proteomics | Ali M, et al, 2025. doi: 10.1038/s41591-025-03833-1 Erratum: 2025. doi: 10.1038/s41591-025-03970-7 |
| ***IRF3I*** | Microglial neuroinflammation | Herzog AR, Wallace RB, 1997. doi: 10.1093/geronb/52b.special_issue.37 |
|  | M1 polarization | Wang N et al, 2014. doi: 10.3389/fimmu.2014.00614 |
|  | Glycolysis | Yan S et al, 2021. doi: 10.1172/JCI144888 |
|  | Cognition | Neupane C et al, 2024. doi: 10.1523/JNEUROSCI.1810-23.2024. Erratum: doi: 10.1523/JNEUROSCI.2192-24.2024 |
|  | T2D | Hu HQ et al, 2020. doi: 10.1016/j.mce.2020.110890 |
|  | Lipids |  |
|  | Obesity | Kumari M et al, 2016. doi: 10.1172/JCI86080 |
|  | Chromatin | Song R et al, 2021. doi: 10.1016/j.celrep.2021.108891 |
| ***IRF5*** | Microglial neuroinflammation | Yu X et al, 2025. doi: 10.3389/fimmu.2025.1535823 |
|  | Aβ |  |
|  | M1 polarization | Krausgruber T et al, 2011. doi: 10.1038/ni.1990 |
|  | Glycolysis | Fang Y et al, 2023. doi: 10.1111/1751-2980.13218 |
|  | Obesity | Dalmas E et al, 2015. doi: 10.1038/nm.3829 |
|  | TD |  |
|  | Kinase cascades | Ren J et al, 2014. doi: 10.1073/pnas.1418516111 |
|  | Phosphoprotein |  |
|  | TF | Yu X et al, 2025. doi: 10.3389/fimmu.2025.1535823 |
|  | Chromatin |  |
| ***IRF7*** | Microglial neuroinflammation | Roy ER et al, 2020. doi: 10.1172/JCI133737 |
|  | Synapse loss |  |
|  | Aβ |  |
|  | Cognition | Carús-Cadavieco M et al, 2024. doi: 10.1038/s41387-024-00325-y |
|  | T2D |  |
|  | M1 polarization | Tanaka T et al, 2015. doi: 10.1002/glia.22770 |
|  | Glycolysis | Meng T et al, 2025. doi: 10.1007/s00018-025-05608-w |
|  | Inflammation | Kuroda M et al, 2020. doi: 10.1371/journal.pone.0233390 |
|  | Obesity |  |
|  | Kinase cascades | Ning S et al, 2011. doi: 10.1038/gene.2011.21 |
|  | Phosphoprotein |  |
|  | Lipids | Hayakawa S et al, 2022. doi: 10.1172/jci.insight.138539 |
| ***ISOC2***  *DECREASED promoter methylation (potentially increased expression) in bezisterim vs placebo* | Mitochondrial integrity | Harmonize 3.0. Diamant I, 2024. [https://maayanlab.cloud/Harmonizome/gene/ISOC2#:~:text=Collectively%2C%20these%20studies%20indicate%20that,cycle%20control%20and%20mitochondrial%20integrity](https://protect.checkpoint.com/v2/r01/___https://maayanlab.cloud/Harmonizome/gene/ISOC2___.YzJ1OmJpb3ZpZTE6YzpvOjljNzMyZTM5MjY4ZDg1MDA1MDcyODAxYjc4YTlmZjRhOjc6Nzg0ZjpjODEzYTlhZWE3YWZhYTZlZTI5MjljMTQzZjMzNzFkZmE5MDE4OGNjZjRmMWVhYjNmZTk0NjIyMTAzOTU2NzkxOnA6RjpG" \l ":~:text=Collectively%2C%20these%20studies%20indicate%20that,cycle%20control%20and%20mitochondrial%20integrity) |
|  | INK4a | Sato A et al, 1991. doi: <http://dx.doi.org/10.3177/jnsv.37.419>  Huang X et al, 2007. doi: 10.1016/j.bbrc.2007.06.181 |
|  | Senescence | Huang X et al, 2007. doi: 10.1016/j.bbrc.2007.06.181 |
| ***ITGAL*** | Inflammation | Walling BL, Kim M, 2018. doi: 10.3389/fimmu.2018.00952 |
|  | AD (neutrophil) | Zenaro E et al, 2015. doi: 10.1038/nm.3913 |
|  | Cognition |  |
|  | Obesity | Wu H, Ballantyne CM, 2020. doi: 10.1161/CIRCRESAHA.119.315896 |
|  | Kinase cascades | Sánchez-Martín L et al, 2004. doi: 10.1074/jbc.M400905200 |
|  | Phosphoprotein | Jahan F et al, 2018. doi: 10.1074/jbc.RA118.004318 |
| ***ITGB8*** | Microglial neuroinflammation | Yin Z et al, 2023. doi: 10.1038/s41590-023-01627-6  Hagemeyer N et al, 2014. doi: 10.15252/embj.201490345 |
|  | AD (TGFβ) |  |
|  | T2D | Kostidou E et al, 2009. doi: 10.1016/j.clinbiochem.2008.12.007 |
| ***ITPKB*** | Kinase cascade | Stygelbout V et al, 2014. doi: 10.1093/brain/awt344 |
|  | ERK |  |
|  | pTau |  |
|  | Aβ |  |
|  | Significantly ↑ AD Plasma Proteomics | Ali M, et al, 2025. doi: 10.1038/s41591-025-03833-1 Erratum: 2025. doi: 10.1038/s41591-025-03970-7 |
|  | NOMINATED** | Agora. [https://agora.adknowledgeportal.org/genes/ENSG00000164136](https://protect.checkpoint.com/v2/r01/___https://agora.adknowledgeportal.org/genes/ENSG00000164136___.YzJ1OmJpb3ZpZTE6YzpvOjljNzMyZTM5MjY4ZDg1MDA1MDcyODAxYjc4YTlmZjRhOjc6MjJhYjoxOTA4MWY0MTE3MWJjYTBlMmE1NjM4ZWUxNmZlZDA4MmIzYjA1N2NlY2FiZDZjMzc3NTFiZWRmNzYxZDgwZWQ0OnA6RjpG) |
| ***ITPKC*** | Kinase cascade | Stygelbout V et al, 2014. doi: 10.1093/brain/awt344 |
|  | ERK |  |
|  | Lipids |  |
|  | Astroglial neuroinflammation |  |
|  | Cognition | Hu Y et al, 2024. doi: 10.1007/s12031-024-02221-7 |
| ***ITPR2*** | Inflammation | Zhang Y et al, 2023. doi: 10.1016/j.ecoenv.2023.115073 |
|  | Pyroptosis |  |
|  | Senescence | Ziegler DV et al, 2021. doi: 10.1038/s41467-021-20993-z |
|  | Aging |  |
|  | NOMINATED** | Agora. [https://agora.adknowledgeportal.org/genes/ENSG00000123104](https://protect.checkpoint.com/v2/r01/___https://agora.adknowledgeportal.org/genes/ENSG00000123104___.YzJ1OmJpb3ZpZTE6YzpvOjljNzMyZTM5MjY4ZDg1MDA1MDcyODAxYjc4YTlmZjRhOjc6MTY3Yzo1YTBjOWUzZGY1ZGY2MDkzYmQ5ZGYxNWE1NWEyMTk0ZmI0YzYzYTI2ZmYzNTRmNTVkYWFmMTM0YTAyNTZhMjhjOnA6RjpG) |
| ***JAK2*** | Kinase cascades | Kong X et al, 2019. doi: 10.1016/j.bbi.2019.01.027 |
|  | Cognition |  |
|  | Microglial neuroinflammation |  |
|  | IL6 |  |
|  | M1 polarization | Huang Z et al, 2024. doi: 10.1016/j.heliyon.2024.e34715  Orecchioni M et al, 2019. doi: 10.3389/fimmu.2019.01084. Erratum: doi: 10.3389/fimmu.2020.00234 |
|  | T2D | Zhang H et al, 2017. doi: 10.1016/j.kint.2017.03.027 |
|  | Obesity | Desai HR et al, 2017. doi: 10.1038/s41598-017-07923-0 |
|  | Inflammation |  |
| ***JDP2*** | Astrocyte neuroinflammation | Ku CC et al, 2020. doi: 10.1038/s41598-020-61692-x |
|  | ROS |  |
|  | Cognition | Wang XH et al, 2018. doi: 10.1016/j.bbrc.2018.08.055 |
|  | Apoptosis |  |
|  | Significantly ↑ AD Plasma Proteomics | Ali M, et al, 2025. doi: 10.1038/s41591-025-03833-1 Erratum: 2025. doi: 10.1038/s41591-025-03970-7 |
| ***JMJD6*** | Cognition | Merchant JP et al, 2023. doi: 10.1038/s42003-023-04791-5 |
|  | Aβ |  |
|  | pTau |  |
|  | Inflammation | Tikhanovich I et al, 2015. doi: 10.1074/jbc.M115.653543 |
|  | TRAF6 |  |
|  | NFκB |  |
|  | Obesity | Hu YJ et al, 2015. doi: 10.1093/nar/gkv645  Zhou J et al, 2022. doi: 10.1016/j.molcel.2022.06.003 |
|  | Lipids |  |
|  | Histone modification | Unoki M et al, 2013. doi: 10.1074/jbc.M112.433284 |
|  | Significantly ↑ AD Plasma Proteomics | Ali M, et al, 2025. doi: 10.1038/s41591-025-03833-1 Erratum: 2025. doi: 10.1038/s41591-025-03970-7 |
|  | NOMINATED** | Agora. [https://agora.adknowledgeportal.org/genes/ENSG00000070495](https://protect.checkpoint.com/v2/r01/___https://agora.adknowledgeportal.org/genes/ENSG00000070495___.YzJ1OmJpb3ZpZTE6YzpvOjljNzMyZTM5MjY4ZDg1MDA1MDcyODAxYjc4YTlmZjRhOjc6Y2Y1NToyOTljZWMwODExNDMzYTBlOTY5YTI0ZjQ1ZDNlMjhlY2U2M2M3YWQwMTU3N2Y1YmJkYWRhMWMwMGE3ODc4YTZlOnA6RjpG) |
| ***KARS1*** | Inflammation | Yun K et al, 2025. doi: 10.1038/s41598-025-96046-y |
|  | Significantly ↑ AD & PD Plasma Proteomics | Ali M, et al, 2025. doi: 10.1038/s41591-025-03833-1 Erratum: 2025. doi: 10.1038/s41591-025-03970-7 |
| ***KCNN4*** | Microglial neuroinflammation | Maezawa I et al, 2012. doi: 10.1155/2012/868972  Kaushal V et al, 2007. doi: 10.1523/JNEUROSCI.3593-06.2007 |
|  | Cognition | Bal NV et al, 2025. doi: 10.1038/s41598-025-89097-8  Mauler F et al, 2004. doi: 10.1111/j.1460-9568.2004.03615.x |
|  | DM | Soret B et al, 2022. doi: 10.1515/hsz-2022-0232 |
|  | Glycolysis | Fan J et al, 2022. doi: 10.3390/ijms23136958 |
|  | NOMINATED** | Agora. [https://agora.adknowledgeportal.org/genes/ENSG00000104783](https://protect.checkpoint.com/v2/r01/___https://agora.adknowledgeportal.org/genes/ENSG00000104783___.YzJ1OmJpb3ZpZTE6YzpvOjljNzMyZTM5MjY4ZDg1MDA1MDcyODAxYjc4YTlmZjRhOjc6MjhmOTo5MjVkNTg1OTEyM2JhNTU5ZDg2YjA4YzkxYjgyZmNiZGMyN2YzYjM4YTM2ZDJkNDVlM2FkNjI5MjVjNTY4ZDIyOnA6RjpG) |
| ***KCTD1*** | Cognition | Liu Z et al, 2016. doi: 10.1038/srep32658 |
|  | AD |  |
|  | Fibril formation |  |
|  | Apoptosis |  |
|  | Obesity | Pirone L et al, 2019. doi: 10.1016/j.bbalip.2019.08.010 |
|  | Lipids |  |
| ***KCTD12*** | Cognition | Cheng J et al, 2024. doi: 10.1073/pnas.2315707121 |
|  | Neuropsychiatric |  |
|  | Stress response | Deng SL et al, 2021. doi: 10.1016/j.phrs.2020.105355 |
|  | Significantly ↑ AD microglial transcripts with Dementia | Kosoy KDTR. et al, 2025. doi: 10.1038/s41593-025-02020-2 |
|  | Braak* |  |
| ***KDM2B*** | Kinase cascades | Li X et al, 2023. doi: 10.1002/iid3.985 |
|  | NFκB |  |
|  | AP-1 pathways |  |
|  | IL6 | Zhou Q et al, 2020. doi: 10.1038/s41423-019-0251-z |
|  | Chromatin |  |
|  | M1 polarization |  |
|  | T2D | Zacharopoulou N et al, 2020. doi: 10.1080/15384047.2020.1736481 |
|  | Kinase cascade |  |
|  | Glycolysis | Xie Z et al, 2021. doi: 10.1080/21655979.2021.2005931 |
|  | TF | Zhou Q et al, 2020. doi: 10.1038/s41423-019-0251-z |
|  | Chromatin |  |
| ***KDM4C*** | Histone | Fu J, An L, 2024. doi: 10.14336/AD.2024.0899 |
|  | Chromatin |  |
|  | Senescence |  |
|  | Inflammation | Ma Y et al, 2025. doi: 10.1186/s12964-024-02006-w |
|  | NFκB |  |
|  | TNF |  |
|  | Glycolysis | Lin CY et al, 2022. doi: 10.1002/ctm2.764 |
| ***KDM5C*** | Chromatin | Qi S et al, 2021. doi: 10.1186/s12974-021-02120-3 |
|  | Microglial inflammation |  |
|  | IRF 4/5 |  |
|  | Cytokines |  |
|  | Cognition | Han J et al, 2025. doi: 10.1016/j.neuint.2025.105975 |
|  | Obesity | Vergnes L et al, 2024. doi: 10.1210/jendso/bvae029 |
| ***KLF10*** | AD | Cheng Z et al, 2018. doi: 10.3389/fncel.2018.00325  Subramaniam M et al, 2010. doi: 10.1002/biof.67 |
|  | Aβ |  |
|  | Inflammation |  |
|  | DM |  |
|  | Phosphoprotein | Luo HY et al, 2022. doi: 10.1016/j.gendis.2022.06.005 |
|  | Chromatin | Mishra VK et al, 2017. doi: 10.1158/0008-5472  Yang S et al, 2022. doi: 10.15252/embr.202154229 |
|  | TF |  |
|  | Lipids |  |
| ***KLF7*** | Kinase cascades | Zhang Z et al, 2024. doi: 10.7150/ijbs.86385  Huang WH et al, 2020. doi: 10.26355/eurrev_202006_21693 |
|  | M1 polarization |  |
|  | T2D | Kawamura Y et al, 2006. doi: 10.1210/me.2005-0138 |
|  | Glycolysis | Wang C et al, 2023. doi: 10.1038/s41467-023-36712-9 |
|  | Phosphoprotein | Jha K et al, 2024. doi: 10.1016/j.bbagrm.2023.195003 |
| ***LAPTM5*** | Macrophages | Glowacka WK et al, 2012. doi: 10.1074/jbc.M112.355917 |
|  | Microglial neuroinflammation | Sun PP et al, 2024. doi: 10.1016/j.heliyon.2024.e36705 |
|  | M1 polarization | Hua W et al, 2023. doi: 10.1007/s12035-023-03484-8 |
|  | Kinase cascades | Zhang Z et al, 2022. doi: 10.3389/fnmol.2022.971361 |
|  | NOMINATED** | Agora. [https://agora.adknowledgeportal.org/genes/ENSG00000162511](https://protect.checkpoint.com/v2/r01/___https://agora.adknowledgeportal.org/genes/ENSG00000162511___.YzJ1OmJpb3ZpZTE6YzpvOjljNzMyZTM5MjY4ZDg1MDA1MDcyODAxYjc4YTlmZjRhOjc6MGU3MjpjNzQzMjRmZDY1OGVmNTAxZmMwMTg3NzQwNGRkOTFjZTA0ZDVmNDM3OTE4MjdhNjQwZWE5MWMwOTY2YTNkNWY3OnA6RjpG) |
| ***LEPR*** | Inflammation | Wang Y et al, 2023. doi: 10.1186/s10020-023-00702-w |
|  | M1 polarization |  |
|  | Glycolysis | Xu Y et al, 2020. doi: 10.3892/mmr.2019.10855 |
|  | Kinase cascades | Park HK, Ahima RS, 2014. doi: 10.12703/P6-73 |
|  | Phosphoprotein | Gong Y et al, 2007. doi: 10.1074/jbc.M702838200 |
| ***LHPP*** | Cognition | Hahs DW et al, 2006. doi: 10.1002/ajmg.b.30257  Zhuang L et al, 2024. doi: 10.1016/j.biopsych.2023.08.026 |
| ***LIME1*** | Inflammation | Park I et al, 2020. doi: 10.14348/molcells.2020.0124 |
|  | Kinase cascades | Hur EM et al, 2003. doi: 10.1084/jem.20030232 |
|  | Phosphoprotein |  |
| ***LINC01126*** | AD | Tang S et al, 2025. doi: 10.1111/odi.15033  Abbayya K et al, 2015. doi: 10.4103/1947-2714.15932 |
|  | Inflammation |  |
|  | Periodontitis |  |
|  | Kinase cascades | Zhou M et al, 2021. doi: 10.1111/cpr.12957 |
| ***LINC01806*** | AD | Rahman R et al, 2019. doi.org/10.1101/482828 |
|  | Cognition |  |
|  | PD transcriptomics | Pantaleo E et al, 2022. doi: 10.3390/genes13050727 |
|  | T2D | Rattanapan Y et al, 2025. doi: 10.3390/biology14040424 |
| ***LIPG*** | Cognition | Yun SM et al, 2019. doi: 10.1186/s12888-019-2174-8 |
|  | Macrophage | Yu JE et al, 2018. doi: 10.14670/HH-11-905 |
|  | Neuro-inflammation |  |
|  | Obesity |  |
|  | Lipids |  |
|  | M1 polarization | Huang J et al, 2010. doi: 10.1016/j.trsl.2010.05.003 |
|  | Inflammation | Hong C et al, 2021. doi: 10.1038/s41417-020-0188-5 |
| ***LPCAT4*** | M1 polarization | Law SH et al, 2019. doi: 10.3390/ijms20051149 |
|  | Endothelial damage |  |
|  | Significantly ↑ AD microglial transcripts with Dementia | Kosoy KDTR. et al, 2025. doi: 10.1038/s41593-025-02020-2 |
| ***LZTFL1*** | Inflammation | Song G et al, 2024. doi: 10.1016/j.molimm.2024.01.010 |
|  | Kinase cascades |  |
|  | ERK |  |
|  | COVID-19 effector | Bose P, 2021. [https://www.news-medical.net/news/20211108/Identification-of-a-candidate-effector-gene-at-COVID-19-risk-locus.aspx#:~:text=LZTFL1%20and%20COVID%2D19,risk%20of%20respiratory%20failure%20twofold](https://protect.checkpoint.com/v2/r01/___https://www.news-medical.net/news/20211108/Identification-of-a-candidate-effector-gene-at-COVID-19-risk-locus.aspx___.YzJ1OmJpb3ZpZTE6YzpvOjljNzMyZTM5MjY4ZDg1MDA1MDcyODAxYjc4YTlmZjRhOjc6OWYzZDo4NzdmZmU3YTMxMWFjNDJhMDc4MTAzNmE3MGU1ODhhZmJhYzdjYzlhMzk1MzRiOTQ5NTljZmRiMTc3MTJlNTEyOnA6RjpG#:~:text=LZTFL1%20and%20COVID%2D19,risk%20of%20respiratory%20failure%20twofold) |
| ***MAF1*** | Cognition | Han Y et al, 2024. doi: 10.1093/brain/awae015 |
|  | Spinogenesis |  |
|  | Obesity | Bonhoure N et al, 2015. doi: 10.1101/gad.258350.115 |
|  | Significantly ↑ AD microglial transcripts with Dementia | Kosoy KDTR. et al, 2025. doi: 10.1038/s41593-025-02020-2 |
|  | Braak* |  |
| ***MAGT1*** | Kinase cascades | Wu Y et al, 2022. doi: 10.1080/21655979.2022.2037214 |
|  | MAPK |  |
|  | Phosphoprotein | Hu Y et al, 2013. doi: 10.1016/j.bbrc.2013.05.048 |
| ***MAML1*** | Cognition | Kapoor A, Nation DA, 2021. doi: 10.1016/j.semcdb.2020.12.011 |
|  | Notch signalling |  |
|  | Inflammation | Jin B et al, 2010. doi: 10.1074/jbc.M109.078865 |
|  | Kinase cascade |  |
|  | NFκB |  |
|  | TF | Hansson ML et al, 2009. doi: 10.1093/nar/gkp163 |
|  | Chromatin |  |
| ***MAMSTR***  *Decreased promoter methylation (potentially increased expression) in bezisterim subjects compared to placebo* | DMRs in Braak* | Zhang L et al, 2020. doi: 10.1038/s41467-020-19791-w |
|  | Microglial neuroinflammation | Deczkowska A et al, 2017. doi: 10.1038/s41467-017-00769-0 |
|  | MEF2C |  |
|  | IFN-I |  |
| ***MAN2A2*** | T2D epigenetics | Lai L et al, 2025. doi: 10.1186/s12933-024-02558-8. Erratum: doi: 10.1186/s12933-025-02760-2 |
| ***MAP2K1*** | Cognition | Zou T et al, 2023. doi: 10.1371/journal.pone.0295320 |
|  | Inflammation | Chen MJ et al, 2021. doi: 10.1002/jnr.24829 |
|  | Astrocyte reactivity | Kaminska P et al, 2025. doi: <http://dx.doi.org/10.1002/ar.21047> |
|  | Glycolysis | Papa S et al, 2019. doi: 10.1038/s41388-018-0582-8 |
|  | M1 polarization | Long ME et al, 2017. doi: 10.4049/jimmunol.1601059 |
|  | Lipids | Yousefi B et al, 2012. doi: 10.5681/bi.2012.019 |
| ***MAP2K3*** | Dementia | Munoz L et al, 2010. doi: 10.1016/j.neuropharm.2009.11.010 |
|  | Cognition | Huentelman MJ et al, 2018. doi: 10.3389/fnagi.2018.00155  Spencer BE et al, 2022. doi: 10.1002/trc2.12321 |
|  | Aging |  |
|  | Microglial neuroinflammation | Iba M et al, 2023. doi: 10.1126/scitranslmed.abq6089 |
|  | C1q | Dejanovic B et al, 2018. doi: <http://dx.doi.org/10.1016/j.neuron.2018.10.014> |
|  | Hypothalamic inflammation | Bian L et al, 2013. doi: 10.1093/hmg/ddt291 |
|  | T2D | Lim AK et al, 2009. doi: 10.1007/s00125-008-1215-5  Bian L et al, 2013. doi: 10.1093/hmg/ddt291 |
|  | Obesity |  |
| ***MAP2K6*** | AD | Zhu X et al, 2001. doi: 10.1046/j.1471-4159.2001.00597.x |
|  | Microglial neuroinflammation | Culbert AA et al, 2006. doi: 10.1074/jbc.M513646200 |
|  | Cognition | Munoz L, Ammit AJ, 2010. doi: 10.1016/j.neuropharm.2009.11.010 |
|  | Obesity | Matesanz N et al, 2017. doi: 10.1038/s41467-017-00948-z |
|  | T2D |  |
|  | Inflammation | Lee S et al, 2021. doi: 10.3390/ijms222413559 |
|  | Lipids |  |
|  | M1 polarization | Cheng Y et al, 2018. doi: 10.1002/mc.22822 |
| ***MAP3K12*** | T2D | Köster KA et al, 2024. doi: 10.3390/cells13040333 |
|  | Obesity |  |
|  | Microglial neuroinflammation | Wan W et al, 2021. doi: 10.1080/21655979.2021.2008638 |
|  | M1 polarization | Neamatallah T, 2019. doi: 10.4103/JMAU.JMAU_68_18 |
|  | Cognition | Le Pichon CE et al, 2017. doi: 10.1126/scitranslmed.aag0394  Köster KA et al, 2024. doi: 10.3390/cells13040333 |
|  | Motor neuron death | Bos PH et al, 2019. doi: 10.1016/j.chembiol.2019.10.005 |
| ***MAP4K1*** | Neuroprotective | Bos PH et al, 2019. doi: 10.1016/j.chembiol.2019.10.005  Lasham DJ et al, 2023. doi: 10.1186/s13041-023-01066-2 |
|  | Cognition |  |
|  | Microglial neuroinflammation | Zhang B et al, 2019. doi: 10.3389/fncel.2018.00531 |
|  | M1 polarization |  |
| ***MAP4K5*** | Neuroprotective | Bos PH et al, 2019. doi: 10.1016/j.chembiol.2019.10.005 |
|  | Microglial neuroinflammation | Wang X et al, 2025. doi: 10.1111/cns.70395. Erratum: doi: 10.1111/cns.70640  Pérez-Cabello JA, et al, 2023. [https://doi.org/10.1101/2023.01.23.524851](https://protect.checkpoint.com/v2/r01/___https://doi.org/10.1101/2023.01.23.524851___.YzJ1OmJpb3ZpZTE6YzpvOjljNzMyZTM5MjY4ZDg1MDA1MDcyODAxYjc4YTlmZjRhOjc6OGM5Yjo0ZWUwYzMxNGVjZTNiNTMxMGVlYWI5OGQ3Y2U1OGEzZGMyNzBiMTMxNzUyNWQ5NTlhZDE1MjcyNmM1YzlkOTM2OnA6RjpG) |
| ***MARK2*** | Neuro-inflammation | Gu GJ et al, 2013. doi: 10.3233/JAD-2012-121357. |
|  | pTau |  |
|  | Microglial neuroinflammation | Asthana S et al, 2024. doi: 10.1111/cbdd.14592 |
|  | T2D | Ruiz M et al, 2016. doi: 10.2337/db15-0238  Klutho PJ et al, 2011. doi: 10.1371/journal.pone.0029304 |
|  | Lipids |  |
|  | Obesity |  |
|  | M1 polarization | DiBona VL et al, 2019. doi: 10.1186/s12974-018-1390-3 |
| ***MARK3*** | pTau | Lund H et al, 2014. doi: 10.1186/2051-5960-2-22 |
|  | Microglial neuroinflammation | Correani V et al, 2017. doi: 10.1002/pmic.201600439 |
|  | Obesity | Lennerz JK et al, 2010. doi: 10.1128/MCB.01472-09 |
|  | Kinase cascades | Drewes G et al,1995. doi: 10.1074/jbc.270.13.7679 |
|  | Phosphoprotein | Sultanakhmetov G et al, 2024. doi: 10.1111/gtc.13101 |
| ***MAT2A*** | AD | Ryder MI, 2020. doi: 10.1002/JPER.20-0104  Jiang L et al, 2023. doi: 10.1080/20002297.2023.2292375 |
|  | Gingivitis |  |
|  | Kinase cascade |  |
|  | NFκB |  |
|  | Phosphoprotein | Ramani K et al, 2015. doi: 10.1002/jcp.24839 |
|  | T2D | Rajabian N et al, 2023. doi: 10.1038/s41467-023-36483-3 |
|  | Muscle strength |  |
|  | Obesity | Zhao C et al, 2018. doi: 10.1016/j.bbalip.2017.11.001  Sáenz de Urturi D et al, 2022. doi: 10.1038/s41467-022-28749-z |
|  | Chromatin |  |
| ***MATR3*** | ALS | Malik AM et al, 2021. doi: 10.1172/jci.insight.143948  Malik AM et al, 2018. doi: 10.7554/eLife.35977 |
|  | FTD |  |
|  | Aβ |  |
|  | Neuro-degeneration |  |
| ***MBNL1*** | AD | Deng S et al, 2023. doi: 10.1097/CM9.0000000000002214  Zhang F et al, 2017. doi: 10.1093/hmg/ddx190 |
|  | Glucose metabolism |  |
|  | Splicing defects |  |
|  | Inflammation | Gabel AM et al, 2025. doi: 10.1371/journal.pone.0321148  Sun C et al, 2015. doi: 10.1152/japplphysiol.00744.2014 |
|  | T2D | Gong Y et al, 2024. doi: 10.1186/s10020-024-00991-9  Echeverria GV et al, 2014. doi: 10.1093/nar/gkt1020 |
|  | Kinase cascades | Botta A et al, 2013. doi: 10.1038/cddis.2013.291 |
|  | Phosphoprotein |  |
| ***MCM2*** | Inflammation | Li LY et al, 2021. doi: 10.1016/j.jaci.2020.11.026  Davies RJ et al, 2004. doi: 10.1111/j.1463-1318.2004.00567.x |
|  | AD | Bonda DJ et al, 2009. doi: 10.18632/aging.10004 |
|  | Phosphoprotein | Fei L, Xu H, 2018. doi: 10.1186/s13578-018-0242-2 |
|  | Kinase cascades | Montagnoli A et al, 2006. doi: 10.1074/jbc.M512921200 |
| ***MECP2*** | Cognition | Kim B et al, 2019. doi: 10.5213/inj.1938196.098 |
|  | Microglial neuroinflammation | Wittrahm R et al, 2021. doi: 10.3390/cells10040860 |
|  | Obesity | Liu C et al, 2020. doi: 10.2337/db19-0502 |
|  | Chromatin | Schmidt A et al, 2020. doi: 10.3390/cells9040878 |
|  | Significantly ↑ AD Plasma Proteomics | Ali M, et al, 2025. doi: 10.1038/s41591-025-03833-1 Erratum: 2025. doi: 10.1038/s41591-025-03970-7 |
| ***MGAT1*** | Glycolysis | Tang X et al, 2023. doi: 10.1038/s41598-023-34787-4  Li Y et al, 2020. doi: 10.1002/1873-3468.13596 |
|  | Glut1 |  |
|  | Inflammation | Radovani B, Gudelj I, 2022. doi: 10.3389/fimmu.2022.893365 |
|  | M1/M2 polarization | Mantuano NR et al, 2019. doi: 10.1016/j.phrs.2019.104285 |
|  | DM | Rudman N et al, 2019. doi: 10.1002/1873-3468.13495 |
|  | Synapses | Parkinson W et al, 2013. doi: 10.1242/dev.099192 |
|  | Significantly ↑ AD microglial transcripts with Dementia | Kosoy KDTR. et al, 2025. doi: 10.1038/s41593-025-02020-2 |
|  | Braak* |  |
| ***MIA3*** | Vascular inflammation | Lei Y et al, 2021. doi: 10.3389/fendo.2021.748216 |
|  | Significantly ↑ AD microglial transcripts with Dementia | Kosoy KDTR. et al, 2025. doi: 10.1038/s41593-025-02020-2 |
| ***MICB*** | Neuro-inflammation | Pogoda-Wesołowska A et al, 2025. doi: 10.1038/s41598-025-12589-0 |
|  | Cognition | Goddard CA et al, 2007. doi: 10.1073/pnas.0702023104 |
| ***MIR4458HG*** | M1 polarization | Zhao S et al, 2025. doi: 10.3389/fimmu.2025.1523190 |
| ***MKRN1*** | T2D | Lee MS et al, 2018. doi: 10.1038/s41467-018-05721-4 |
|  | Obesity |  |
|  | Kinase cascades |  |
| ***MLF1*** | Senescence | Lv J et al, 2025. doi: 10.1093/nar/gkae1176 |
|  | Chromatin |  |
|  | Neuro-degeneration | Sun Y, et al, 2017. doi: 10.1016/j.bbamcr.2017.01.016 |
|  | Significantly ↑ AD Plasma Proteomics | Ali M, et al, 2025. doi: 10.1038/s41591-025-03833-1 Erratum: 2025. doi: 10.1038/s41591-025-03970-7 |
| ***MPC1*** | pTau | Ceyzériat K et al, 2024. doi: 10.1016/j.nbd.2024.106623 |
|  | Aβ |  |
|  | Cognition |  |
|  | Inflammation | Zhu B et al, 2023. doi: 10.1126/sciimmunol.adf0348 |
|  | COVID-19 |  |
|  | Significantly ↑ AD Plasma Proteomics | Ali M, et al, 2025. doi: 10.1038/s41591-025-03833-1 Erratum: 2025. doi: 10.1038/s41591-025-03970-7 |
| ***MPI*** | Aβ | Liang C et al, 2025. doi: 10.1002/advs.202409105 |
|  | Glycolysis | Liang R et al, 2024. doi: 10.1038/s41467-024-46415-4 |
| ***MTHFR*** | AD | Román GC et al, 2019. doi: 10.3390/ijms20020319 |
|  | ↑ Homocysteine |  |
|  | Microglial neuroinflammation | GEN, 2023. [https://www.genengnews.com/topics/translational-medicine/alzheimers-risk-increased-by-microglia-mutation/](https://protect.checkpoint.com/v2/r01/___https://www.genengnews.com/topics/translational-medicine/alzheimers-risk-increased-by-microglia-mutation/___.YzJ1OmJpb3ZpZTE6YzpvOjljNzMyZTM5MjY4ZDg1MDA1MDcyODAxYjc4YTlmZjRhOjc6YzQ2NzoyMjlhNTRkYjVhNzVkNTA0ZThlMzkwZjNiOGI3NjlhYzI1OWRiZjZmODMwZDNmMmU0ZmZlZDFlNjU0Yzc4YjExOnA6RjpG) |
|  | Psychiatric disorders | Zhang YX et al, 2022. doi: 10.3389/fpsyt.2022.976428 |
|  | Obesity | Raghubeer S et al, 2021. doi: 10.3390/nu13124562 |
|  | Lipids |  |
|  | T2D | Diniz TG et al, 2021. doi: 10.3389/fphys.2020.618672 |
| ***MTOR*** | pTau | Davoody S et al, 2024. doi: 10.1111/cns.14463 |
|  | Aβ |  |
|  | Cognition | Perluigi M et al, 2021. doi: 10.1016/j.freeradbiomed.2021.04.025 |
|  | ROS |  |
|  | Microglial neuroinflammation | Xie L et al, 2014. doi: 10.4049/jimmunol.1303492 |
|  | M1 polarization |  |
|  | Glycolysis | Linke M et al, 2017. doi: 10.1002/1873-3468.12711 |
|  | Obesity | Magdalon J et al, 2017. doi: 10.1590/S1679-45082017RB4106 |
|  | T2D | Tuo Y, Xiang M, 2019. doi: 10.1002/JLB.3MR0317-095RR |
| ***MTTP*** | Lipids | Jamil H et al,1995. doi: 10.1074/jbc.270.12.6549 |
|  | M1 polarization | Hewing B et al, 2013. doi: 10.1016/j.atherosclerosis.2012.12.026 |
|  | Obesity | Bartels ED et al, 2002. doi: 10.2337/diabetes.51.4.1233 |
|  | Cognition | Dimache AM et al, 2021. doi: 10.3390/nu13062118 |
| ***MUTYH*** | Inflammation | Oka S, Nakabeppu Y, 2011. doi: 10.1111/j.1349-7006.2011.01869.x. |
|  | ROS |  |
|  | Phosphoprotein | Kundu S et al, 2010. doi: 10.1016/j.dnarep.2010.07.002 |
| ***MYL12A*** | Inflammation | Yokoyama M et al, 2021. doi: 10.3389/fimmu.2020.594297 |
|  | M1 polarization |  |
| ***MYO1G*** | Microglial neuroinflammation | Wang Y et al, 2021. doi: 10.1038/s41419-021-03983-3 |
| ***N4BP2L1*** | Obesity | Watanabe K et al, 2019. doi: 10.1016/j.bbrep.2019.100676 |
| ***NAGK*** | NOD2 | Stafford CA et al,2022. doi: 10.1038/s41586-022-05125-x |
|  | Significantly ↑ AD Plasma Proteomics | Ali M, et al, 2025. doi: 10.1038/s41591-025-03833-1 Erratum: 2025. doi: 10.1038/s41591-025-03970-7 |
| ***NARF*** | Aging | Wang Y et al, 2022. doi: 10.1073/pnas.2118695119 |
|  | Obesity | Wegner L et al, 2007. doi: 10.2337/db06-0927 |
|  | T2D |  |
|  | Significantly ↑ AD Plasma Proteomics | Ali M, et al, 2025. doi: 10.1038/s41591-025-03833-1 Erratum: 2025. doi: 10.1038/s41591-025-03970-7 |
| ***NEAT1*** | AD | Zhao Y, et al, 2019. doi: https://doi.org/10.1101/643718 |
|  | pTau |  |
|  | Glycolysis |  |
|  | Microglial neuroinflammation | Pan Y et al, 2022. doi: 10.2147/JIR.S338162 |
|  | NFκB |  |
|  | M1 polarization | Wang H, et al, 2023. doi: 10.3389/fncel.2023.1182621 |
|  | Lipids | Pan Y et al, 2024. doi: 10.1007/s00018-023-05045-7  Huang-Fu N et al, 2018. doi: 10.3892/mmr.2017.8211  Chen X et al, 2019. doi: 10.1016/j.lfs.2019.116829 |
|  | DM | Wu X et al, 2022. doi: 10.1097/FJC.0000000000001177  Chen K et al, 2024. doi: 10.1080/15592294.2023.2293409  Yang YL et al, 2020. doi: 10.1038/s12276-020-0381-5 |
|  | Obesity | Corral A et al, 2022. doi: 10.1016/j.bcp.2022.115305  Lo PK et al, 2016. doi: 10.1007/s11515-016-1433-z |
|  | Cognition | Wu Y et al, 2022. doi: 10.1016/j.ymthe.2022.03.011 |
| ***NEU1*** | Microglial neuroinflammation | Allendorf DH, Brown GC, 2022. doi: 10.3389/fncel.2022.917884 |
|  | M1 polarization | Escalona E et al, 2024. doi: 10.3389/fimmu.2024.1462853 |
| ***NFE2L2*** | Atherosclerosis | Freigang S et al, 2011. doi: 10.1002/eji.201041316 |
|  | Obesity | He F et al, 2020. doi: 10.3390/ijms21134777 |
|  | Lipids |  |
|  | TF |  |
|  | Chromatin |  |
|  | Glycolysis | Song MY et al, 2021. doi: 10.3390/ijms22094376 |
|  | ↓ Neurogenesis | Anandhan A et al, 2021. doi: <http://dx.doi.org/10.1111/acel.13385>  Garcia AD et al, 2004. doi: http://dx.doi.org/10.1038/nn1340 |
|  | Phosphoprotein | Han LQ et al, 2018. doi: 10.3168/jds.2017-14257 |
| ***NFKB2*** | Inflammation | Tak PP, Firestein GS, 2001. doi: 10.1172/JCI11830 |
|  | AD | Jha NK et al, 2019. doi: 10.1111/jnc.14687  Jantaratnotai N et al, 2013. doi: 10.3233/JAD-122191 |
|  | Cognition | Yang D et al, 2024. doi: 10.1016/j.ejphar.2024.177038 |
|  | NFκB | Sim N et al, 2023. doi: 10.1038/s42003-023-04821-2 |
|  | Kinase cascades | Sun SC, 2011. doi: 10.1038/cr.2010.177 |
|  | TF | Jantaratnotai N et al, 2013. doi: 10.3233/JAD-122191  Bhatt D, Ghosh S, 2014. doi: 10.3389/fimmu.2014.0007 |
|  | Chromatin |  |
| ***NFKBIA*** | AD | Sun E et al, 2022. doi: 10.3390/ijms23168972 |
|  | Inflammation | Nist MD et al, 2024. doi: 10.1177/10998004241257664 |
|  | ERK | Li C et al, 2018. doi: 10.1159/000491059 |
|  | Microglial neuroinflammation | Varsamos I et al, 2025. doi: 10.7759/cureus.79367  Frakes AE et al, 2014. doi: 10.1016/j.neuron.2014.01.013  Dalal NV et al, 2012. doi: 10.1016/j.neulet.2012.08.060 |
|  | Glycolysis | Guo D et al, 2022. doi: 10.1016/j.cmet.2022.08.002  Londhe P et al, 2018. doi: 10.3389/fonc.2018.00104 |
|  | DM | Suryavanshi SV et al, 2017. doi: 10.3389/fphar.2017.00798  Meyerovich K et al, 2018. doi: 10.1530/JME-16-0183. |
|  | Lipids | Ringseis R et al, 2015. chrome-extension://efaidnbmnnnibpcajpcglclefindmkaj/http://eir-isei.de/2015/eir-2015-058-article.pdf |
|  | Obesity |  |
|  | M1 polarization | Li YF et al, 2022. doi: 10.3389/fnagi.2022.901117  Wu YG et al, 2023. doi: 10.4103/1673-5374.355747  Zhou L et al, 2020. doi: 10.3389/fphar.2020.01126 |
|  | NOMINATED** | Agora. [https://agora.adknowledgeportal.org/genes/ENSG00000100906](https://protect.checkpoint.com/v2/r01/___https://agora.adknowledgeportal.org/genes/ENSG00000100906___.YzJ1OmJpb3ZpZTE6YzpvOjljNzMyZTM5MjY4ZDg1MDA1MDcyODAxYjc4YTlmZjRhOjc6YzcwZDplZWYwMDQ0ZWM3NmI0ZTIxNDYwMjFhN2EwZGU0OTc4MmM1Y2M3NzhiODJjYWQxZGMzZjFhNDc5ZWZmMzhhNTE2OnA6RjpG) |
| ***NFKBIZ*** | Microglial neuroinflammation | GeneCards. [https://www.genecards.org/cgi-bin/carddisp.pl?gene=NFKBIZ#:~:text=NCBI%20Gene%20Summary%20for%20NFKBIZ,PubMed:16513645%2C%2016622025](https://protect.checkpoint.com/v2/r01/___https://www.genecards.org/cgi-bin/carddisp.pl?gene=NFKBIZ___.YzJ1OmJpb3ZpZTE6YzpvOjljNzMyZTM5MjY4ZDg1MDA1MDcyODAxYjc4YTlmZjRhOjc6ZjNkNDpmZmJjM2VmYjczZmRmYzg1NWVhYjQzZjY0ZTIxMmNhMjNkYjI5OWJkMTM5OWFmZGNjNDllMjAzZmEwNjJlNWM1OnA6RjpG#:~:text=NCBI%20Gene%20Summary%20for%20NFKBIZ,PubMed:16513645%2C%2016622025)  NIH, 2025. [https://www.ncbi.nlm.nih.gov/gene/64332](https://protect.checkpoint.com/v2/r01/___https://www.ncbi.nlm.nih.gov/gene/64332___.YzJ1OmJpb3ZpZTE6YzpvOjljNzMyZTM5MjY4ZDg1MDA1MDcyODAxYjc4YTlmZjRhOjc6MWE2ZTpmMWViOGM5MjQ2ZTEwNzY2YjhmOTJlMTdmYzA4ZDIyNGQxMGY5NmYwNTU5MTgzNjQzOTJhZGJkMTRlNmQ2ZjU2OnA6RjpG) |
|  | Kinase cascades |  |
|  | IL-6 |  |
|  | Cognition | Wahl D et al, 2024. doi: 10.1186/s12974-024-03182-9  Vazquez-Coto D et al, 2025. doi: 10.1016/j.bbr.2024.115264  Jong Huat T et al, 2024. doi: 10.1038/s41598-024-65248-1 |
|  | Glycolysis | Londhe P et al, 2018. doi: 10.3389/fonc.2018.00104 |
|  | M1 polarization | Gautam P et al, 2022. doi: 10.1002/ctm2.1032 |
|  | DM | Li X et al, 2020. doi: 10.1016/j.ymthe.2020.07.016  Suryavanshi SV et al, 2017. doi: 10.3389/fphar.2017.00798  Baker RG et al, 2011. doi: 10.1016/j.cmet.2010.12.008 |
|  | Obesity | Catrysse L et al, 2017. doi: 10.1016/j.tcb.2017.01.006  Hill AA et al, 2015. doi: 10.1016/j.molmet.2015.07.005 |
|  | TF | Bhatt D et al, 2014. doi: 10.3389/fimmu.2014.00071 |
|  | Chromatin |  |
|  | NOMINATED** | Agora. [https://agora.adknowledgeportal.org/genes/ENSG00000144802](https://protect.checkpoint.com/v2/r01/___https://agora.adknowledgeportal.org/genes/ENSG00000144802___.YzJ1OmJpb3ZpZTE6YzpvOjljNzMyZTM5MjY4ZDg1MDA1MDcyODAxYjc4YTlmZjRhOjc6NjllOTpjNTE0N2E0ZTg1YzUyZjY1YzUxYjI5OTJmNDc3ZDIyYWM4M2Q0NjQ1NTgyYmVmOTBlODU4YjMxN2EyNTU5MTVkOnA6RjpG) |
| ***NFYC*** | Glycolysis | Benatti P et al, 2016. doi: 10.18632/oncotarget.6453 |
|  | T2D |  |
|  | TF | Ly LL et al, 2013. www.ajcr.us /ISSN:2156-6976/ajcr0000217 |
| ***NINJ2*** | Neuro-inflammation | Liu S et al, 2025. doi: 10.1038/s41598-025-02097-6  Sorosina M et al, 2022. doi: 10.3390/genes13111946 |
|  | SLAMF8 |  |
|  | Monocytes |  |
|  | Neurovascular risk | Lin KP et al, 2011. doi: 10.1371/journal.pone.0020573 |
| ***NLRP1*** | Inflammation | Fenini G et al, 2020. doi: 10.3390/ijms21134788 |
|  | Microglial neuroinflammation (Brain injury) | Mi L et al, 2022. doi: 10.3389/fimmu.2022.863774 |
|  | AD | Španić E et al, 2022. doi: 10.3390/cells11142223 |
|  | Kinase cascades | Jenster LM et al, 2023. doi: 10.1084/jem.20220837 |
|  | Phosphoprotein |  |
|  | Lipids | Li X et al, 2024. doi: 10.1016/j.bbalip.2023.159428 |
| ***NME3***  *Decreased DNA promoter methylation in bezisterim vs placebo subjects* | Cell survival (mitochondria) | Chen CW et al, 2019. doi: 10.1073/pnas.1818629116 |
|  | Inflammation | Chen CW et al, 2020. doi: 10.3390/ijms21145048 |
|  | ROS |  |
|  | Kinase cascades | Hoff S et al, 2018. doi: 10.1074/jbc.RA117.000847 |
| ***NOTCH4*** | Inflammation | Harb H et al, 2021. doi: 10.1016/j.immuni.2021.04.002 |
|  | Glycolysis | Slaninova V et al, 2016. doi: 10.1098/rsob.150155 |
|  | Obesity | Lai PY et al, 2013. doi: 10.1016/j.bbrc.2012.12.024 |
| ***NR3C1*** | Stress response | Berretta E et al, 2021. doi: 10.1016/j.neubiorev.2021.03.003  González Ramírez C et al, 2020. doi: 10.1016/j.psychres.2020.112797 |
|  | DM | Wu T et al, 2023. doi: 10.1080/15548627.2023.2200625  de Souza MLM et al, 2022. doi: 10.1016/j.lfs.2022.120940  Mohan A et al, 2021. [http://www.molvis.org/molvis/v27/429](https://protect.checkpoint.com/v2/r01/___http://www.molvis.org/molvis/v27/429___.YzJ1OmJpb3ZpZTE6YzpvOjljNzMyZTM5MjY4ZDg1MDA1MDcyODAxYjc4YTlmZjRhOjc6YjlmMTo0MDMwOGQ4YmYyYjc1OWQ5MzVmMDQ5ZDVlMGFmOWM3OTRlMTU3ZGNhY2E5NGE3YjhjOGNhNTI1NmQ1MDNmOGViOnA6RjpG) |
|  | Obesity |  |
|  | TF | NIH, 2025. [https://www.ncbi.nlm.nih.gov/gene/2908](https://protect.checkpoint.com/v2/r01/___https://www.ncbi.nlm.nih.gov/gene/2908___.YzJ1OmJpb3ZpZTE6YzpvOjljNzMyZTM5MjY4ZDg1MDA1MDcyODAxYjc4YTlmZjRhOjc6YzBlOToxODViNjBjNzgwNmJmNmY0YzQ2MTM0ZjUxM2YyZTQ2NGE5ZTRjOTQ3NGE2ZjA2NzU3MTJkYTQwMDdjOWFlMGEwOnA6RjpG) |
|  | Chromatin |  |
|  | NOMINATED** | Agora. [https://agora.adknowledgeportal.org/genes/ENSG00000113580](https://protect.checkpoint.com/v2/r01/___https://agora.adknowledgeportal.org/genes/ENSG00000113580___.YzJ1OmJpb3ZpZTE6YzpvOjljNzMyZTM5MjY4ZDg1MDA1MDcyODAxYjc4YTlmZjRhOjc6MmMzYTpjNzY4OGIwNTI5YTZkZGY0MDAxNWYwMTc4OGIwM2MzMmFmYzc4ZDBiNGZmOTJjMGUyNDI0ZTk0ZmQ1MGEyODI0OnA6RjpG) |
| ***NRBP1*** | Aβ | Yasukawa T et al, 2020. doi: 10.1016/j.celrep.2020.02.059  Yao W et al, 2022. doi: 10.1038/s41380-021-01377-7. Erratum: doi: 10.1038/s41380-021-01417-2 |
|  | Cognition |  |
|  | Microglial neuroinflammation |  |
|  | Significantly ↑ AD Plasma Proteomics | Ali M, et al, 2025. doi: 10.1038/s41591-025-03833-1 Erratum: 2025. doi: 10.1038/s41591-025-03970-7 |
| ***NUBP1*** | Aβ | Yasukawa T et al, 2020. doi: 10.1016/j.celrep.2020.02.059 |
|  | Significantly ↑ AD Plasma Proteomics | Ali M, et al, 2025. doi: 10.1038/s41591-025-03833-1 Erratum: 2025. doi: 10.1038/s41591-025-03970-7 |

| ***OCIAD1*** | Cognition Synapse | Li, X et al, 2020. doi: 10.1016/j.ebiom. 2019.11.030 |
| --- | --- | --- |
|  | Aβ |  |
|  | PTau |  |
|  | BP |  |
|  | Apoptosis |  |
|  | Microglial neuroinflammation | Dongre, P et al, 2025. doi: 10.1186/s12974-025-03415-5 |
| ***OSBPL11*** | Neuro-inflammation | Zhang, H et al, 2025, doi: 10.1007/s10753-025-02252-1 |
|  | AD |  |
|  | Atherosclerosis |  |
|  | Obesity | Guillemot-Legris, O et al, 2016, doi: 10.1038/srep19694 |
|  | Lipids |  |
|  | CYP enzymes |  |
|  | Phosphoprotein | Nhek S et al, 2010, doi: 10.1091/mbc.e10-02-0090 |
| ***P2RX1*** | Aβ toxicity | Godoy P et al, 2019. doi: 10.3389/fphar.2019.01330  Franke H et al, 2007. doi: 10.1007/s11302-00709082-y  Monif M et al, 2009. doi: 10.1523/JNEUROSCI.5512-08.2009 |
|  | Microglial neuroinflammation |  |
|  | Glycolysis | Wang Xu et al, 2020. doi: 10.3389/fimmu.2020.549179 |
|  | M1 polarization | Wang Xu et al, 2021.doi: 10.3389/fimmu.2021.696766 |
|  | Obesity | Cabral-Garcia G et al, 2024. doi: 10.3390/ph17101291 |
| ***PAFAH1B3*** | Neuro-inflammation | Page R et al, 2012. doi: 10.1523/JNEUROSCI.2681-12.2012 |
|  | Aβ |  |
|  | Inflammation | Xie T et al, 2021. doi: 10.3389/fonc.2021.591545 |
|  | M1 polarization |  |
|  | Significantly ↑ AD microglial transcripts with Dementia | Kosoy R et al, 2025. doi: 10.1038/s41593-025-02020-2 |
| ***PAK1*** | Inflammation | Frost J et al, 2000. doi: 10.1074/jbc.M909860199 |
|  | Microglial neuroinflammation | Wilkerson B et al, 2006. doi: 10.1186/1742-2094-3-30 |
|  | AD | Ma Q et al, 2012. doi: 10.4161/cl.21602 |
|  | Cognition |  |
|  | M1 polarization | Zhang W et al, 2014. doi: 10.1038/cdd.2014.142 |
|  | Glycolysis | Murugan S et al, 2025. doi: 10.1016/j.jbc.2025.108409 |
|  | PDHA1 |  |
|  | T2D | Ahn M et al, 2024. doi: 10.1007/s00125-024-06286-2 |
|  | Kinase cascades | Wang Z et al, 2013. doi: 10.1074/jbc.M112.426023 |
|  | Phosphoprotein | King C et al, 2000. doi: 10.1074/jbc.M006553200 |
|  | Lipids | Malecka K et al, 2013. doi: 10.1074/jbc. M112.428904 |
|  | NOMINATED** | [https://agora.adknowledgeportal.org/genes/ENSG00000149269](https://protect.checkpoint.com/v2/r01/___https://agora.adknowledgeportal.org/genes/ENSG00000149269___.YzJ1OmJpb3ZpZTE6YzpvOjljNzMyZTM5MjY4ZDg1MDA1MDcyODAxYjc4YTlmZjRhOjc6OWYxNjozMjM4MGVjN2RkZjBjMzg0MDdjOTgzYWUwNTdlYWZlZjgwM2Q1OWRmNzhiYzI2MzE1NmI0YTNkOTBmOWIwYjNiOnA6RjpG) |
| ***PARD6G-AS1*** | Cognition | van Dongen J et al, 2015. doi: 10.1017/thg.2015.74 |
| ***PBX3*** | TF | Zhang Y et al, 2018. doi: 10.3892/mmr.2018.8609 |
|  | Inflammation |  |
|  | Glycolysis | Wang D et al, 2022. doi: 10.7150/ijbs.69134 |
|  | Kinase cascades | Han H et al, 2014. doi: 10.3748/wjg.v20.i48.18260 |
|  | Lipids | Zhang X et al, 2025. doi: 10.3390/ijms26115210 |
| ***PCK2*** | Cognition | Han J et al, 2021. doi: 10.1016/j.celrep.2021.110102 |
|  | Aβ |  |
|  | M1 polarization | Dong H et al, 2021. doi: 10.3389/fcell.2021.726931 |
|  | Macrophages | Kang Z et al, 2025. doi: 10.3389/fphar.2025.1546045 |
| ***PDE7A*** | Cognition | Zorn A et al, 2023. doi: 10.1016/j.cellsig.2023.110689  Giembycz A et al, 2006. doi: 10.2174/138161206778194123 |
|  | Inflammation |  |
|  | Microglial  neuroinflammation | Ponsaerts L et al, 2021. doi: 10.3390/biomedicines9070703 |
|  | Significantly ↑ AD Plasma Proteomics | Ali M, et al, 2025. doi: 10.1038/s41591-025-03833-1. Erratum: doi: 10.1038/s41591-025-03970-7 |
| ***PDK3*** | Microglial neuroinflammation | Pinky et al, 2025. doi: 10.1016/j.brainres.2025.149476  Wright B et al, 2024. DOI: 10.3390/ijms252111638 |
|  | TNF |  |
|  | M1 polarization | Li C et al, 2023. DOI: 10.3389/fimmu.2023.1296687 |
|  | Glycolysis | Xu J et al, 2019. DOI: 10.7150/thno.31301 |
|  | T2D | Lee I 2014. DOI: 10.4093/dmj.2014.38.3.181 |
|  | Obesity |  |
|  | Lipids | Mayer A et al, 2019. DOI: 10.1126/scisignal.aav9150 |
| ***PDZD8*** | DM | Liu Y et al, 2024. DOI: 10.4093/dmj.2023.0275 |
| ***PEG10*** | Cognition | Mou D et al, 2025. DOI: 10.3892/or.2025.8893 |
|  | Neurodegenerative diseases |  |
|  | Glycolysis | Yin D et al, 2025. DOI: 10.1158/1538-7445.AM2025-5399 |
|  | Obesity | Hishida T et al, 2007. DOI: 10.1016/j.febslet.2007.07.074 |
|  | Phosphoprotein | Abed M et al, 2019. DOI: 10.1371/journal.pone.0214110 |
| ***PEMT*** | DM | Watanabe M et al, 2014. DOI: 10.1371/journal.pone.0092647 |
|  | Obesity |  |
| ***PFKFB4*** | Inflammation | Zhou Y et al, 2022. DOI: 10.2147/IJGM.S369126  Gu X et al, 2023. DOI:10.1038/s41598-023-49212-z  Miller J et al, 1987. DOI: 10.1007/BF01456102 |
|  | Glycolysis |  |
|  | Immune cell infiltration |  |
|  | Obesity | Leiherer A et al, 2016. DOI: 10.3390/nu8050282 |
|  | Microglial neuroinflammation | Vizuete A et al, 2024. DOI: 10.1016/j.bbih.2024.100901  Huang Q et al, 2024. DOI: 10.14336/AD.2023.0807 |
| ***PFKP*** | Glycolysis | Lee J et al, 2017. DOI: 10.1038/s41467-017-00906-9 |
|  | Microglial neuroinflammation | Vizuete A et al, 2024. DOI: 10.1016/j.bbih.2024.100901  Huang Q et al, 2024. DOI: 10.14336/AD.2023.0807 |
|  | T2D | Ausina P et al, 2018. DOI: 10.1016/j.biopha.2018.04.033 |
|  | M1 polarization | Zhu G et al, 2024. DOI: 10.1186/s12931-024-02926-8  Wang S et al, 2022. DOI: 10.3389/fimmu.2022.840029 |
|  | Cognition | Zhang F et al, 2016. DOI: 10.1007/s12264-016-0032-y  Zhang F et al, 2021. DOI: 10.1016/j.neuroscience.2021.01.037 |
|  | Increased AD microglia w/ dementia | Kosoy R et al, 2025. DOI: 10.1038/s41593-025-02020-2 |
|  | Braak* |  |
|  | NOMINATED** | Agora. [https://agora.adknowledgeportal.org/genes/ENSG00000067057](https://protect.checkpoint.com/v2/r01/___https://agora.adknowledgeportal.org/genes/ENSG00000067057___.YzJ1OmJpb3ZpZTE6YzpvOjljNzMyZTM5MjY4ZDg1MDA1MDcyODAxYjc4YTlmZjRhOjc6NDk1YTpkNzIyNjk0ZjcwODEwMDc0NjU3Y2NkMjUwZDRiM2M5OTZjOTY4MzFkZmJmYzBkOGE4Y2M1YjcxZTc4MjRiMGNhOnA6RjpG) |
| ***PGD*** | Neuroinflammation ROS | Martins R et al, 1986. DOI: 10.1111/j.1471-4159.1986.tb00615.x  Palmer A, 1999. DOI: 10.1007/s007020050161 |
|  | Brain peroxides |  |
|  | Vascular inflammation | Lu A et al, 2024. DOI: 10.1016/j.jvssci.2024.100214 |
| ***PGM1*** | Glycolysis | Liu S et al, 2023. DOI: 10.1016/j.bbrc.2023.03.034 |
|  | Phosphorylation | Li Y et al, 2020. DOI: 10.1016/j.canlet.2020.03.007 |
|  | Inhibit glycogen metabolism |  |
|  | Significantly ↑ AD Plasma Proteomics | Ali M, et al, 2025. doi: 10.1038/s41591-025-03833-1. Erratum: doi: 10.1038/s41591-025-03970-7 |
| ***PGPEP1*** | Inflammation | Jiang H et al, 2023. DOI: 10.3389/fimmu.2023.1301539 |
| ***PGRMC1*** | Glycolysis | McGuire M et al, 2021. DOI: 10.1016/j.jbc.2021.101316 |
|  | Lipids | Lee S et al, 2021. DOI:10.1038/s41598-021-88251-2 |
|  | Phosphoprotein | Willibald M et al, 2017. 10.18632/oncotarget.19819 |
|  | Kinase cascade | Pedroza D et al, 2020. DOI: 10.1038/s41416-020-0992-6 |
| ***PHACTR1*** | Microglial,  astrocytic & endothelial neuroinflammation | Jing Y et al, 2022. DOI: 10.4103/1673-5374.357904  Zhang Z et al, 2018. DOI: 10.1016/j.atherosclerosis.2018.08.041 |
|  | NFκB |  |
|  | M1 polarization |  |
|  | Atherosclerosis |  |
| ***PIK3R1*** | Microglial inflammation | Wright B et al, 2024. DOI: 10.3390/ijms252111638  Sanchez-Alegria K et al, 2018. DOI: 10.3390/ijms19123725 |
|  | Cognition |  |
|  | M1 polarization | Linton M et al, 2019. DOI: 10.3390/ijms20112703 |
| ***PIK3R3*** | Microglial neuroinflammation | Wright B et al, 2024. DOI: 10.3390/ijms252111638  Sanchez-Alegria K et al, 2018. DOI: 10.3390/ijms19123725 |
|  | Cognition |  |
|  | M1 polarization | Linton M et al, 2019. DOI: 10.3390/ijms20112703 |
| ***PIK3R5*** | Microglial neuroinflammation | Wright B et al, 2024. DOI: 10.3390/ijms252111638  Sanchez-Alegria K et al, 2018. DOI: 10.3390/ijms19123725 |
|  | Cognition |  |
|  | M1 polarization | Linton M et al, 2019. DOI: 10.3390/ijms20112703 |
| ***PIM3*** | Kinase cascades | Meur S et al, 2024. DOI: 10.1007/s12035-024-04257-7 |
|  | mTOR |  |
|  | Cognition |  |
|  | Inflammation | Clements A, 2022. DOI: 10.3390/cells11223700 |
|  | Immune invasion |  |
|  | T2D | Vlicich G et al, 2010. DOI: 10.4161/isl.2.5.13058 |
|  | Significantly ↑ FTD Plasma Proteomics | Ali M et al, 2025. DOI: 10.1038/s41591-025-03833-1  Erratum: doi: 10.1038/s41591-025-03970-7 |
| ***PIP4K2B*** | T2D | Voss M et al, 2014. DOI: 10.1016/j.bbrc.2014.05.024  Carricaburu V et al, 2003. DOI: 10.1073/pnas.1734038100 |
|  | Obesity | Lamia K et al, 2004. DOI: 10.1128/MCB.24.11.5080-5087.2004 |
|  | Lipids | Lundquist M et al, 2018. DOI: 10.1016/j.molcel.2018.03.037 |
|  | Kinase cascades |  |
| ***PITX1*** | Immune infiltration | Pandey S et al, 2012. DOI: 10.1242/bio.20121305  Overmiller A et al, 2024. DOI: 10.1172/jci.insight.182844 |
|  | TF | UniProt, 2025. [https://www.uniprot.org/uniprotkb/P78337/entry#:~:text=Function-,function,identity%20or%20structure%20of%20hindlimb](https://protect.checkpoint.com/v2/r01/___https://www.uniprot.org/uniprotkb/P78337/entry___.YzJ1OmJpb3ZpZTE6YzpvOjljNzMyZTM5MjY4ZDg1MDA1MDcyODAxYjc4YTlmZjRhOjc6MmQ5Mzo4MjgwOTQ5NDU0YzAxMmY1MGM4MGE4M2U4ZDAxNzMyNGQ1NTAxYjlhMmYwNTczNzZmYzAxZDllN2U2YzY4OThiOnA6RjpG#:~:text=Function-,function,identity%20or%20structure%20of%20hindlimb) |
| ***PKM*** | Cognition | Crary J et al, 2006. DOI: 10.1097/01.jnen.0000218442.07664.04 |
|  | Migroglial neuroinflammation | Zhang Qi et al, 2025. DOI: 10.1038/s41420-025-02453-5 |
|  | M1 polarization |  |
|  | Glycolysis |  |
|  | DM | Tu C et al, 2022. DOI: 10.2147/DMSO.S366403 |
|  | Microangiopathy |  |
|  | Obesity | Rodriguez-Munoz A et al, 2024. DOI: 10.1007/s13679-024-00561-4 |
| ***PKNOX1*** | Down syndrome | Sanchez-Font et al, 2003. DOI: https://doi.org/10.1093/nar/gkg396 |
|  | Lipids |  |
|  | Overexpression |  |
|  | FABP7 |  |
|  | Inflammation | Xu X et al, 2024. DOI: 10.1016/j.clinsp.2024.100354  Zhuang G et al, 2012. DOI: 10.1161/CIRCULATIONAHA.111.087817 |
|  | M1 polarization |  |
|  | Cognition | [https://www.biorxiv.org/content/10.1101/2023.06.21.545934v1](https://protect.checkpoint.com/v2/r01/___https://www.biorxiv.org/content/10.1101/2023.06.21.545934v1___.YzJ1OmJpb3ZpZTE6YzpvOjljNzMyZTM5MjY4ZDg1MDA1MDcyODAxYjc4YTlmZjRhOjc6YTc0ZTo1NWYyOTNjMDAxM2FhMWUyMjk5ZDYwNDIwZDNiN2ExYWY1M2U4NDg1MmU0MGIyOTYyYmU2ZWVlY2I5NmQ3OWE3OnA6RjpG) |
|  | T2D | Ye D et al, 2018. DOI: doi.org/10.1111/jcmm.13902 |
|  | TF | Purushothaman D et al, 2022. DOI: 10.1038/s42003-022-03406-9 |
| ***PLAU*** | Inflammation | Kanno Y, 2023. DOI: 10.3390/ijms24021796 |
|  | Obesity | Alessi M et al, 2007. DOI: 10.1097/MOL.0b013e32814e6d29 |
|  | Kinase cascades | Nguyen D et al, 2000. DOI: 10.1074/jbc.M909575199 |
| ***PLCG2*** | Microglial neuroinflammation | Tsai A et al, 2022. DOI: 10.1186/s13073-022-01022-0 |
|  | Obesity | Dahlman I et al, 2016. doi: 10.2337/db15-0828. |
|  | Significantly ↑ AD Plasma Proteomics | Ali M, et al, 2025. doi: 10.1038/s41591-025-03833-1. Erratum: doi: 10.1038/s41591-025-03970-7 |
|  | NOMINATED** | Agora. [https://agora.adknowledgeportal.org/genes/ENSG00000197943](https://protect.checkpoint.com/v2/r01/___https://agora.adknowledgeportal.org/genes/ENSG00000197943___.YzJ1OmJpb3ZpZTE6YzpvOjljNzMyZTM5MjY4ZDg1MDA1MDcyODAxYjc4YTlmZjRhOjc6ODk0NDozYTk3ZmFmMDI1YzVjYWRhZmM1NmMyODZiYWNlMTYxYTFjM2NkYTUyZTFhYTZlYzYwOWIwYWIyNjViOWI2ZTllOnA6RjpG) |
| ***PLSCR2*** | IFNs | Ma R et al, 2025. DOI: 10.1128/jvi.02085-24  Lundstrom K et al, 2022. DOI:10.1016/j.cellsig.2022.110495  Tsai M et al, 2018. DOI: 10.3389/fimmu.2018.01886 |
|  | COVID-19 | Ma R et al, 2025. DOI: 10.1128/jvi.02085-24 |
|  | Significantly ↑ AD Plasma Proteomics | Ali M, et al, 2025. doi: 10.1038/s41591-025-03833-1. Erratum: doi: 10.1038/s41591-025-03970-7 |
| ***PLXNC1*** | Inflammation | Konig K et al, 2014. DOI: 10.1002/eji.201343968 |
|  | Survival | Granja T et al, 2013. DOI: 10.1038/mi.2013.104 |
|  | Dopaminergic circuits | Chabrat A et al, 2017. DOI: 10.1038/s41467-017-01042-0 |
|  | T2D | Murugesan A et al, 2024. DOI: 10.1186/s12967-024-05928-8 |
| ***POLR2A*** | Macrophage inflammation | Chen Xi et al, 2024. DOI: 10.1126/scitranslmed.adq5091 |
|  | Phosphoprotein | Phatnani H et al, 2006. DOI: 10.1101/gad.1477006 |
|  | TF | Kulaeva O et al, 2007. DOI: 10.1016/j.mrfmmm.2006.05.040 |
|  | Chromatin |  |
| ***POU2F2*** | Glycolysis | Yang R et al, 2021. 10.1038/s41419-021-03719-3 |
|  | Cognition | Masgutova, G et al, 2019. DOI: 10.3389/fnmol.2019.00263  Harris A et al, 2019. DOI: : 10.3389/fncel.2019.00184 |
|  | Interneuron dysfunction |  |
|  | Significantly ↑ FTD Plasma Proteomics | Ali M et al, 2025. DOI: 10.1038/s41591-025-03833-1  Erratum: doi: 10.1038/s41591-025-03970-7 |
|  | Transcription factor | [https://www.ncbi.nlm.nih.gov/gene/5452#:~:text=Summary,the%20germinal%20center%20transcriptional%20program](https://protect.checkpoint.com/v2/r01/___https://www.ncbi.nlm.nih.gov/gene/5452___.YzJ1OmJpb3ZpZTE6YzpvOjljNzMyZTM5MjY4ZDg1MDA1MDcyODAxYjc4YTlmZjRhOjc6ZTI5NDo2MTNlMWJkYjNiODE3ZDJjYjY4MjZhMDdhYjU4NWY1ZGVhZmZhZDc5ZTVmYjIxZDhiYzMwYmFlNzMwMzVkNzY5OnA6RjpG#:~:text=Summary,the%20germinal%20center%20transcriptional%20program) <-- |
| ***PPP1R13L*** | Cognition | Liu N et al, 2021. DOI: 10.1371/journal.pgen.1009363 |
|  | Epigenetic control | Karlsson I et al, 2021. DOI: 10.1186/s13148-021-01075-9 |
|  | Significantly ↑ AD microglial transcripts with Dementia | Kosoy R et al, 2025. DOI: 10.1038/s41593-025-02020-2 |
| ***PPP3CB*** | AD | Norris C, 2023. DOI: 10.3233/JAD-230780 |
|  | Cognition |  |
|  | Inflammation | Cheng Z et al, 2017. DOI: 10.1189/jlb.4A0517-197R |
|  | T2D | Xian L et al, 2015. DOI: 10.18632/aging.100725 |
|  | Kinase cascade | Dougherty M et al, 2009. DOI: 10.1016/j.molcel.2009.06.001 |
|  | Phosphoprotein | Hashmoto Y et al, 1988. DOI: 10.1073/pnas.85.18.7001 |
|  | Astrocytic neuroinflammation | Pfuhlmann K et al, 2018. DOI: 10.1186/s12974-018-1076-x |
|  | Obesity | Pfluger P et al, 2015. DOI: 10.1016/j.cmet.2015.08.022 |
| ***PQBP1*** | Microglial inflammation | Tanaka H et al, 2022. DOI: 10.3390/ijms23116227 |
|  | Aggregation |  |
|  | pTau |  |
|  | Significantly ↑ AD & FTD Plasma Proteomics | Ali M et al, 2025. DOI: 10.1038/s41591-025-03833-1  Erratum: doi: 10.1038/s41591-025-03970-7 |
|  | Chromatin | Koues O et al, 2009. DOI: 10.1016/j.bbagrm.2009.07.006 |
| ***PRDX5*** | M1 polarization | Wu W et al, 2025. DOI: 10.1016/j.intimp.2025.114332 |
|  | Microglial neuroinflammation | Kweon H et al, 2025. DOI: 10.1016/j.atherosclerosis.2024.119052 |
|  | Significantly ↑ AD & PD Plasma Proteomics | Ali M et al, 2025. DOI: 10.1038/s41591-025-03833-1  Erratum: doi: 10.1038/s41591-025-03970-7 |
| ***PRKAB2*** | Glycolysis | Jeon S, 2016. DOI: 10.1038/emm.2016.81 |
|  | T2D |  |
|  | Obesity |  |
|  | M1 polarization | Lei J et al, 2025. DOI: 10.1007/s10753-024-02070-x |
| ***PRKCA*** | Aging cognition | Brennan A et al, 2009. DOI: 0.1016/j.neurobiolaging.2007.08.020 |
|  | M1 polarization | Cheng Y et al, 2018. DOI: 10.1002/mc.22822 |
|  | Obesity | Considine R et al, 1995. DOI: 10.1172/JCI118001 |
|  | Inflammation | Considine R et al, 1995. DOI: 10.1172/JCI118001 |
|  | T2D | Pan D et al, 2022. DOI: 10.3389/fendo.2022.973058 |
|  | Lipids | Schmitz-Peiffer C et al, 2008. DOI: 10.2337/db07-1769 |
|  | NOMINATED** | [https://agora.adknowledgeportal.org/genes/ENSG00000154229](https://protect.checkpoint.com/v2/r01/___https://agora.adknowledgeportal.org/genes/ENSG00000154229___.YzJ1OmJpb3ZpZTE6YzpvOjljNzMyZTM5MjY4ZDg1MDA1MDcyODAxYjc4YTlmZjRhOjc6ZmQ2ZToyYTdiZGIwMTAzZWE2MThhOWE4ZmU3MjI0Mjg0MzhhYjRmM2I3NDgwM2ZjOTlhZWNhNjYxMDMwODgyOTA2ZDUwOnA6RjpG) |
| ***PRKCI*** | Increase BACE & Aβ | Sajan M et al, 2018. DOI: 10.1016/j.neurobiolaging.2017.09.001 |
|  | Kinase cascades | Sarkar A et al, 2017. DOI: 10.1101/gad.296640.117 |
|  | TNF |  |
|  | Significantly ↑ FTD Plasma Proteomics | Ali M et al, 2025. DOI: 10.1038/s41591-025-03833-1  Erratum: doi: 10.1038/s41591-025-03970-7 |
| ***PRKRA*** | Cognition | Hugon J et al, 2017. DOI: 10.1186/s13195-017-0308-0 |
|  | Inhibition of translation |  |
|  | Microglial neuroinflammation |  |
|  | Obesity | Nakamura T et al, 2010. DOI: 10.1016/j.cell.2010.01.001 |
|  | T2D |  |
|  | Significantly ↑ AD Plasma Proteomics | Ali M et al, 2025. DOI: 10.1038/s41591-025-03833-1  Erratum: doi: 10.1038/s41591-025-03970-7 |
| ***PRKRIP1*** | Microglial neuroinflammation | Ofengeim D et al, 2017. DOI: 10;114(41):E8788-E8797 |
|  | Cognition |  |
|  | Aβ |  |
| ***PSMA1*** | Inflammation | Alfaro E et al, 2022. DOI: 10.3390/biom12030442 |
|  | COVID-19 |  |
| ***PSMB1*** | Microglial neuroinflammation | Orre M et al, 2013. DOI: 10.1093/brain/awt083 |
|  | Obesity | Otoda T et al, 2013. DOI: 10.2337/db11-1652  Kitamura H et al, 2023. DOI: 10.3390/ijms24043219 |
|  | T2D |  |
|  | Cognition | Parker D et al, 2025. DOI: 10.1111/acel.14492 |
|  | Microglial neuroinflammation | Orre M et al, 2013. DOI: 10.1093/brain/awt083 |
| ***PSMB10*** | Neurodegeneration | Leister H et al, 2024. DOI: 10.1093/braincomms/fcae017 |
|  | Brain aging | Davidson K et al, 2023. DOI: 10.3389/fcell.2023.1124907 |
|  | Cognition |  |
|  | DM | Thomaidou S et al, 2023. DOI: 10.1007/s00125-023-05991-8 |
|  | Obesity | Fletcher E et al, 2020. DOI: 10.1139/apnm-2020-0655 |
|  | Lipids |  |
| ***PSMB3*** | Inflammation | Sun J et al, 2016. DOI: 10.1016/j.celrep.2015.12.069 |
|  | Cognition | Parker D et al, 2025. DOI: 10.1111/acel.14492 |
| ***PSMB9*** | Vascular inflammation | Li Shu et al, 2024. DOI: 10.1161/CIRCRESAHA.122.322360 |
|  | Atherosclerosis |  |
|  | Microglial neuroinflammation | Orre M et al, 2013. DOI: 10.1093/brain/awt083 |
| ***PSMC5*** | Microglial neuroinflammation | Bi W et al, 2023. DOI: 10.1186/s12974-023-02904-9 |
|  | M1 polarization |  |
|  | Cognition |  |
|  | Significantly ↑ AD & PD Plasma Proteomics | Ali M et al, 2025. DOI: 10.1038/s41591-025-03833-1  Erratum: doi: 10.1038/s41591-025-03970-7  Parker D et al, 2025. DOI: 10.1111/acel.14492 |
| ***PTEN*** | Cognition | Gonzalez M et al, 2021. DOI: 10.3389/fnsyn.2021.683290 |
|  | pTau | Chen Z et al, 2012. DOI: 10.1016/j.ijdevneu.2012.08.003 |
|  | Obesity | Gupta A et al, 2012. DOI: 10.1091/mbc.E12-05-0337 |
|  | Brain IR |  |
|  | Cholesterol inhibition | Kaysudu I et al, 2023. DOI: 10.1111/cas.15960 |
|  | Chromatin | Yang J et al, 2020. DOI: 10.1101/cshperspect.a036160 |
| ***PTGER2*** | Aβ | Wei L et al, 2010. DOI: 10.1007/s12264-010-0703-z |
|  | Hippocampal apoptosis |  |
|  | Microglial neuroinflammation |  |
|  | Lipid peroxidation |  |
|  | Cognition | Jiang C et al, 2020. DOI: 10.1016/j.bbih.2020.100132 |
|  | Lipids | Kaczmarek I et al, 2023. DOI: 10.1016/j.isci.2023.107841 |
|  | BP | Guan Y et al, 2007. doi: <http://dx.doi.org/10.1172/JCI29838> |
|  | Obesity | Civelek E et al, 2022. DOI: 10.1016/j.plefa.2022.102508 |
| ***PTK2B*** | Aβ | Guo Y et al, 2023. DOI: 10.2174/0115672050299004240129051655 |
|  | pTau |  |
|  | Cognition | de Pins B et al, 2021. DOI: 10.3389/fnsyn.2021.749001 |
|  | Glycolysis | Huang T et al, 2025. DOI: 10.1038/s41598-025-88538-8 |
|  | M1 polarization | Lin Y et al, 2023. DOI: 10.1038/s41467-023-43419-4 |
|  | Obesity | Prida E et al, 2024. DOI: 10.1016/j.isci.2024.111120 |
|  | DM | Yu Y et al, 2005. DOI: 10.1016/j.bbrc.2005.06.198 |
|  | Significantly ↑ AD Plasma Proteomics | Ali M et al, 2025. DOI: 10.1038/s41591-025-03833-1  Erratum: doi: 10.1038/s41591-025-03970-7 |
|  | NOMINATED** | [https://agora.adknowledgeportal.org/genes/ENSG00000120899](https://protect.checkpoint.com/v2/r01/___https://agora.adknowledgeportal.org/genes/ENSG00000120899___.YzJ1OmJpb3ZpZTE6YzpvOjljNzMyZTM5MjY4ZDg1MDA1MDcyODAxYjc4YTlmZjRhOjc6NWZhZTphYWNhZDQ2ZDRjMDQ0MWZjOTY3YjkyZmEwMzdjNzc4ZDNlY2RkYmRmODkyNDI3OWU5NDUxYmRmNWE1YjNlNWFhOnA6RjpG) |
| ***PTPN6*** | *CD33* variant | Beckers L et al, 2024. DOI: 10.3390/genes15091204 |
|  | Microglial neuroinflammation | Wang X et al, 2023. DOI: 10.1186/s13195-023-01311-9 |
|  | HHcy |  |
|  | Obesity | Ahn D et al, 2022. DOI: 10.3390/ijms23095020  Gone G et al, 2024. DOI: 10.3390/nu16050647 |
|  | DM |  |
|  | TF | Kumar A et al, 2023. doi: 10.1016/j.jbc.2023.105164 |
|  | Significantly ↑ AD Plasma Proteomics | Ali M et al, 2025. DOI: 10.1038/s41591-025-03833-1  Erratum: doi: 10.1038/s41591-025-03970-7 |
|  | NOMINATED** | Agora. [https://agora.adknowledgeportal.org/genes/ENSG00000111679](https://protect.checkpoint.com/v2/r01/___https://agora.adknowledgeportal.org/genes/ENSG00000111679___.YzJ1OmJpb3ZpZTE6YzpvOjljNzMyZTM5MjY4ZDg1MDA1MDcyODAxYjc4YTlmZjRhOjc6ZmQ3NDo5ZjMwZTgyOTFhMmY3OWNhNDc1MTA4YzM5ZjJkZTEyYzk3MTgzZTZmODZmNjU0M2Y5ZjU2ZGUxZTE2YTJkOGQwOnA6RjpG) |
| ***QRICH1*** | Unfolded protein response | Kumar A et al, 2023. doi: 10.1016/j.jbc.2023.105164  You K et al, 2021. doi: 10.1126/science.abb6896  Wu Y et al, 2025. [https://doi.org/10.32604/biocell.2025.061289](https://protect.checkpoint.com/v2/r01/___https://doi.org/10.32604/biocell.2025.061289___.YzJ1OmJpb3ZpZTE6YzpvOjljNzMyZTM5MjY4ZDg1MDA1MDcyODAxYjc4YTlmZjRhOjc6MmM3MTphZGRhMTQyMmFlYjU1Yjg4OTZkMjU0N2I4OWU4OTIxNDRhNWRmYWEzOGE5MWEzMjU3M2RhZDZhZWJlYjg4ZTY5OnA6RjpG) |
|  | Apoptosis |  |
|  | TF |  |
|  | Kinase cascades |  |
|  | Cognition | Wang S et al, 2025. doi: 10.1016/j.bbadis.2024.167621 |
| ***RAC1*** | Cognition | Wu W et al, 2019. doi: 10.1007/s13238-019-0641-0  Zhang H et al, 2022. doi: 10.3389/fnagi.2022.914491 |
|  | Microglial neuroinflammation | D'Ambrosi N et al, 2014. doi: 10.3389/fncel.2014.00279 |
|  | Glycolysis | Ganapathy-Kanniappan S, 2020. doi: 10.1080/15384047.2020.1809923 |
|  | M1 polarization | Fu H et al, 2023. doi: 10.1038/s41419-023-06150-y |
|  | DM | Kowluru RA et al, 2021. doi: 10.1038/s41598-021-93420-4 |
|  | Obesity | Sun M et al, 2012. doi: 10.1038/oby.2012.63 |
|  | Lipids | Pacia MZ et al, 2022. doi: 10.1007/s00018-022-04362-7  Hasegawa K et al, 2023. doi: 10.3390/ijms24054608 |
|  | Kinase cascades | Manser E et al, 1994. doi: 10.1038/367040a0  Bosco EE et al, 2009. doi: 10.1007/s00018-008-8552-x  Owen D et al, 2003. doi: 10.1074/jbc.M304313200 |
|  | Phosphoprotein | Hou Y et al, 2004. doi: 10.1016/j.cellsig.2004.03.002 |
| ***RAI14*** | Glial inflammation | Shen X et al, 2019. doi: 10.1007/s10571-018-0644-z |
|  | Kinase cascades |  |
|  | NFκB |  |
| ***RBBP4*** | Inflammation | Farjo KM et al, 2012. doi: 10.1128/MCB.00820-12 |
|  | Kinase cascades |  |
|  | NFκB |  |
|  | DM |  |
| ***RBM15*** | Glycolysis | Tang M et al, 2024. doi: 10.7150/ijms.97185 |
|  | M1 polarization |  |
|  | DM | Fang J et al, 2023. doi: 10.1186/s10020-023-00615-8 |
|  | Inflammation | Meng Y et al, 2021. doi: 10.1038/s41419-021-04012-z |
|  | COVID-19 |  |
|  | Lipids | Cai X et al, 2025. doi: 10.1016/j.bbalip.2024.159580 |
|  | Tumorigenesis | Jiang A et al, 2022. doi: 10.1016/j.csbj.2022.08.068 |
|  | Obesity | Shen WB et al, 2022. doi: 10.1016/j.reprotox.2021.12.002 |
| ***RBM23*** | Inflammation | Han H et al, 2021. doi: 10.1155/2021/6697476 |
|  | Aging | Titus MB et al, 2021. doi: 10.1016/j.ydbio.2021.01.011 |
|  | AD |  |
|  | Significantly ↑ AD Plasma Proteomics | Ali M, et al, 2025. doi: 10.1038/s41591-025-03833-1. Erratum: doi: 10.1038/s41591-025-03970-7 |
| ***RFX5*** | TF | Hu Z et al, 2022. doi: 10.1038/s42255-022-00585-x |
|  | Microglial neuroinflammation | Fan Z et al, 2024. doi: 10.1093/gerona/glae031 |
|  | AD |  |
|  | Aβ |  |
| ***RGS2*** | T2D | Vazquez-Jimenez JG et al, 2021. doi: 10.3390/metabo11020091 |
| ***RLIM*** | Obesity | Wang F et al, 2025. doi: 10.1101/2025.07.23.66637 |
|  | Kinase cascades | Yu C et al, 2023. doi: 10.1111/jcmm.17757 |
|  | Phosphoprotein | Huang Y et al, 2022. doi: 10.1038/s41419-021-04493-y |
| ***RNF144B*** | Macrophage priming | Ariffin JK et al, 2016. doi: 10.1189/jlb.2AB0815-339R |
| ***RNF31*** | Neuro-inflammation | Li S et al, 2021. doi: 10.1016/j.cbi.2021.109623 |
|  | NFκB |  |
|  | M1 polarization | Zhang Y et al, 2024. doi: 10.1016/j.lfs.2024.122893 |
| ***RNF4*** | TNF-α | Shimada T et al, 2021. doi: 10.3390/ijms22115796 |
|  | NRF2 | Komaravelli N et al, 2017. doi: 10.1016/j.freeradbiomed.2017.10.380 |
|  | Chromatin | Galanty Y et al, 2012. doi: 10.1101/gad.188284 |
| ***RNY3*** | Macrophage | Clancy RM et al, 2010. doi: 10.4049/jimmunol.0902248 |
|  | Significantly ↑ AD microglial transcripts with Dementia | Kosoy R. et al, 2025. doi: 10.1038/s41593-025-02020-2 |
|  | Braak* |  |
| ***RUNX2*** | Cognition | Nakatsu D et al, 2023. doi: 10.1016/j.stemcr.2023.01.004. Erratum: doi: 10.1016/j.stemcr.2023.03.017 |
|  | Senescence |  |
|  | TF | NIH, 2025. [https://www.ncbi.nlm.nih.gov/gene/860](https://protect.checkpoint.com/v2/r01/___https://www.ncbi.nlm.nih.gov/gene/860___.YzJ1OmJpb3ZpZTE6YzpvOjljNzMyZTM5MjY4ZDg1MDA1MDcyODAxYjc4YTlmZjRhOjc6MWYyNDo5N2YzYjkwMzI0OGU2OTcwZDBkOTQ4MTAxYjc3ZDkyMzcxZDc0ZmE5Y2NlODA5MTM0NTExYjA1YzY3N2RiNjlkOnA6RjpG) |
|  | Chromatin | Lian JB et al, 2003. [https://doi.org/10.1080/03008200390152232](https://protect.checkpoint.com/v2/r01/___https://doi.org/10.1080/03008200390152232___.YzJ1OmJpb3ZpZTE6YzpvOjljNzMyZTM5MjY4ZDg1MDA1MDcyODAxYjc4YTlmZjRhOjc6MGIyZDo2Y2Y4MzMxYjYzZmFlOWJjOWIwYTA4YTY5NGJiM2JmMmI4OTA1Yzg2NmFiMzc1ZGFhNzIwNjA2Y2Y2ODQwMzg4OnA6RjpG) |
| ***SAT1*** | Cognition | Dang Y et al, 2022. doi: 10.3390/ph15101177 |
|  | AD |  |
|  | Ferroptosis |  |
|  | Lipids | Stockwell BR et al, 2017. doi: 10.1016/j.cell.2017.09.021  Ou Y et al, 2016. doi: 10.1073/pnas.1607152113 |
|  | Vascular inflammation | Chmielewski S et al, 2016. doi: 10.3109/08830185.2015.1087519 |
|  | IR | Yuan F et al, 2018. doi: 10.1016/j.metabol.2018.04.007 |
| ***SAT2*** | Inflammation | Bevers MB et al, 2019. doi: 10.1002/ana.25545  Duodu P et al, 2022. doi: 10.1089/jir.2022.0117. |
|  | Neurovascular injury |  |
|  | Cognition | Dandan-Zong et al, 2023. doi: 10.1016/j.intimp.2022.109604 |
|  | Significantly ↑ AD Plasma Proteomics | Ali M, et al, 2025. doi: 10.1038/s41591-025-03833-1. Erratum: doi: 10.1038/s41591-025-03970-7 |
| ***SCO2*** | Glycolysis | Gujarati NA et al, 2021. doi: 10.2337/db21-0316 |
|  | ROS |  |
|  | T2D |  |
|  | Phosphoprotein | Miyazaki T et al, 2003. doi: 10.1083/jcb.200209098 |
| ***SELPLG*** | Atherosclerosis | Kisucka J et al, 2009. doi: 10.1182/blood-2008-10-186650 |
|  | BBB permeability |  |
|  | Inflammation | Ley K, 2003. doi: 10.1016/s1471-4914(03)00071-6 |
|  | Microglial neuroinflammation | D'Mello C et al, 2013. doi: 10.1523/JNEUROSCI.1329-13.2013 |
|  | Peripheral organ inflammation |  |
|  | IR | Sato C et al, 2011. doi: 10.2337/db09-1894 |
|  | Obesity |  |
|  | Kinase cascades | Hidari KI et al, 1997. doi: 10.1074/jbc.272.45.28750 |
|  | Phosphoprotein | Hirata T et al, 2004. doi: 10.1074/jbc.M409868200 |
|  | Lipids | Needham LK et al, 1993. doi: 10.1073/pnas.90.4.1359 |
| ***SERBP1*** | Neuronal cell regulation | Barbato C et al, 2022. doi: 10.3390/cells11061052 |
|  | Cognition |  |
|  | Phosphoprotein | Martini S et al, 2021. doi: 10.1038/s41467-021-27189-5 |
| ***SERPIN6*** | Dementia | Lomas DA, Carrell RW, 2002. doi: 10.1038/nrg907 |
|  | Significantly ↑ AD microglial transcripts with Dementia | Kosoy R. et al, 2025. doi: 10.1038/s41593-025-02020-2 |
|  | Significantly ↑ FTD Plasma Proteomics | Ali M, et al, 2025. doi: 10.1038/s41591-025-03833-1. Erratum: doi: 10.1038/s41591-025-03970-7 |
| ***SH2B3*** | Inflammation | Dale BL et al, 2016. doi: 10.1097/MNH.0000000000000196 |
|  | HTN |  |
|  | Neuronal differentiation | Wang TC et al, 2011. doi: 10.1371/journal.pone.0026433 |
| ***SF3A2/ SF3A3*** | Inflammation | De Arras L, Alper S, 2013. doi: 10.1371/journal.pgen.1003855 |
|  | M1 polarization |  |
| ***SGK3*** | Cognition | Lang UE et al, 2006. doi: 10.1016/j.bbr.2005.08.017 |
|  | Chromatin | Chen Q et al, 2025. doi: 10.1172/JCI186534 |
|  | Phosphoprotein |  |
|  | Obesity |  |
|  | Lipids |  |
|  | Kinase cascades |  |
| ***SH3GLB2*** | Cognition | Lee MH et al, 2017. doi: 10.1016/j.trci.2017.02.001 |
|  | Neuron death |  |
|  | Aβ |  |
|  | Microglial neuroinflammation |  |
|  | NOMINATED** | Agora. [https://agora.adknowledgeportal.org/genes/ENSG00000148341](https://protect.checkpoint.com/v2/r01/___https://agora.adknowledgeportal.org/genes/ENSG00000148341___.YzJ1OmJpb3ZpZTE6YzpvOjljNzMyZTM5MjY4ZDg1MDA1MDcyODAxYjc4YTlmZjRhOjc6YzFhNDpmZmFkNjU0YTAzZTlkODQyZGQwOTZjOWM3YTE4NjNlMDExMjhhNDkxNzNlNTUxYmNhNzQ4OWVmZGZmMWQxMjBkOnA6RjpG) |
| ***SH3BGRL2*** | T2D | Gu X et al, 2020. doi: 10.1186/s12911-020-01223-w  Collares CVA et al, 2013. doi: 10.1007/s11033-013-2635-y |
|  | Significantly ↑ AD Plasma Proteomics | Ali M, et al, 2025. doi: 10.1038/s41591-025-03833-1. Erratum: doi: 10.1038/s41591-025-03970-7 |
| ***SIRT2*** | Cognition | Bai N et al, 2022. doi: 10.1016/j.celrep.2022.111062 |
|  | Neuro-inflammation | Lu W et al, 2023. doi: 10.3389/fimmu.2023.1174180 |
|  | M1 polarization | Yuan K et al, 2025. doi: 10.1007/s12975-024-01282-5 |
|  | T2D | Arora A et al, 2014. doi: 10.1016/j.bbadis.2014.04.027 |
|  | Chromatin | Malgulwar PB et al, 2024. doi: 10.1093/neuonc/noad155 |
|  | Significantly ↑ AD Plasma Proteomics | Ali M, et al, 2025. doi: 10.1038/s41591-025-03833-1. Erratum: doi: 10.1038/s41591-025-03970-7 |
| ***SLC15A4*** | Inflammation | Chiu TY et al, 2024. doi: 10.1038/s41589-023-01527-8 |
|  | M1 polarization | Kobayashi T et al, 2021. doi: 10.1073/pnas.2100295118 |
| ***SLC1A5*** | Cognition | Zhao X et al, 2019. doi: 10.1111/acel.12947 |
|  | Aging |  |
|  | Lipids |  |
|  | Inflammation | Chen P et al, 2023. doi: 10.18632/aging.204911 |
|  | Ferroptosis |  |
|  | Glutamine homeostasis | Bhutia YD et al, 2016. doi: 10.1016/j.bbamcr.2015.12.017  [https://www.researchgate.net/publication/390286460_SLC1A5_Structure_Function_and_Clinical_Significance](https://protect.checkpoint.com/v2/r01/___https://www.researchgate.net/publication/390286460_SLC1A5_Structure_Function_and_Clinical_Significance___.YzJ1OmJpb3ZpZTE6YzpvOjljNzMyZTM5MjY4ZDg1MDA1MDcyODAxYjc4YTlmZjRhOjc6NDI5MDo3N2ZmMDYzMmM2OTI0NTRjZDFjNDdlYTM4YzU0OGY2NmI3ODA3NDFmNDFlMjAxNjAyMTMxYWQyMzU2ZjQ2ODYxOnA6RjpG) |
|  | Excitotoxicity |  |
|  | Neuro-degeneration | James S et al, 1990. doi: 10.1016/0006-8993(90)91090-4 |
|  | Microglial neuroinflammation | Shi AC et al, 2021. doi: 10.3389/fneur.2020.626999 |
|  | NOMINATED** | Agora. [https://agora.adknowledgeportal.org/genes/ENSG00000105281](https://protect.checkpoint.com/v2/r01/___https://agora.adknowledgeportal.org/genes/ENSG00000105281___.YzJ1OmJpb3ZpZTE6YzpvOjljNzMyZTM5MjY4ZDg1MDA1MDcyODAxYjc4YTlmZjRhOjc6OTNkZDo1Njk0OGMyZWE5MTcyYTZhMmI0ZjNkYmZjM2IzYzUwNjkzMTlmNzYwMDYxMjc0YWQyZDI3YTUyOTZkN2VmYTAwOnA6RjpG) |
| ***SLC25A12*** | Cognition | Lepagnol-Bestel AM et al, 2008. doi: 10.1038/sj.mp.4002120 |
|  | Neuronal networks |  |
| ***SLC28A2-AS1*** | Cognition | Ayka A et al, 2020. doi: 10.9758/cpn.2020.18.2.174  Neutzner M et al, 2023. doi: 10.1186/s12987-023-00412-9. |
| ***SLC43A3*** | Inflammation | Hasbargen KB et al, 2020. doi: 10.1194/jlr.RA119000294 |
|  | FFA |  |
| ***SLC9A7*** | Cognition | Meda SA et al, 2012. doi: 10.1016/j.neuroimage.2011.12.076 |
|  | AD |  |
|  | Significantly ↑ AD microglial transcripts with Dementia | Kosoy R et al, 2025. DOI: 10.1038/s41593-025-02020-2 |
|  | Braak* |  |
| ***SLC9B2***  *Decreased promoter DNAm (potentially increased* *expression) in bezisterim subjects vs placebo* | Aging | Deisl C et al, 2016. doi: 10.1371/journal.pone.0163568 |
|  | IR |  |
|  | Obesity |  |
|  | Skeletal metabolism | Creative Biolabs. 2025. [https://www.creative-biolabs.com/slc9b2-membrane-protein-introduction.html](https://protect.checkpoint.com/v2/r01/___https://www.creative-biolabs.com/slc9b2-membrane-protein-introduction.html___.YzJ1OmJpb3ZpZTE6YzpvOjljNzMyZTM5MjY4ZDg1MDA1MDcyODAxYjc4YTlmZjRhOjc6ZmMzNTo3YzUyNGI2MjRmNjYyMmE3YmJiZTBjYTQ4OGI0MjUyOTQwMjIzNjAzOTBlMDEyYzlhMmQwMzJjYjYwMzI4ZTkzOnA6RjpG)  Ho TM et al, 2022. doi: 10.2533/chimia.2022.1019 |
|  | HTN |  |
|  | DM |  |
|  | M1 polarization | Anderegg MA et al, 2022. doi: 10.3389/fphys.2022.898508 |
|  | Significantly ↑ AD microglial transcripts with Dementia | Kosoy R et al, 2025. DOI: 10.1038/s41593-025-02020-2 |
|  | Braak* |  |
| ***SLU7*** | AD | Biamonti G et al, 2021. doi: 10.1007/s40520-019-01360-x  Angarola BL, Anczuków O, 2021. doi: 10.1002/wrna.1643 |
|  | Splicing |  |
|  | Inflammation | Wang J et al, 2018. doi: 10.1016/j.ajpath.2018.05.004 |
|  | Cognition | Li D et al, 2024. doi: 10.1172/JCI171235 |
| ***SMAD1*** | Cognition | Nakatsu D et al, 2023. doi: 10.1016/j.stemcr.2023.01.004. Erratum: doi: 10.1016/j.stemcr.2023.03.017 |
|  | AD neurogenesis |  |
|  | Inflammation | Rosendahl A et al, 2002. doi: 10.1165/ajrcmb.27.2.4779  Besson-Fournier C et al, 2012. doi: 10.1182/blood-2012-02-411470 |
|  | Obesity | Blázquez-Medela AM et al, 2019. doi: 10.1111/obr.12822 |
|  | Lipids |  |
|  | Phosphoprotein | Ramachandran A et al, 2018. doi: 10.7554/eLife.31756 |
|  | TF |  |
|  | Chromatin | Itoh Y et al, 2019. doi: 10.1074/jbc.RA119.009877 |
| ***SMAD3*** | Cognition | Xu L et al, 2021. doi: 10.1016/j.bbi.2021.03.013 |
|  | Aβ |  |
|  | Neutrophils | Chung JY et al, 2023. doi: 10.1038/s41467-023-37515-8 |
|  | Inflammation | Ashcroft GS et al, 1999. doi: 10.1038/12971 |
|  | Glycolysis | Cai Y et al, 2025. doi: 10.1186/s13287-025-04541-w |
|  | DM | Yadav H et al, 2011. doi: 10.1016/j.cmet.2011.04.013 |
|  | Obesity |  |
|  | Kinase cascades | Tzavlaki K, Moustakas A, 2020. doi: 10.3390/biom10030487 |
|  | Phosphoprotein |  |
|  | Lipids | Yi Y et al, 2023. doi: 10.1016/j.devcel.2023.07.005 |
|  | TF | Ross S et al, 2006. doi: 10.1038/sj.emboj.7601332 |
|  | Chromatin |  |
|  | NOMINATED** | Agora. [https://agora.adknowledgeportal.org/genes/ENSG00000166949](https://protect.checkpoint.com/v2/r01/___https://agora.adknowledgeportal.org/genes/ENSG00000166949___.YzJ1OmJpb3ZpZTE6YzpvOjljNzMyZTM5MjY4ZDg1MDA1MDcyODAxYjc4YTlmZjRhOjc6MDMyMzozYmJjMDBlZGIyZGZkNjhkZDk5YzNlZDY1Y2Q1MDhiMTA2MTUxYzEwNjQ3OTEwMjJjYzIzZTA3N2JlZjdhMmU4OnA6RjpG) |
| ***SMARCC1*** | Inflammation | Gatchalian J et al, 2020. doi: 10.1016/j.it.2019.12.002  Church MC et al, 2021. doi: 10.3390/ijms221910274 |
|  | Aging |  |
|  | Lipids | Sun J et al, 2023. doi: 10.1016/j.isci.2023.108207 |
|  | M1 polarization | Gatchalian J et al, 2020. doi: 10.1016/j.it.2019.12.002 |
|  | Obesity | Jeon J et al, 2024. doi: 10.3390/ijms252111681  Lee YS et al, 2007. doi: 10.1128/MCB.00490-06 |
|  | Chromatin |  |
|  | Significantly ↑ PD Plasma Proteomics | Ali M, et al, 2025. doi: 10.1038/s41591-025-03833-1. Erratum: doi: 10.1038/s41591-025-03970-7 |
| ***SMARCD2*** | Inflammation | Foster KS et al, 2006. doi: 10.1038/sj.onc.1209496  Gullett JM et al, 2022. doi: 10.1038/s41422-022-00688-w |
|  | Epigenetic methylation |  |
|  | Chromatin | Priam P et al, 2024. doi: 10.1016/j.devcel.2024.08.007 |
| ***SMC3*** | Aging | Chen Z et al, 2019. doi: 10.1084/jem.20181505 |
|  | Inflammation |  |
|  | NFκB |  |
|  | Chromatin | Weill Cornell Medicine, 2021. [https://news.weill.cornell.edu/news/2021/02/scientists-identify-gene-that-plays-critical-role-in-immune-cell-development#:~:text=Much%20of%20the%20three%2Ddimensional,and%20become%20prone%20to%20malignancy](https://protect.checkpoint.com/v2/r01/___https://news.weill.cornell.edu/news/2021/02/scientists-identify-gene-that-plays-critical-role-in-immune-cell-development___.YzJ1OmJpb3ZpZTE6YzpvOjljNzMyZTM5MjY4ZDg1MDA1MDcyODAxYjc4YTlmZjRhOjc6ZjFjYjpiY2ZlMjJhYWE0N2VmNzk4NjRiYzc1ZDEyYTE4ODg2MGE4NDM2MzQ4M2M0MGQ3OTljNGI3MGM1NjIzOTM4YTJhOnA6RjpG#:~:text=Much%20of%20the%20three%2Ddimensional,and%20become%20prone%20to%20malignancy) |
|  | NOMINATED** | Agora. [https://agora.adknowledgeportal.org/genes/ENSG00000108055](https://protect.checkpoint.com/v2/r01/___https://agora.adknowledgeportal.org/genes/ENSG00000108055___.YzJ1OmJpb3ZpZTE6YzpvOjljNzMyZTM5MjY4ZDg1MDA1MDcyODAxYjc4YTlmZjRhOjc6YTdlNTpmZmQyYmQ1MjI3OTAwYzg2NmM5OGJlOTAxZThmZTQ2NTRjYmJlMzc0YTRlYTY0N2I0NTY1NjUyZTIyODc4ZjdiOnA6RjpG) |
| ***SNRPA*** | Cognition | Chen PC et al, 2022. doi: 10.1038/s43587-022-00290-0 |
|  | Splicing |  |
|  | Neuro-inflammation | Bai B, 2018. doi: 10.3389/fnagi.2018.00075 |
|  | Cell cycle |  |
|  | Significantly ↑ AD Plasma Proteomics | Ali M, et al, 2025. doi: 10.1038/s41591-025-03833-1. Erratum: doi: 10.1038/s41591-025-03970-7 |
| ***SOAT1*** | Lipids | Huang L et al, 2025. doi: 10.1016/j.bbi.2025.04.032. Erratum: doi: 10.1016/j.bbi.2025.06.008.  Huynh TN et al, 2024. doi: 10.3390/ijms252413690 |
|  | Microglial neuroinflammation |  |
|  | BBB dysfunction |  |
|  | Aging |  |
|  | M1 polarization | Peng P et al, 2025. doi: 10.1007/s11010-025-05246-7 |
|  | T2D | Liu X et al, 2020. doi: 10.1016/j.kint.2020.06.040 |
|  | Obesity | Kim SQ, 2023. [https://hammer.purdue.edu/articles/thesis/The_Role_of_Sterol_O-acyltransferase_1_In_Obesity_And_In_Prostate_Cancer/22704412?file=40356370](https://protect.checkpoint.com/v2/r01/___https://hammer.purdue.edu/articles/thesis/The_Role_of_Sterol_O-acyltransferase_1_In_Obesity_And_In_Prostate_Cancer/22704412?file=40356370___.YzJ1OmJpb3ZpZTE6YzpvOjljNzMyZTM5MjY4ZDg1MDA1MDcyODAxYjc4YTlmZjRhOjc6NTk4MjowYTRhYmQxMjM1Y2I5ZmZjYzEyYzRhMGIxN2I4MzZhZjg3MTFkM2EyY2U4YmJkZjg3NGU0OTlkYzE4MTU4ZjJhOnA6RjpG)  Liu Q et al, 2024. doi: 10.1016/j.jlr.2024.100680 |
| ***SOD2*** | Inflammation | Yoon Y et al, 2018. doi: 10.1111/odi.12933 |
|  | DNA Methylation | Hurt EM et al, 2007. doi: 10.1038/sj.bjc.6604000  Ishihara Y et al, 2015. doi: 10.1074/jbc.M115.659151 |
| ***SP140***  *SP140 promoter DNA methylation was decreased in bezisterim* *subjects compared to placebo (potential anti-inflammatory increase in expression)* | Chromatin | Fraschilla I et al, 2020. doi: 10.1016/j.it.2020.04.007 |
|  | Microglial neuroinflammation | Saddala MS et al, 2021. doi: 10.1016/j.ygeno.2021.07.001 |
|  | NF-κB | Karaky M et al, 2018. doi: 10.1093/hmg/ddy284 |
|  | AD | Citron BA et al, 2015. www.AJND.us /ISSN:2165-591X/AJND0020388 |
|  | Cognition |  |
|  | Significantly ↓ AD microglial transcripts with Dementia | Kosoy R et al, 2025. DOI: 10.1038/s41593-025-02020-2 |
|  | Braak* |  |
| ***SP2*** | Cholesterol | Terrados G et al, 2012. doi: 10.1093/nar/gks544 |
|  | Lipids |  |
|  | TF |  |
|  | Inflammation | Zschemisch NH et al, 2016. doi: 10.1371/journal.pone.0155821  Ilarregui JM et al, 2016. doi: 10.1038/icb.2016.66  Mills KHG, 2023. doi: 10.1038/s41577-022-00746-9  Bidgood GM et al, 2024. doi: 10.3389/fimmu.2024.1419951 |
|  | M1 polarization | Liu B et al, 2025. doi: 10.1186/s40001-025-02947-z |
| ***SP3*** | AD | Yamakawa H et al, 2017. doi: 10.1016/j.celrep.2017.07.044  Pang X et al, 2025. doi: 10.1007/s12017-025-08844-2 |
|  | TF |  |
|  | Cognition |  |
|  | Microglial neuroinflammation | Pang X et al, 2025. doi: 10.1007/s12017-025-08844-2  Beug ST et al, 2019. doi: 10.1126/scisignal.aat9563 |
|  | M1 polarization |  |
|  | NFκB |  |
|  | Kinase cascades |  |
|  | Chromatin | Stielow B et al, 2008. doi: 10.1038/embor.2008.127 |
| ***SQSTM1*** | M1 polarization | Yang W et al, 2022. doi : <https://doi.org/10.1177/1721727X2211103> |
|  | Inflammation |  |
|  | NFκB |  |
|  | Kinase cascades |  |
|  | Glycolysis | Chen K et al, 2016. doi: 10.1242/jcs.178756 |
|  | Phosphoprotein | Matsumoto G et al, 2011. doi: 10.1016/j.molcel.2011.07.039 |
| ***SSBP4*** | Cognition | Zarrella JA et al, 2024. doi: 10.18632/aging.205609 |
| ***ST6GAL1*** | Glycolysis | Moll T et al, 2020. doi: 10.1093/brain/awz358  Kang Y et al, 2024. doi: 10.3389/fnagi.2024.1398641  Yang K et al, 2025. DOI: [10.1016/j.eng.2025.02.016](https://protect.checkpoint.com/v2/r01/___https://www.engineering.org.cn/engi/EN/10.1016/j.eng.2025.02.016___.YzJ1OmJpb3ZpZTE6YzpvOjljNzMyZTM5MjY4ZDg1MDA1MDcyODAxYjc4YTlmZjRhOjc6ZDYyZjo5MDA0Y2YxOGRkYjVjZWZhNDUzNDQ5NGE2MDNkMzdiZjdiYTJkNjNkM2I5OWM5OTNkYWE5Y2MwNjUwMjhkNDA3OnA6RjpG) |
|  | Neurodegeneration |  |
|  | Aβ |  |
|  | Inflammation | Holdbrooks AT et al, 2020. doi: 10.1371/journal.pone.0241850 |
|  | Significantly ↑ AD Plasma Proteomics | Ali M, et al, 2025. doi: 10.1038/s41591-025-03833-1. Erratum: doi: 10.1038/s41591-025-03970-7 |
| ***STARD3*** | Lipids | Wilhelm LP et al, 2017. doi: 10.15252/embj.201695917 |
|  | DM | Hu J et al, 2024. doi: 10.1016/j.lfs.2024.122722 |
|  | Obesity | Zhou X et al, 2018. doi: 10.1016/j.bbrc.2018.06.030 |
|  | ROS |  |
| ***STAT3*** | Neuro-inflammation | Wen X et al, 2024. doi: 10.1016/j.intimp.2024.112936  Wang Y et al, 2022. doi: 10.3389/fncel.2022.980722  Millot P et al, 2020. doi: 10.1016/j.imlet.2020.10.004 |
|  | Aβ | Millot P et al, 2020. doi: 10.1016/j.imlet.2020.10.004  Reichenbach N et al, 2019. doi: 10.15252/emmm.201809665 |
|  | Cognition |  |
|  | M1 polarization | Chen X et al, 2022. doi: 10.1590/1678-7757-2022-0316 |
|  | Glycolysis | Li M et al, 2017. doi: 10.18632/oncotarget.15801 |
|  | T2D | Mashili F et al, 2013. doi: 10.2337/db12-0337 |
|  | Obesity | Wunderlich CM et al, 2013. doi: 10.4161/jkst.23878 |
|  | TF | Egwuagu CE, 2009. doi: 10.1016/j.cyto.2009.07.003 |
|  | Chromatin | Wingelhofer B et al, 2018. doi: 10.1038/s41375-018-0117-x. |
|  | Significantly ↑ AD & PD Plasma Proteomics | Ali M, et al, 2025. doi: 10.1038/s41591-025-03833-1. Erratum: doi: 10.1038/s41591-025-03970-7 |
|  | NOMINATED** | Agora. [https://agora.adknowledgeportal.org/genes/ENSG00000168610](https://protect.checkpoint.com/v2/r01/___https://agora.adknowledgeportal.org/genes/ENSG00000168610___.YzJ1OmJpb3ZpZTE6YzpvOjljNzMyZTM5MjY4ZDg1MDA1MDcyODAxYjc4YTlmZjRhOjc6OWM3MToyYWJiNmRhMTU3ZjkzMWQ5ZDEyZjJhZGNlZmMwZmRlOGY1NjZiMzIyOThmZTVmZGFlMzk4Yzk0M2RhMzljZGMxOnA6RjpG) |
| ***STAT5A*** | Microglial neuroinflammation | Conte F et al, 2022. doi: 10.1038/s41598-022-20404-3 |
|  | M1 polarization | Jesser EA et al, 2021. doi: 10.1186/s13058-021-01481-0 |
|  | Glycolysis | Zhang L et al, 2021. doi: 10.1038/s41419-021-03908-0 |
|  | Chromatin | Wingelhofer B et al, 2018. doi: 10.1038/s41375-018-0117-x. |
|  | TF | Hennighausen L et al, 2008. doi: 10.1101/gad.1643908 |
|  | Cytokines |  |
| ***STAU1*** | Microglial neuroinflammation | Zhong Y et al, 2020. doi: 10.3892/or.2020.7769 |
|  | pTau |  |
|  | Aβ |  |
|  | M1 polarization |  |
|  | Kinase cascades |  |
|  | Apoptosis | Gandelman M et al, 2020. doi: 10.1038/s41418-020-0553-9. Erratum: doi: 10.1038/s41418-021-00734-x |
|  | Unfolded protein response |  |
|  | Autophagy | Zhao R et al, 2024. doi: 10.1083/jcb.202311127 |
|  | Obesity | Jiang S et al, 2023. doi: 10.1016/j.bbalip.2023.159293. |
|  | NOMINATED** | Agora. [https://agora.adknowledgeportal.org/genes/ENSG00000124214](https://protect.checkpoint.com/v2/r01/___https://agora.adknowledgeportal.org/genes/ENSG00000124214___.YzJ1OmJpb3ZpZTE6YzpvOjljNzMyZTM5MjY4ZDg1MDA1MDcyODAxYjc4YTlmZjRhOjc6OTU1MjpmMzY5YmVhOWU3MmJkZmE5NmE5NjFiN2IxMjUxNjQ3MWZjYjI0OTQ0OGMwY2RiNzAyZmNhYzk0OTU0YTM0NDI2OnA6RjpG) |
| ***TAF1*** | Transcription | Crombie EM et al. 2024. doi: 10.1098/rsos.240790. |
|  | Neurodegeneration |  |
| ***TAF7*** | Transcription | TAF7 TATA-box binding protein associated factor 7 [Homo sapiens (human)]. Gene ID:6879. Updated 19 Aug 2025. |
|  | HAT regulation |  |
| ***TAF4B*** | Transcription | Shipper, M, 2013. https://doi.org/10.17192/z2013.0077 |
|  | Chromatin |  |
|  | Inflammation | National Center for Biotechnology Information, 2026. <https://www.ncbi.nlm.nih.gov/gene/6875#:~:text=Summary,provided%20by%20RefSeq%2C%20Jun%202014%5D>  Gura MA, et al, 2022. doi: 10.1242/dev.200074 |
|  | TNF-α |  |
|  | NFκB |  |
| ***TAF9*** | Transcription | Buss H et al. 2004. doi: 10.1074/jbc.M409825200.  Liu X et al. 2008. doi: 10.1128/MCB.01402-07.  Fan W, et al. 2017. doi: 10.1371/journal.pgen.1006664. |
|  | Inflammation |  |
|  | Lipids |  |
| ***TBC1D14*** | Neuroinflammation | Li QS et al. 2021. doi: 10.1016/j.bbih.2021.100227. |
|  | Autophagy | Longatti A et al. 2012. doi: 10.1083/jcb.201111079.  Lu T et akl. 2022. doi: 10.7150/ijbs.68992. |
| ***TBP*** | TATA-box binding protein | Reid SJ et al. 2004. doi: 10.1016/j.molbrainres.2004.03.018.  Hardivillé S et al, 2020. doi: 10.1016/j.molcel.2019.11.022 |
|  | Neuroinflammation |  |
|  | Lipids |  |
|  | Inflammation | Tjitro R et al. 2019. doi: 10.3389/fimmu.2018.03110. |
|  | Cognition | Fujigasaki H et al. 2001. doi: 10.1093/brain/124.10.1939 |
|  | Kinase cascades | Chibazakura T et al, 1997. doi: 10.1111/j.1432-1033.1997.01166.x |
|  | Phosphoprotein | Maldonado E, Allende JE, 1999. doi: 10.1016/s0014-5793(98)01734-7 |
|  | TF | Ravarani CNJ et al, 2020. doi: 10.1038/s41467-020-16182-z |
|  | Chromatin | Lomvardas S, Thanos D, 2001. doi: 10.1016/s0092-8674(01)00490-1 |
| ***TCF12*** | Cognition | Wu K et al. 2012. doi: 10.1016/j.nlm.2011.09.006 |
|  | Inflammation | Zheng H et al. 2023. doi: 10.1016/j.jot.2023.11.006 |
|  | M1 polarization | Tampella G. 2015. doi: 10.4049/jimmunol.1403238 |
|  | DM | Thakar S et al. 2025. doi: 10.1016/j.ijbiomac.2025.144963 |
|  | Chromatin |  |
| ***TEF*** | Apoptosis | Ritchie A. 2009. doi: 10.1038/cdd.2009.13  Anbanandam. 2006. doi: 10.1073/pnas.0607171103. |
| ***TERF2IP*** | Inflammation | Le NT et al, 2017. [https://doi.org/10.1161/atvb.37.suppl_1.41](https://protect.checkpoint.com/v2/r01/___https://doi.org/10.1161/atvb.37.suppl_1.41___.YzJ1OmJpb3ZpZTE6YzpvOjljNzMyZTM5MjY4ZDg1MDA1MDcyODAxYjc4YTlmZjRhOjc6NTY3MjpkYmNiNTdkMGU2ZDdjODkzYWEyNGVhMWY3YTE2ZmUyNDgyMjBkYzQyYTRhNjhmYjEzYjY3ZWJhOTdkNDVkYmUxOnA6RjpG) |
|  | Kinase cascades |  |
|  | NFκB |  |
|  | Phosphoprotein |  |
|  | Senescence |  |
|  | Cognition |  |
|  | Aβ | Wu Q et al, 2019. doi: 10.1080/01616412.2019.1580456 |
|  | pTau |  |
|  | Lipids | Fan W et al, 2017. doi: 10.1371/journal.pgen.1006664 |
|  | TF | UniProt, 2025. https://www.uniprot.org/uniprotkb/Q91VL8/entry#:~:text=Acts%20both%20as%20a%20regulator%20of%20telomere,telomere%20length%20and%20protection%20as%20a%20component |
| ***TF*** | AD | Petralla S et al. 2024. doi: 10.1007/s12035-024-03990-3 |
|  | Oxidative stress |  |
|  | Inflammation |  |
|  | Cognition | Guan J et al. 2020. doi: 10.3389/fnagi.2020.00038 |
|  | T2D | Vernochet C. 2012. doi: 10.1016/j.cmet.2012.10.016 |
|  | Obesity |  |
|  | Phosphoprotein | Reardon S. 2022. doi: 10.1016/j.jbc.2022.101815 |
| ***TFAM*** | Obesity | Reardon SD et al. 2022. doi: 10.1016/j.jbc.2022.101815 |
|  | TF | Ngo HB et al. 2011. doi: 10.1038/nsmb.2159 |
| ***TFB2M*** | Glycolysis | Chang H et al. 2021. doi: 10.1111/jgh.15548 |
|  | Phosphoprotein | Bostwick AM et al 2020. doi: 10.1016/j.bbrc.2020.05.141 |
|  | Kinase cascades | Geng X et al. 2020. doi: 10.1111/liv.14440 |
|  | NFκB |  |
|  | Chromatin | Watanabe A et al, 2011. doi: 10.1093/cvr/cvq374 |
| ***TFE3*** | Cytokines | Pastore N et al. 2016. doi: 10.1080/15548627.2016.1179405 |
|  | Inflammation | Li X et al. 2023. doi: 10.1038/s41420-023-01395-0 |
|  | Insulin signaling | Iwasaki H et al. 2012. doi: 10.1152/ajpendo.00204.201 |
|  | Glycolysis | Jeong E et al. 2022. doi: 10.1080/15548627.2022.2029671 |
| ***TGFBR2*** | Microglial neuroinflammation | Hagemeyer N et al, 2014. doi: 10.15252/embj.201490345 |
|  | M1 polarization |  |
|  | *Irf7* transcription |  |
|  | Cognition | El Hamamy A et al. 2024. doi: 10.21203/rs.3.rs-4438544/v1 |
| ***TGIF1*** | Inflammation | Hneino M et al. 2012. doi: 10.1074/jbc.M112.388389 |
|  | DM | Friend CJ, 2023. https://scholarscompass.vcu.edu/etd/7513/ |
|  | Phosphoprotein | Chang YH et al. 2024. doi: 10.1002/1873-3468.14849 |
|  | Lipids | Pramfalk C et al. 2015. doi: 10.1016/j.bbalip.2014.07.019 |
|  | TF | NIH, 2025. [https://www.ncbi.nlm.nih.gov/gene/7050](https://protect.checkpoint.com/v2/r01/___https://www.ncbi.nlm.nih.gov/gene/7050___.YzJ1OmJpb3ZpZTE6YzpvOjljNzMyZTM5MjY4ZDg1MDA1MDcyODAxYjc4YTlmZjRhOjc6MGJiYTo4MGNhYzE1NTViZGIyN2YwODRlYzBlY2FmNWNjNTA3NDM1NmQyY2IzNDE5MGYzMjhjOWYzYTU4NzA3YjZkODk0OnA6RjpG) |
|  | Chromatin | He X et al. 2021. doi: 10.3390/ijms22147452 |
| ***TIGD1*** | AD | Jönsson M et al. 2020. doi: 10.1016/j.tig.2020.05.004 |
|  | Inflammation | Wang L et al. 2024. doi: 10.1101/gr.280357.124 |
|  | Chromatin | Paul S K et al. 2024. doi: 10.1038/s41467-024-48663-w  Rostami M R et al. 2021. doi: 10.1186/s13100-021-00241-3 |
|  | Immune imbalance |  |
|  | AD | Feng Y et al. 2024. doi: 10.1002/alz.14164 |
|  | Microglial neuroinflammation | Roy N et al. 2024. doi: 10.1007/s00401-024-02835-6 |
|  | Tau-transposable elements | Guo C et al. 2018. doi: 10.1016/j.neurobiolaging.2017.11.003  Evering T H et al. 2023. doi: 10.1016/j.tins.2022.12.003 |
|  | Cognition | Ahmadi A et al. 2020. doi: 10.1016/j.arr.2020.101153 |
| ***TMBIM6*** | T2D | Li B. et al. 2014. doi: 10.2174/1566524014666140603101113 |
|  | Obesity |  |
|  | Cognition |  |
|  | Significantly ↑ AD microglial transcripts with Dementia | Kosoy R et al, 2025. doi:10.1038/s41593-025-02000-6 |
| ***TMED10*** | Inflammation | Liu L et al. 2024. doi:10.1038/s41467-024-52299-1 |
|  | Significantly ↑ AD microglial transcripts with Dementia | Kosoy R et al, 2025. doi:10.1038/s41593-025-02000-6 |
|  | Braak* |  |
| ***TMEM127*** | T2D | Srikantan S et al. 2019. doi: 10.1038/s41467-019-12661-0 |
|  | Obesity |  |
|  | Astrocyte neuroinflammation | Leonard J et al. 2024. doi: 10.1038/s41598-024-58904-z |
| ***TMEM14C*** | Significantly ↑ AD microglial transcripts with Dementia | Kosoy R et al. 2025. doi:10.1038/s41593-025-02000-6 |
|  | Braak* |  |
| ***TNFRSF10A*** | Brain amyloid-β load, apoptosis, cognition, microglial neuroinflammation | Frenkel D et al. 2015. doi: 10.1093/brain/awu334  Cantarella G et al. 2015. doi: 10.1093/brain/awu318. |
|  | M1 polarization | Lin S et al. 2021. doi: 10.1016/j.freeradbiomed.2021.07.014 |
|  | Obesity | Karason K et al. 2022. doi: 10.1038/s41366-022-01194-0. |
|  | Significantly ↑ AD Plasma Proteomics | Ali M et al, 2025. doi: 10.1038/s41591-025-03833-1. Erratum: doi: 10.1038/s41591-025-03970-7 |
| ***TNFRSF10*** | Aβ | Frenkel D et al. 2015. doi: 10.1093/brain/awu334  Cantarella G et al. 2015. doi: 10.1093/brain/awu318 |
|  | Apoptosis |  |
|  | Cognition |  |
|  | Microglial neuroinflammation |  |
|  | M1 polarization | Lin S et al. 2021. doi: 10.1016/j.freeradbiomed.2021.07.014 |
|  | Obesity | Karason K et al. 2022. doi: 10.1038/s41366-022-01194-0 |
|  | Significantly ↑ AD Plasma Proteomics | Ali M, et al, 2025. doi: 10.1038/s41591-025-03833-1. Erratum: doi: 10.1038/s41591-025-03970-7 |
| ***TNIP2*** | Neuroinflammation, cognition, MDD | Chiang T et al. 2021. doi: 10.1016/j.bbi.2021.04.021. |
| ***TOMM40*** | Microglial neuroinflammation | Chen Y-C et al. 2023. doi: 10.3390/ijms24044085 |
|  | M1 polarization |  |
|  | Cognition | Gottschalk W K et al. 2014. doi: 10.13188/2376-922X.1000003  Gui W et al. 2021. doi: 10.3389/fpsyt.2021.617773 |
|  | Lipids | Yang NV et al. 2024. doi: 10.1016/j.molmet.2024.102056 |
|  | T2D | Greenbaum L et al. 2014. doi: 10.1016/j.euroneuro.2014.06.002 |
|  | Aging | Kulminski AM et al. 2019. doi: 10.1111/acel.12869. |
|  | Obesity |  |
| ***TP53*** | Aggregation | Farmer KM et al. 2020. doi: 10.1186/s40478-020-01012-6 |
|  | pTau |  |
|  | DNA damage |  |
|  | Phosphoprotein |  |
|  | Microglial neuroinflammation | Aloi MS et al. 2015. doi: 10.1615/critrevimmunol.v35.i5.40 |
|  | MicroRNAs |  |
|  | Inflammation | Holtman IR et al. 2017. doi: 10.1172/JCI90604 |
|  | T2D | Strycharz J et al. 2017. doi: 10.1155/2017/9270549 |
|  | Obesity | Zwezdaryk K et al. 2018. doi: 10.3389/fendo.2018.00457 |
|  | TF | Hernández Borrero LJ, El-Deiry WS. 2021. doi: 10.1016/j.bbcan.2021.188556 |
|  | Chromatin | Serra F et al. 2024. doi: 10.1038/s41467-024-46666-1 |
| ***TPI1*** | AD | Tajes M et al. 2014. doi: 10.3233/JAD-131685 |
|  | Cognition |  |
|  | Obesity | Varma V et al. 2008. doi: 10.2337/db07-0840 |
| ***TRADD*** | Microglial neuroinflammation | Hassan M et al. 2021. doi: 10.1038/s41598-021-91606-4  NIH. 2025. https://www.ncbi.nlm.nih.gov/Structure/cdd/cd10576#:~:text=TNFRSF1A,-Entrez&text=TNFRSF1A%20(also%20known%20as%20type,D12E)%20in%20the%20TNFRSF1A%20gene.&text=Conserved%20Features/Sites%20?,PubMed%20References  Steeland S et al. 2018. doi: 10.15252/emmm.201708300 |
|  | Apoptosis |  |
|  | NFκB |  |
|  | M1 polarization | Perez S, Rius-Perez S. 2022. doi: 10.3390/antiox11071394 |
|  | T2D | Nault JC, Zucman-Rossi J, 2010. doi: 10.1016/j.jhep.2010.05.011 |
|  | Obesity |  |
| ***TRAF6*** | Microglial neuroinflammation | Liu Y et al. 2023. doi: 10.1007/s12035-023-03453-8 |
|  | NFκB |  |
|  | Cognition | Wang S et al. 2018. doi: 10.1007/s12031-018-1032-3 |
|  | M1 polarization | Zhao Y et al. 2024. doi: 10.1007/s10565-024-09900-6 |
|  | Significantly ↑FTD proteomics | Ali M et al, 2025. doi: 10.1038/s41591-025-03833-1. Erratum: doi: 10.1038/s41591-025-03970-7 |
| ***TRIM25*** | Inflammation | Liu Y et al. 2020. doi: 10.4049/jimmunol.1900482 |
|  | TNF |  |
|  | NFκB |  |
|  | M1 polarization | Wu H et al. 2024. doi: 10.1007/s00011-024-01906-4 |
|  | Atherosclerosis |  |
|  | Apoptosis |  |
|  | Glycolysis | Li C et al. 2022. doi: 10.3390/ijms23169325 |
|  | Lipids | Zhang H et al. 2025. doi: 10.1002/advs.202414646 |
|  | Phosphoprotein | Lee N-R et al. 2018. doi: 10.1016/j.cellimm.2018.08.004 |
| ***TRIM26*** | Kinase cascades | Zhoa J et al, 2021. https://doi.org/10.1038/s41418-021-00803-1  Zou J et al, 2024. doi: 10.2174/0109298665311516240621114519 |
|  | NFκB |  |
|  | Chromatin |  |
| ***TRIM27*** | Brain aging | Kang J et al, 2022. [https://doi.org/10.1101/2022.12.30.22284052](https://protect.checkpoint.com/v2/r01/___https://doi.org/10.1101/2022.12.30.22284052___.YzJ1OmJpb3ZpZTE6YzpvOjljNzMyZTM5MjY4ZDg1MDA1MDcyODAxYjc4YTlmZjRhOjc6NDNhMzpiYjZhNmZhNDI4ZWIxY2VmMmVkNGZhMTA1NGZmMTA4NTM4MGQwNmM5NmVkNzNkYTMxMzg1MDdhMGY4ZjQ3NDU1OnA6RjpG)  Li P et al, 2025. doi: 10.1016/j.ibneur.2025.01.001 |
|  | Obesity |  |
|  | Cognition |  |
|  | Microglial neuroinflammation | Wang C et al. 2023. doi: 10.1016/j.jchemneu.2023.102251 |
|  | Significantly ↑AD proteomics | Ali M et al, 2025. doi: 10.1038/s41591-025-03833-1. Erratum: doi: 10.1038/s41591-025-03970-7 |
| ***TRIM52*** | Inflammation | Ma J-P et al. 2023. doi: 10.15586/aei.v51i1.737  Zhang P et al. 2020. doi: 10.12659/MSM.925356 |
|  | TLR4 |  |
|  | NFκB |  |
|  | Kinase cascades |  |
|  | Pyroptosis |  |
| ***TRMT112*** | Inflammation | Zhou D et al. 2024. doi: 10.1038/s12276-024-01315-x |
|  | Significantly ↑AD & PD plasma proteomics | Ali M et al, 2025. doi: 10.1038/s41591-025-03833-1. Erratum: doi: 10.1038/s41591-025-03970-7 |
| ***TMX3*** | Protein misfold | Honjo Y et al, 2014. doi: 10.3233/JAD-130632 |
|  | Neurofibrillary tangles |  |
|  | Apoptosis |  |
|  | Vascular inflammation | Cho J. 2013, doi: 10.1111/jth.12413 |
|  | Thrombosis |  |
|  | Significantly ↑AD plasma proteomics | Ali M et al, 2025. doi: 10.1038/s41591-025-03833-1. Erratum: doi: 10.1038/s41591-025-03970-7 |
| ***TRABD*** | Microglial neuroinflammation | Duan W et al, 2025. doi: 10.3389/fcell.2025.1619339 |
|  | Tau toxicity |  |
|  | Significantly ↑AD plasma proteomics | Ali M et al, 2025. doi: 10.1038/s41591-025-03833-1. Erratum: doi: 10.1038/s41591-025-03970-7 |
| ***TSC2*** | Cognition | Tang G. 2022. https://protect.checkpoint.com/v2/r01/___https://cdmrp.health.mil/tscrp/research_highlights/22Guomei_Tang_highlight___.YzJ1OmJpb3ZpZTE6YzpvOjljNzMyZTM5MjY4ZDg1MDA1MDcyODAxYjc4YTlmZjRhOjc6NWI4MDowNTE2MDE1YTg5N2NiM2ZkMTY5MTQzMjc5MTJhNGMxNDIyNzRjMzUzNjYwYjE4NTkyOWUwYjc1M2QxNWJmNGRkOnA6RjpG" \l ":~:text=Tuberous%20sclerosis%20complex%20(TSC)%20is%20a%20multi%2Dsystem,in%20either%20the%20TSC1%20or%20TSC2%20gene |
|  | Microglial neuroinflammation | Kagitani-Shimono K et al. 2023, doi: 10.1016/j.nicl.2022.103288 |
|  | Cognition |  |
|  | T2D | Jurca CM et al, 2023. doi: 10.3390/genes14020433 |
| ***TSC22D1*** | Inflammation | Ding W et al, 2025. doi: 10.3748/wjg.v31.i31.109605. |
|  | M1 polarization |  |
|  | Lipids |  |
|  | TF | NIH, 2025. https://www.ncbi.nlm.nih.gov/gene/8848 |
| ***TSC22D4*** | AD | Yasukawa T et al, 2020. doi: 10.1016/j.celrep.2020.02.059  Kuhn MK et al, 2023. doi: 10.1101/2023.04.07.536014 |
|  | Microglial neuroinflammation |  |
|  | TF |  |
|  | Aβ |  |
|  | Cytokines |  |
|  | Phosphoprotein | Demir S et al, 2022. doi: 10.1126/sciadv.abo5555 |
|  | T2D |  |
|  | IR |  |
|  | Lipids | Jones et al, 2013, doi: 10.1002/emmm.201201869 |
|  | M1 polarization |  |
| ***TSPAN33*** | Inflammation | Ruiz-Garcia A et al, 2016. doi: 10.4049/jimmunol.1600421 |
|  | Kinase cascades |  |
|  | NFκB |  |
| ***TUBA1B*** | Inflammation | Hu X et al. 2022. doi: 10.3390/diagnostics12040858 |
|  | Significantly ↑ AD microglial transcripts with Dementia | Kosoy R et al, 2025. doi:10.1038/s41593-025-02000-6 |
| ***TUBA4A*** | Microglial neuroinflammation | Hausrat TJ et al, 2022. doi: 10.1038/s41467-022-31776-5 |
|  | AD |  |
|  | pTau |  |
| ***TXNIP*** | Inflammation | Tsubaki H et al, 2020. doi: 10.3390/ijms21249357 |
|  | NLRP3 |  |
|  | Neurodegeneration |  |
|  | Microglial neuroinflammation | Sbai O et al, 2022. doi: 10.1038/s41419-022-04758-0 |
|  | Cognition | Yang C et al, 2024. doi: 10.1016/j.heliyon.2024.e27423 |
|  | T2D | Lu B et al, 2022. doi: 10.1210/endocr/bqac133 |
| ***TYW3*** | T2D | Qi, Q et al, 2012. doi: 10.1093/hmg/dds300 |
|  | PD | Akrioti E et al, 2022. doi: 10.3390/biom12070876 |
|  | ALS | Wei L et al, 2019. doi: 10.1212/NXG.0000000000000375 |
| ***UBA7*** | Microglial neuroinflammation | Przanowski P et al, 2018. doi: 10.1016/j.neuint.2017.07.013 |
| ***UBE2C***  *Promoter DNA methylation was decreased (potentially increased expression) in bezisterim vs placebo subjects* | Inflammation | Elliott PJ et al, 2003. doi: 10.1007/s00109-003-0422-2  Chitra S at l, 2012. doi: 10.1111/j.1756-185X.2012.01737.x |
|  | NFκB |  |
|  | Proteosome | Parker D et al, 2025. doi: 10.1111/acel.14492 |
|  | Cognition |  |
|  | Significantly ↓AD plasma proteomics | Ali M et al, 2025. doi: 10.1038/s41591-025-03833-1. Erratum: doi: 10.1038/s41591-025-03970-7 |
| ***UBE2V1*** | Inflammation | Xu N et al. 2020. doi: 10.1161/CIRCRESAHA.119.316444  Li L et al, 2025. doi: 10.1038/s42003-025-08214-5  UniProt, 2025. [https://www.uniprot.org/uniprotkb/Q13404/entry#:~:text=Function-,function,production%20(PubMed:31006531)](https://protect.checkpoint.com/v2/r01/___https://www.uniprot.org/uniprotkb/Q13404/entry___.YzJ1OmJpb3ZpZTE6YzpvOjljNzMyZTM5MjY4ZDg1MDA1MDcyODAxYjc4YTlmZjRhOjc6MDM4MToyZGNkNmZiNTgwMGJhODU3MjE1ZDI5M2U5ZmUzMWEzNmM0YzY1ZWQyODllYTQzNTE1NmQwYjQzYzZlNjNjZGEwOnA6RjpG#:~:text=Function-,function,production%20(PubMed:31006531)) |
|  | Protein aggregation |  |
|  | NFκB | Parker D et al, 2025. doi: 10.1111/acel.14492 |
|  | Cognition |  |
| ***USF1*** | Inflammation | Zhuang S et al, 2025. doi: 10.1016/j.bbalip.2024.159581 |
|  | ROS |  |
|  | Lipids |  |
|  | NFκB | Song X et al, 2018. doi: 10.3892/etm.2018.6608 |
|  | Lipids | Wu S et al, 2010. doi: 10.1093/hmg/ddp526 |
|  | T2D |  |
|  | Obesity |  |
|  | Cognition | Sideromenos S et al, 2022. doi: 10.1038/s41398-022-02266-5 |
|  | Kinase cascades | Horbach T et al, 2015. doi: 10.3389/fphar.2015.00003. Erratum: 2016. doi: 10.3389/fphar.2016.00092 |
|  | Phosphoprotein | Lupp S et al, 2014. doi: 10.1016/j.cellsig.2014.08.028 |
|  | TF | NIH, 2025. <https://www.ncbi.nlm.nih.gov/gene/7391#:~:text=USF1%20and%20USF2%20are%20important,1%20to%20the%20SNP%20site> |
| ***USP19*** | Obesity | Coyne ES et al, 2019. doi: 10.1007/s00125-018-4754-4 |
|  | T2D |  |
|  | Lipids | Zhu Y et al, 2021. doi: 10.1016/j.celrep.2021.110174 |
|  | PD | Schorova L et al, 2023. doi: 10.1038/s41531-023-00601-1 |
|  | Α-synuclein |  |
| ***USP22*** | Lipids | Ning Z et al, 2022. doi: 10.1038/s41467-022-29846-9 |
|  | Glycolysis | Chen S et al, 2024. doi: 10.1111/jcmm.70239 |
| ***USP3*** | Inflammation | Zhuang W et al, 2022. doi: 10.1038/s41423-022-00917-7 |
|  | Significantly ↑ AD Plasma Proteomics | Ali M, et al, 2025. doi: 10.1038/s41591-025-03833-1. Erratum: doi: 10.1038/s41591-025-03970-7 |
|  | Infammation |  |
| ***YWHAG*** | AD | Oh HS et al, 2025. doi: 10.1038/s41591-025-03565-2 |
|  | Cognition |  |
|  | Inflammation | Autieri MV et al, *Cell Growth Differ*. 1996 Nov;7(11):1453-60. |
|  | Kinase cascades | Tian T et al, 2024. doi: 10.1186/s12967-024-06003-y |
| ***YWHAZ*** | pTau | Qiang Q et al, 2024. doi: 10.1016/j.jns.2023.122861 |
|  | Cognition |  |
|  | Inflammation |  |
|  | Glycolysis | Shi J et al, 2019. doi: 10.3892/or.2018.6920 |
|  | Obesity | Rial SA et al, 2025. doi: 10.1016/j.molmet.2025.102159 |
|  | IR |  |
| ***ZBTB4*** | T2D | Song Y et al, 2022. doi: 10.3389/fgene.2022.1015879 |
|  | AD (onset age) | NIAGADS, 2025. [https://www.niagads.org/publications/variants-regulating-zbtb4-are-associated-with-age-at-onset-of-alzheimers-disease/#:~:text=Variants%20regulating%20ZBTB4%20are%20associated,onset%20of%20Alzheimer's%20disease%20%E2%80%93%20NIAGADS](https://protect.checkpoint.com/v2/r01/___https://www.niagads.org/publications/variants-regulating-zbtb4-are-associated-with-age-at-onset-of-alzheimers-disease/___.YzJ1OmJpb3ZpZTE6YzpvOjljNzMyZTM5MjY4ZDg1MDA1MDcyODAxYjc4YTlmZjRhOjc6Zjk1ZTpiYTY0NDdlNGU0OWVjZjg2N2NkNTgzZDhlOThlZThjMzRhNjNjODcyMzAwZWVlNmMzYTkyMjJjZmZiMTY1ZjI5OnA6RjpG#:~:text=Variants%20regulating%20ZBTB4%20are%20associated,onset%20of%20Alzheimer's%20disease%20%E2%80%93%20NIAGADS) |
|  | TF | Yang WS et al, 2014. doi: 10.1016/j.neo.2014.09.011 |
| ***ZCCHC17*** | Cognition | Chen PC et al, 2022. doi: 10.1038/s43587-022-00290-0. |
|  | Neuroexcitability |  |
|  | Significantly ↑AD plasma proteomics | Ali M et al, 2025. doi: 10.1038/s41591-025-03833-1. Erratum: doi: 10.1038/s41591-025-03970-7 |
| ***ZDHHC14*** | Lipids | Natale F et al, 2024. doi: 10.1073/pnas.2402604121 |
|  | Cognition |  |
| ***ZEB1*** | Microglial neuroinflammation | Poonaki E et al, 2022. doi: 10.1186/s12974-022-02636-2 |
|  | M1 polarization | Jiang H et al, 2022. doi: 10.1038/s41419-022-04632-z |
|  | Glycolysis |  |
|  | TF | Wang Y et al, 2023. doi: 10.1038/s41467-023-42428-7 |
| ***ZHX2*** | Atherosclerosis | Erbilgin A et al. 2018. doi: 10.1161/ATVBAHA.118.311266 |
|  | Apoptosis |  |
|  | Lipids |  |
|  | Inflammation |  |
|  | M1 polarization | Tan S et al. 2023. doi: 10.1038/s41418-023-01202-4 |
|  | TF |  |
|  | Glycolysis | Wang Z et al, 2020. doi: https://doi.org/10.4049/jimmunol.1901246 |
|  | Significantly ↑ AD microglial transcripts with Dementia | Ali M, et al, 2025. doi: 10.1038/s41591-025-03833-1. Erratum: doi: 10.1038/s41591-025-03970-7 |
| ***ZMYM3*** | TF | Kosoy R et al, 2025. doi: 10.1038/s41593-025-02020-2 |
|  | Chromatin | Hiatt SM et al. 2023. doi: 10.1016/j.ajhg.2022.12.007. |
|  | Inflammation |  |
| ***ZNF395*** | Phosphoprotein | Herwartz C et al. 2015. doi: 10.1155/2015/804264. |
|  | TF | Jordanovski D et al. 2013. doi: 10.1371/journal.pone.0074911. |
|  | Kinase cascades |  |
|  | Lipids |  |
|  | Obesity | Hasegawa R et al. 2013. doi: 10.1016/j.yexcr.2012.11.003. |

α-KG, alpha ketoglutyrate; Aβ, amyloid beta; AD, Alzheimer’s disease; ALS, amyotrophic lateral sclerosis; AP-1, activator protein 1; APP, amyloid precursor protein; ATR, ataxia-telangiectasia and Rad3-related; CVD, cardiovascular disease; CYP, cytochrome P450 enzymes; DM, diabetes mellitus; EPO, erythropoietin; ERK, extracellular signal-related kinases; FABP7, fatty acid–binding protein 7; FFA, free fatty acids; FTD, frontotemporal dementia; Glut1, glucose transporter 1; GM1, ganglioside monosialotetra-hexoganclioside; HHcy, Hyperhomocysteinemia; HTN, hypertension; H3K4me3, histone HS lysine trimethylation; HDAC1, histone deacetylase 1; Hebp1, heme-binding protein 1; HHcy, hyperhomocysteinemia; HIF1α, hypoxia-inducible factor 1 alpha; HTN, hypertension; IFN, interferon; IR, insulin resistance; IRF, interferon regulatory factor; IRS1, insulin receptor substrate 1; LOAD, late-onset Alzheimer’s disease; M1, macrophage “classical” activation; M2, macrophage apoptosis; MAPK, mitogen-activated protein kinase; MCP1, monocyte chemoattractant protein-1; MEF2C, myocyte-specific enhancer factor 2C; mTOR, mammalian target of rapamycin; NLRP, nucleotide-binding oligomerization domain-like receptor family pyrin domain-containing proteins; NOD2, nucleotide-binding oligomerization domain-containing protein 2; NRF2, nuclear factor-erythroid 2-related factor 2; PD, Parkinson’s disease; PDHA1, phosphorylated pyruvate dehydrogenase E1α; pTau, phosphorylated tau; RIPK1, receptor-interacting serine/threonine-protein kinase 1; ROS, reactive oxygen species; SLAMF8, signaling lymphocytic activation molecule family member 8; T2D, type 2 diabetes; T3D, type 3 diabetes; TF, transcription factor; TLR, toll-like receptor; TNF, tumor necrosis factor.
